# Mayo Normative Studies: regression-based normative data for remote self-administration of the Stricker Learning Span, Symbols Test and Mayo Test Drive Screening Battery Composite and validation in individuals with Mild Cognitive Impairment and dementia

**DOI:** 10.1101/2024.09.14.24313641

**Authors:** Nikki H. Stricker, Ryan D. Frank, Elizabeth A. Boots, Winnie Z. Fan, Teresa J. Christianson, Walter K. Kremers, John L. Stricker, Mary M. Machulda, Julie A. Fields, John A. Lucas, Jason Hassenstab, Paula A. Aduen, Gregory S. Day, Neill R. Graff-Radford, Clifford R. Jack, Jonathan Graff-Radford, Ronald C. Petersen

**Author notes:** Corresponding Author: Nikki H. Stricker, Ph.D., ABPP-CN, Mayo Clinic, 200 First Street SW, Rochester, MN 55905, (email).

## Abstract

**Objective:** Few normative data for unsupervised, remotely-administered computerized cognitive measures are available. We examined variables to include in normative models for Mayo Test Drive (a multi-device remote cognitive assessment platform) measures, developed normative data, and validated the norms.

**Method:** 1240 Cognitively Unimpaired (CU) adults ages 32-100-years (96% white) from the Mayo Clinic Study of Aging and Mayo Alzheimer’s Disease Research Center with Clinical Dementia Rating® of 0 were included. We converted raw scores to normalized scaled scores and derived regression-based normative data adjusting for age, age^2^, sex and education (base model); alternative norms are also provided (age+age^2^+sex; age+age^2^). We assessed additional terms using an *a priori* cut-off of 1% variance improvement above the base model. We examined low test performance rates (<-1 standard deviation) in independent validation samples (n=167 CU, n=64 mild cognitive impairment (MCI), n=14 dementia). Rates were significantly different when 95% confidence intervals (CI) did not include the expected 14.7% base rate.

**Results:** No model terms met the *a priori* cut-off beyond the base model, including device type, response input source (e.g., mouse, etc.) or session interference. Norms showed expected low performance rates in CU and greater rates of low performance in MCI and dementia in independent validation samples.

**Conclusion:** Typical normative models appear appropriate for remote self-administered MTD measures and are sensitive to cognitive impairment. Device type and response input source did not explain enough variance for inclusion in normative models but are important for individual-level interpretation. Future work will increase inclusion of individuals from under-represented groups.

## Introduction

Normative data (i.e., norms) provide an estimate of where an individual’s test performance falls relative to other individuals. Norms are a critical component of neuropsychological tests to aid individual level interpretation of performance, helping to determine normal versus abnormal performance in conjunction with expert clinician judgment. Despite their importance, they are often only available in technical manuals without undergoing peer review. Further, many computerized tests do not have publicly available normative data despite the availability of norms for users (Alden et al., 2021), or are briefly mentioned in supplements to manuscripts (Tsoy et al., 2020). Peer-reviewed norms are available for only select computerized neuropsychological tests, and these typically represent in-clinic administration (Casaletto et al., 2015; Gualtieri & Johnson, 2006). Normative data for self-administered computerized measures completed in unsupervised remote environments are starting to become available (Singh et al., 2021) but remain relatively limited and frequently do not include platforms that offer smartphone compatibility (Feenstra, Vermeulen, Murre, & Schagen, 2018; Stiver et al., 2024; Visser et al., 2021).

Mayo Test Development through Rapid Iteration, Validation and Expansion (Mayo Test Drive, MTD) is a cognitive testing platform developed for remote self-administered digital cognitive assessment (J. L. Stricker et al., 2022; N. H. Stricker et al., 2022). MTD is a web-based, multi-device compatible (smartphone, tablet, desktop/laptop computer) platform that can be easily accessed and completed by individuals being assessed. MTD typically takes 15-20 minutes to complete and shows high usability (98.5% completion rates remotely (Patel et al., 2024)) and adequate test-retest reliability (Hughes et al., 2024; N. H. Stricker et al., 2022). Additionally, MTD’s cognitive subtests provide more in-depth assessment of targeted cognitive domains relative to traditional screening tests. These subtests include the Stricker Learning Span (SLS), a novel computer adaptive word list memory test with learning and delay trials (J. L. Stricker et al., 2022; N. H. Stricker, Stricker, et al., 2024) and the Symbols Test, an open-source measure of visual matching and processing speed/executive function (Boots et al., 2024; Nicosia et al., 2022). The SLS shows significant associations with an in-person memory measure, the Rey Auditory Verbal Learning Test (RAVLT) (Boots et al., 2024). The SLS also shows significant associations with amyloid and tau PET, hippocampal volume, and white matter hyperintensities (Boots et al., 2024), as well as entorhinal cortical thickness (N. H. Stricker et al., 2022). The SLS differentiates individuals with a biological definition of Alzheimer’s disease (AD) based on PET imaging markers similarly to the RAVLT and shows sensitivity to preclinical AD (N. H. Stricker, Stricker, et al., 2024). The Symbols test shows significant associations with in-person processing speed/executive functioning measures including Trail Making Test B and Wechsler Adult Intelligence Scale-Revised (WAIS-R) Digit Symbol Coding (Boots et al., 2024). The SLS and Symbols Test are combined into an MTD screening battery composite total raw score (MTD Composite) (Boots et al., 2024). Both the MTD Composite and SLS show large effect sizes for differentiating individuals with MCI/dementia from cognitively unimpaired individuals (Boots et al., 2024). In summary, MTD shows promise for clinical utility and will benefit from published norms to further support clinical use.

Normative data commonly adjust for key demographic variables such as age, sex and education to provide a more tailored estimate of an individual’s deviation from expected performance.

Considering adjustment for factors beyond demographics may be needed in developing normative data for remote self-administered assessments. For example, environmental context, interruptions, device type, and response input source used (e.g., mouse versus touch) all have the potential to impact test performance. We previously showed significantly faster response times in individuals completing the Cogstate Brief Battery in clinic on a desktop/laptop computer with a mouse compared to when completed on iPads with touch response in clinic, but there were no differences across devices on measures of accuracy (visual memory, working memory); longitudinal trajectories were also largely similar for both device types (Stricker et al., 2019). Our prior work also demonstrated lower performance accuracy on a visual memory measure (Cogstate One Card Learning) when completed at home compared to in clinic (N. H. Stricker et al., 2020); however, many studies comparing remote and in-clinic performances of computerized measures have shown no performance differences (Backx, Skirrow, Dente, Barnett, & Cormack, 2020; Cromer et al., 2015). It is possible that our isolated finding of worse at-home performance may have been due to increased frequency of environmental distractions in the home environment. For example, Madero and colleagues (Madero et al., 2021) showed that about 7% of participants experienced environmental distraction defined as looking away from a camera using eye tracking while completing a 5-minute visual paired-comparison task, and primary outcome scores were about 3% lower for participants who were distracted. Many platforms have self-report questions to help inform the potential for environmental distractions to influence performance, but no studies have examined whether there is a need to incorporate this into normative models for remote assessment measures.

The aim of this study was to develop normative data for measures completed in predominantly unsupervised remote environments via the MTD platform including the Stricker Learning Span (SLS), Symbols Test, and MTD Composite. We hypothesized that MTD measures would show significant relationships with age, sex, and education. We examined whether other variables may be necessary to include in normative models given the digital remote self-administration emphasis, including device type, response input source, and potential interference or distraction. We evaluated normative model performance by applying normative data to independent samples (cognitively unimpaired, mild cognitive impairment, and dementia) to provide initial validation data for these norms.

## Method

### Participants

The MTD study is an ancillary protocol that recruits from collaborating parent studies. Most participants in the current study were recruited from the Mayo Clinic Study of Aging (MCSA). The MCSA is population-based study of aging in Olmsted County, Minnesota; participants are randomly sampled by age- and sex-stratified groups using the resources of the Rochester Epidemiology Project medical records-linkage system (St Sauver et al., 2012; St. Sauver, Grossardt, Yawn, Melton, & Rocca, 2011). MCSA exclusion criteria are having a terminal illness or receiving hospice care. Study visits consist of a physician examination that includes administration of the Short Test of Mental Status (Kokmen, Smith, Petersen, Tangalos, & Ivnik, 1991), study coordinator interview that includes the Clinical Dementia Rating (CDR®) instrument (Morris, 1993) and neuropsychological testing supervised by a clinical neuropsychologist (MMM, JAF) (Roberts et al., 2008), each of whom makes an independent diagnostic determination of CU, MCI, or dementia. Only the neuropsychologist has access to in-person neuropsychological data for this diagnostic determination. Following independent diagnostic assessment, a final diagnosis of cognitively unimpaired, MCI (Petersen, 2004), or dementia (American Psychiatric Association, 1994) is determined through consensus agreement (Petersen, 2004; Roberts et al., 2008). Mayo Test Drive is independent of diagnosis (e.g., the data are not available for review at the time of diagnosis). The MCSA diagnostic evaluation also does not consider prior clinical information, prior diagnoses, or knowledge of biomarker status. Further details about the MCSA study protocol are available (Roberts et al., 2008). Additional participants were recruited from the Mayo Alzheimer’s Disease Research Center (ADRC) in Rochester, MN and Jacksonville, FL. The diagnostic procedure for ADRC participants is similar to MCSA in that data from a neurologic evaluation, mental status examination, CDR®, and neuropsychological assessments are independently reviewed by respective specialists, with final diagnosis determined by consensus agreement. Unlike MCSA procedures, however, the final consensus diagnosis does reflect prior clinical information, prior diagnoses and biomarker status.

This study was completed in accordance with the Helsinki Declaration. Study protocols were approved by the Mayo Clinic Institutional Review Board (IRB); the MCSA parent study protocol also includes approval by the Olmsted Medical Center IRB. All participants provided written informed consent for the primary study protocols (MCSA, ADRC); oral consent (provided after reading informed consent elements sent in an email or described verbally) was obtained for the ancillary Mayo Test Drive study protocol. No compensation was provided for participation in the ancillary study.

### Inclusion Criteria for Normative Sample

The inclusion criteria for the normative sample included: (1) baseline MTD session; (2) concordant CU diagnosis for MCSA participants, meaning that the study coordinator, examining physician, and neuropsychologist each independently assigned a CU diagnosis; a consensus diagnosis of CU was used for ADRC participants; (3) Global CDR® of 0; (4) MTD Composite not missing (e.g., all SLS and all Symbols Test data available); (5) valid session; and (6) test naïve, defined as not having been exposed to MTD prior to participation in the current study. We prioritized stringent inclusion criteria (e.g., all evaluators assigning a CU diagnosis, CDR=0) as our prior work has demonstrated the importance of careful refinement of normative samples to improve sensitivity to MCI and dementia (N. H. Stricker, Christianson, et al., 2024). See **Figure 1** for a summary of participant exclusions. Baseline MTD sessions for all included individuals were completed between May 25, 2021 and April 30, 2023 using MTD platform v1 (1.0104-1.0111).

**Figure 1.**
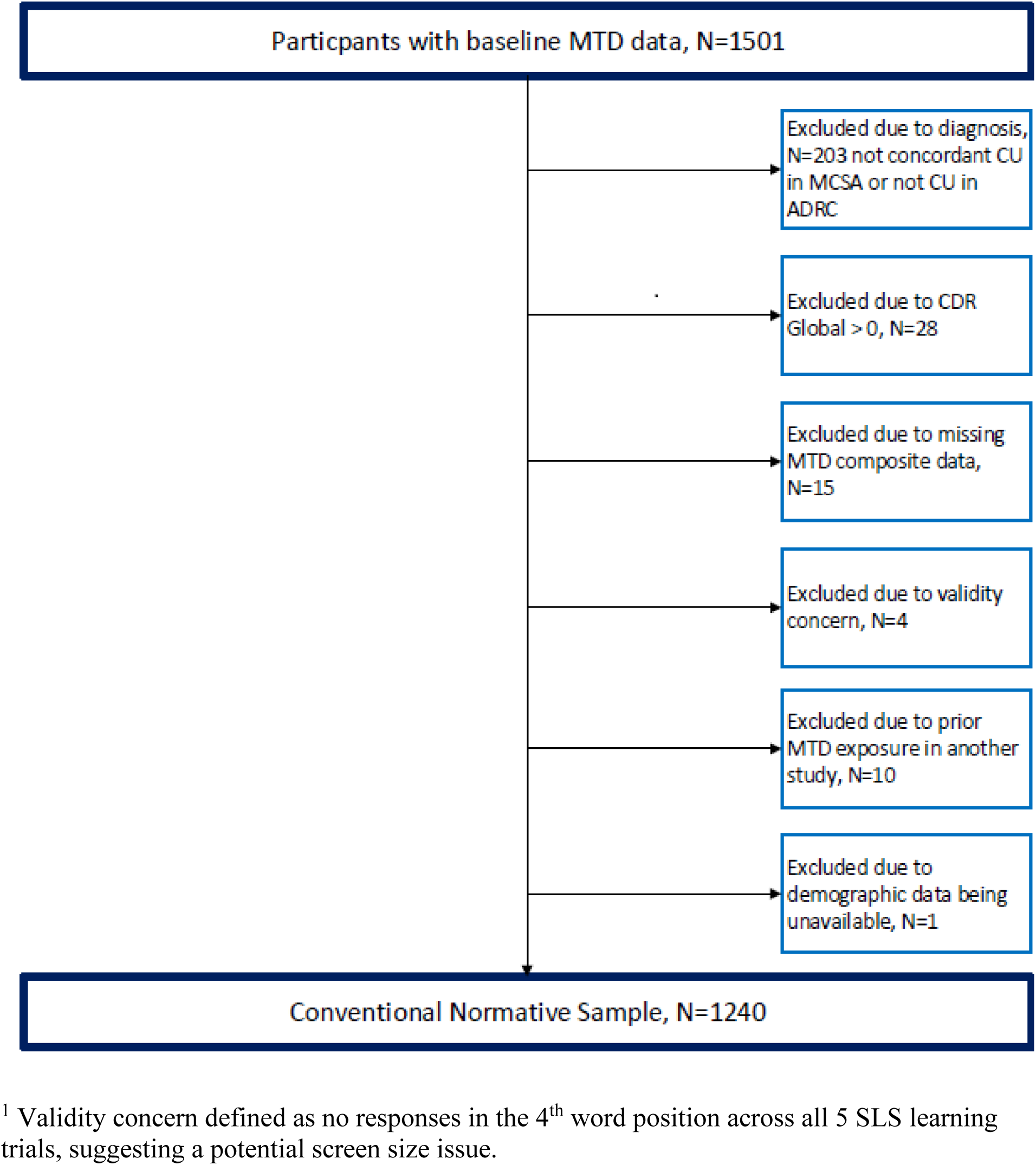
Flow chart showing participant selection for normative data.

### Inclusion Criteria for Independent Validation Samples

We derived independent validation samples to examine performance of normative data models. Baseline MTD sessions completed between May 25, 2021, and December 14, 2023, with all SLS and Symbols data available (MTD Composite not missing) and no missing demographic data that were not included in the normative sample were eligible for inclusion. We report results by diagnostic groups, defined as follows: *Concordant CU* – the same inclusion criteria for the normative sample were applied and only individuals who completed MTD between May 1, 2023 and December 14, 2023 were included; *Discordant CU* – the participant had a consensus diagnosis of CU but at least one rater did not assign a CU diagnosis in their independent determination; this group was included as an exploratory validation sample; *MCI* – per consensus diagnosis; *Dementia* – per consensus diagnosis.

### Mayo Test Drive Measures

MTD platform v1 was used for this study. We created normative data for three primary MTD variables: (1) MTD Composite; (2) SLS Sum of Trials; and (3) Symbols Test accuracy-weighted average correct item response time (referred to as Accuracy-Weighted Symbols or SYM_AW_) (Boots et al., 2024; N. H. Stricker, Stricker, et al., 2024). We also generated normative data for secondary variables to aid clinical interpretation. See **Table 1** for a definition of each variable. Note that SYM_AW_ represents the average response time (RT) on correct items on Symbols across all four trials (inversed), multiplied by the accuracy of performance (see Supplemental Online Materials for specific computation details). Although this makes the primary SYM variable slightly more complicated to interpret based on that score alone, the purpose of this variable is to 1) inverse Symbols so that it can be added to SLS performance for the MTD Composite, and 2) to allow accuracy performance to contribute to interpretation of the SYM primary variable. Users can reference the secondary variables to inform the underlying pattern of performance (e.g., accuracy and various versions of response speed separately). Some research studies may choose a secondary SYM variable as a preferred outcome, with the choice depending on the nature of the population being studied and the goals of the research.

**Table 1.** Mayo Test Drive (MTD) Measure Descriptions, Variable Names, and Observed Characteristics in Normative Sample.

| Measure | Abbreviated Variable Name <sup>a</sup> | Normative Sample Mean (SD), Range | Formula / Description |
| --- | --- | --- | --- |
| **Primary Variables** |  |  |  |
| MTD Composite <sup>b</sup> | mtdrawcompositev1 | 109.1 (19.7), 33-148 | SLS Sum of Trials + Accuracy-Weighted SYM. This raw score composite functions like a total score. |
| SLS Sum of Trials | slssumoftrials | 77.4 (16.8), 20-108 | SLS Trials 1-5 Total Correct + SLS Delay Correct / 108 possible <sup>a</sup> |
| Accuracy-Weighted SYM | symall4corrartaccweighted | 31.7 (6.2), 0-42.7 | Accuracy-Weighted Symbols Average Response Time Correct Items (see supplemental materials for computation details) |
| **Secondary Variables** |  |  |  |
| **Stricker Learning Span (SLS)** |  |  |  |
| SLS Trial 1 Correct | slsr1corr | 6.6 (1.3), 2-8 | SLS Trial 1 Correct / 8 |
| SLS Trial 2 Correct | slsr2corr | 10.0 (2.2), 2-13 | SLS Trial 2 Correct / 13 possible <sup>c</sup> |
| SLS Trial 3 Correct | slsr3corr | 12.9 (3.0), 4-18 | SLS Trial 3 Correct / 18 possible <sup>c</sup> |
| SLS Trial 4 Correct | slsr4corr | 15.7 (4.2), 4-23 | SLS Trial 4 Correct / 23 possible <sup>c</sup> |
| SLS Trial 5 Correct | slsr5corr | 16.6 (4.0), 3-23 | SLS Trial 5 Correct / 23 possible <sup>c</sup> |
| SLS Max Learning Span | slsmaxspan | 17.2 (3.8), 4-23 | Maximum number of words recognized across any of the 5 learning trials / 23 possible <sup>c</sup> |
| SLS Trials 1-5 Total Correct | slstotcorr | 61.9 (12.9), 18-85 | SLS Trial 1 Correct + SLS Trial 2 Correct + SLS Trial 3 Correct + SLS Trial 4 Correct + SLS Trial 5 Correct / 85 possible <sup>c</sup> |
| SLS Delay Correct | slsdelaycorr | 15.4 (4.3), 1-23 | SLS Delay Correct / 23 possible <sup>c</sup> |
| SLS Percent Retention | slsretention | 89.3 (14.3), 17-150 | (SLS Delay Correct / SLS Max Learning Span) * 100 |
| **Symbols Test (SYM)** |  |  |  |
| SYM Total Correct | symsumcorr | 46.6 (2.0), 25-48 | SYM Trial 1 Correct + SYM Trial 2 Correct + SYM Trial 3 Correct + SYM Trial 4 Correct (12 items for each trial, / 48 total) <sup>d</sup> |
| SYM Trial 1 seconds | symr1sec | 50.9 (18.9), 20-248 | Seconds to complete Symbols Trial 1 <sup>e</sup> |
| SYM Trial 2 seconds | symr2sec | 42.2 (13.6), 20-138 | Seconds to complete Symbols Trial 2<sup>e</sup> |
| SYM Trial 3 seconds | symr3sec | 39.9 (12.3), 17-112 | Seconds to complete Symbols Trial 3<sup>e</sup> |
| SYM Trial 4 seconds | symr4sec | 38.9 (11.9), 18-99 | Seconds to complete Symbols Trial 4<sup>e</sup> |
| SYM Middle Trials Avg<br>Completion Time, sec | symmiddle2secavg | 41.7 (12.4), 20-116 | Average seconds to complete a trial when averaged<br>across the “middle” 2 trials, excluding highest and<br>lowest performances<sup>e</sup> |
| SYM Avg Trial Completion<br>Time, sec | symall4secavg | 43.0 (13.0), 20-141 | Seconds to complete each trial (SYM Trial 1 seconds<br>+ SYM Trial 2 seconds + SYM Trial 3 seconds +<br>SYM Trial 4 seconds) / 4 to generate the average<br>seconds to complete a trial<sup>e</sup> |
| SYM Avg Response Time<br>Correct Items, sec | symall4corrartsec | 3.4 (1.1), 1.5-11.5 | Symbols Average Response Time for Correct Items<br>only (correct items across all 4 Symbols trials)<sup>e</sup> |

<sup>a</sup> Abbreviated variable names are used in the Supplementary Online Material and any MTD raw data exports.
<sup>b</sup> Sometimes called MTD Raw Composite to clarify this is not a z-score or standard score composite. See supplemental materials for full computation details.
<sup>c</sup> Actual number presented can vary per computer adaptive testing rules.
<sup>d</sup> Note that this variable has a non-normal distribution with most individuals performing near ceiling. We chose to apply consistent normative methods (T-score derivation) to simplify reporting and score types for users).
<sup>e</sup> Score is inversed as part of the norming process, so a lower normative score indicates lower performance, but on the raw score scale a higher score indicates slower performance.
*Note.* Table used with permission of Mayo Foundation for Medical Education and Research, all rights reserved.

### Normative Methods

We applied normative methods similar to our other recent Mayo Normative Studies publications (Karstens et al., 2023; N. H. Stricker et al., 2021; N. H. Stricker, Christianson, et al., 2024) by using a regression-based normative approach.

### Unadjusted Scaled Score Derivation

Each test score distribution was first normalized (Casaletto et al., 2015; Heaton, Miller, Taylor, & Grant, 2004). Specifically, raw test scores were converted to normalized unadjusted scaled scores (unadj. SS) by calculating their percentile rank based on the cumulative frequency distributions of raw scores and then transformed to follow a normal distribution with mean of 10 and standard deviation of 3 with the use of PROC RANK in Statistical Analysis Software (SAS) Version 9.4. **Figure S1** shows how unadjusted scaled scores vary by level of age, sex and education.

### Model Selection

Quantitative (e.g., *R^2^* = percent variance explained via linear regressions) and visual inspection methods were used to investigate the effects of demographic variables on test performance. Single and multivariable regression models examined effects of *a priori* selected demographic variables of age, age^2^, sex, and education on scaled scores. These *a priori* selections are supported by visualization (**Figures 2 and 3**).

**Figure 2.**
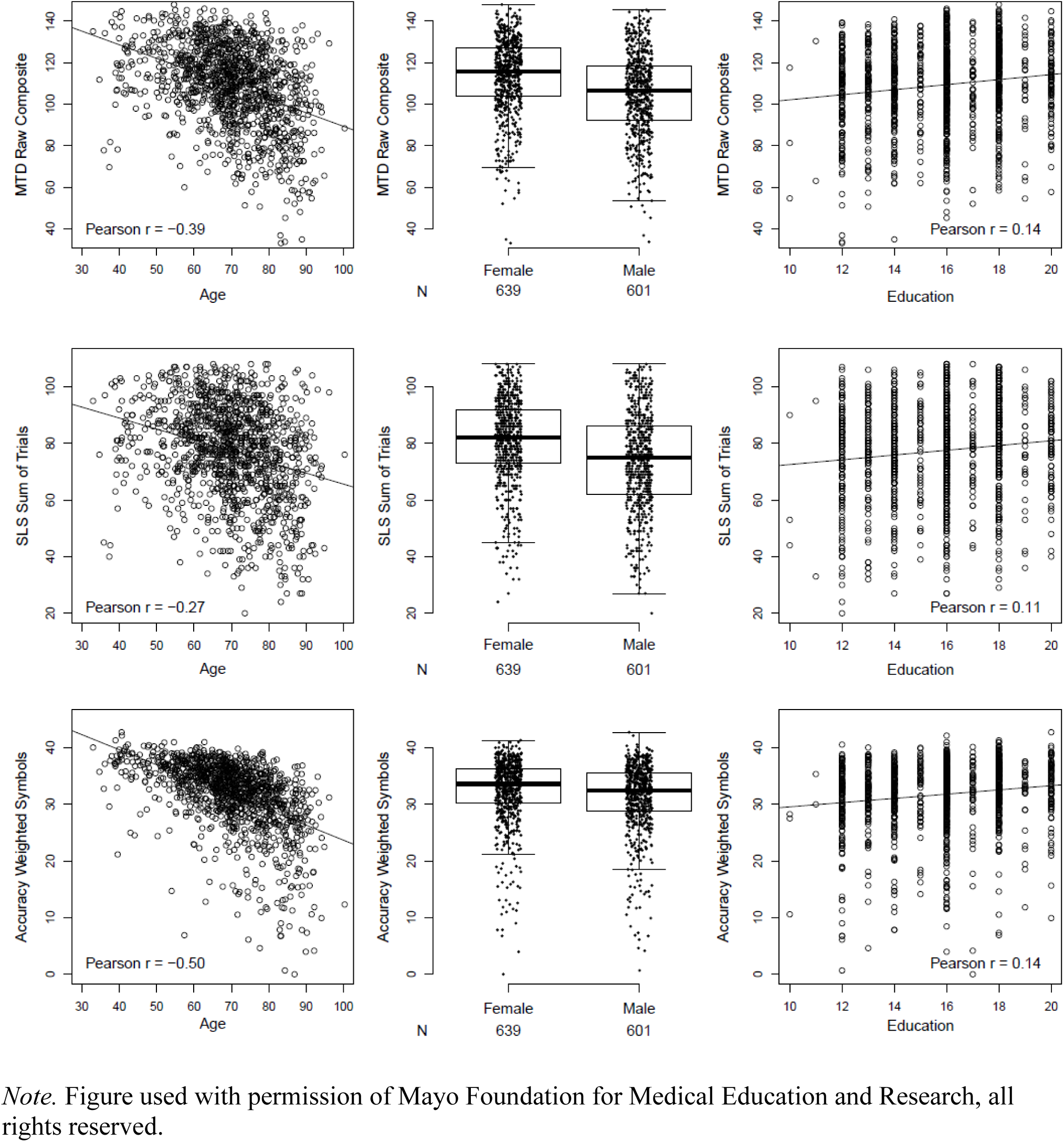
MTD associations with age, sex and education.

**Figure 3.**
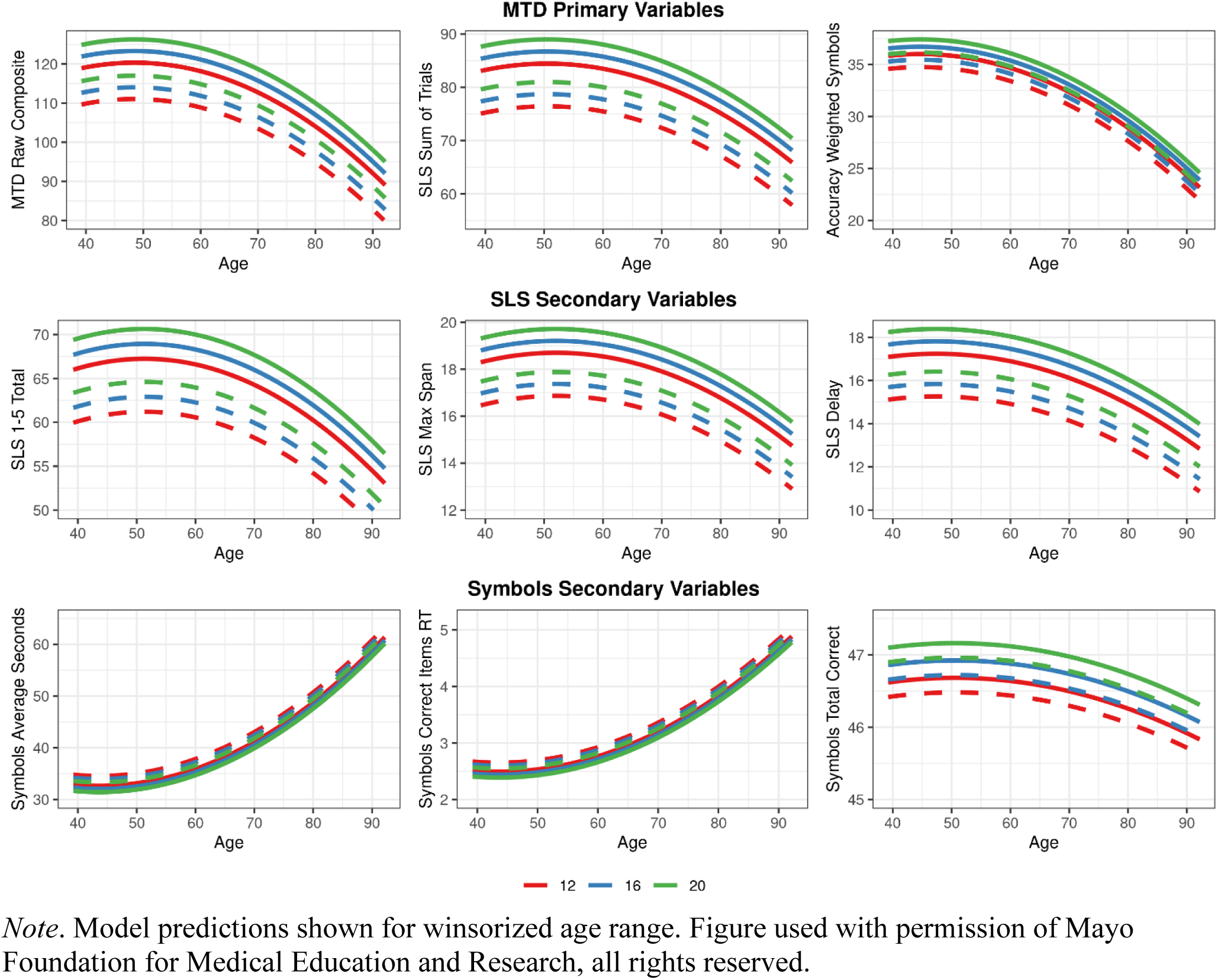
Regression models showing the effect of age, age squared, sex (women, solid lines; men, dashed lines), and years of education (green, 20 years; blue, 16 years; red, 12 years) on raw score performance for the normative sample.

To avoid overfitting the model on statistical significance alone, additional predictors were considered for inclusion if at least 1% incremental variance (adjusted *R*^2^) was explained beyond *a priori* predictors (age, age^2^, sex, education = base model). The unadjusted scaled score was the outcome variable (Y). Additional model terms were added to the base model to determine the need for potential additional variables. Adjusted *R^2^* was used for these models to correct for the number of model terms included. No additional variables resulted in an increment in *R*^2^ >1%, so the base model was retained as the final model; see results for details.

### Fully-Adjusted Normative T-Score Derivation (Age, Age^2^, Sex, and Education)

Next, we generated linear regression models to predict scaled scores from age, age^2^, sex, and education. Residuals for linear regression equations were obtained and converted to T-scores, that is standardized to have a mean of 50 and SD of 10. We examined the need for smoothing to ensure variance of residuals was equal in magnitude across the range of predictors and applied smoothing as necessary. We calculated the predicted mean (SD) age, sex, and education adjusted T-scores for categorized levels of age (30-59, 60-69, 70-79, 80+), sex, or education (<=12, 13-15, 16, 17+). The desired mean and SD was 50 (10) in each category, but a mean within 3 points (Heaton et al., 2004) and SD between 9.4 and 10.6 was considered acceptable (Karstens et al., 2023; N. H. Stricker et al., 2021).

### Age and Sex-Adjusted Normative T-Score Derivation (Age, Age^2^, and Sex)

We also created norms that adjust for age and sex only. While adjustment for education is often recommended, remote assessment can result in situations where educational history is not available.

For example, educational level may be unknown if remote assessment screening is employed pre-visit.

### Age-Adjusted Normative T-Score Derivation (Age and Age^2^)

We also derived norms that only adjust for age. Normative models that adjust for age only can aid comparison to other measures that only adjust for age. The degree of demographic adjustments applied can impact sensitivity to cognitive impairment and further exploration of this is needed. For example, we recently reported an unexpected finding that shows age-adjusted norms provide better sensitivity to MCI/dementia for men; though this increase in sensitivity also leads to reduced specificity (N. H. Stricker, Christianson, et al., 2024).

### Formulas and How to Use the Norms

Automated calculations are built into the Mayo Test Drive platform (v2) and are also available in an excel spreadsheet upon request. First, unadjusted scaled scores are derived from the raw scores using lookup tables. Look up tables for the raw score to scaled score conversions are provided in Tables 2-4. Then, T-scores for a subject’s raw score(s) are calculated with the formulas provided in the Supplemental Online Material. **Table S1** shows the smoothing applied and formulas for all variables. Formulas are provided for age-, sex- and education-adjusted T-scores (ASE), age- and sex-adjusted T-scores (AS), and age-adjusted T-scores (A). Education level determination rules are the same as previously reported (Karstens et al., 2023).

**Table 2.**
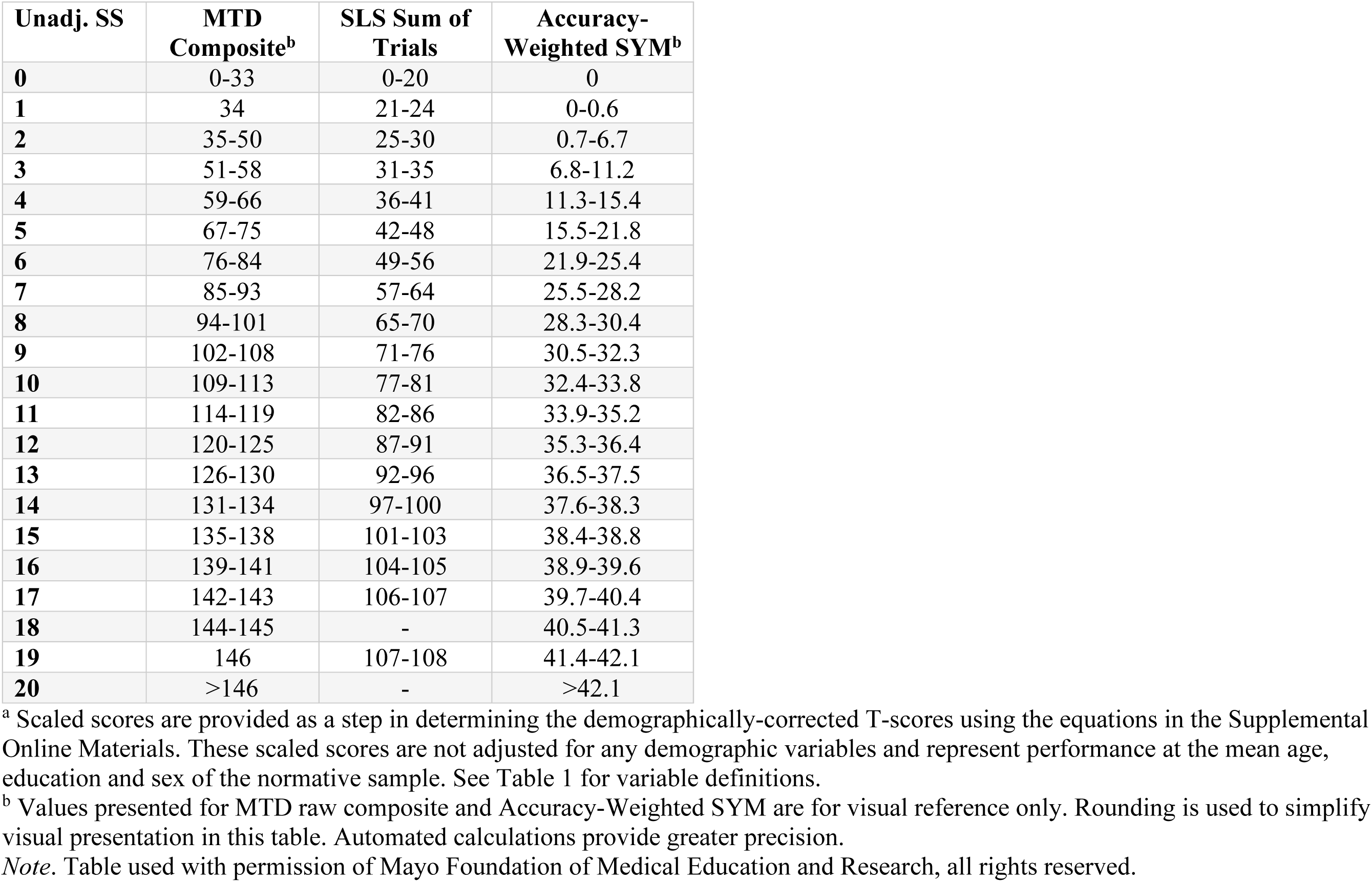
Table for converting raw scores to unadjusted scaled scores for primary variables. ^a^.

**Table 3.** Table for converting raw scores to unadjusted scaled scores for Stricker Learning Span (SLS) secondary variables. ^a^.

| **Unadj.**<br>**SS** | **SLS**<br>**Trial 1** | **SLS**<br>**Trial 2** | **SLS**<br>**Trial 3** | **SLS**<br>**Trial 4** | **SLS**<br>**Trial 5** | **SLS Max**<br>**Span** | **SLS 1-5**<br>**Total** | **SLS**<br>**Delay** | **SLS**<br>**Retention** |
| --- | --- | --- | --- | --- | --- | --- | --- | --- | --- |
| **0** | - | 0-2 | - | - | 0-3 | 0-4 | 0-18 | 0-1 | <16 |
| **1** | 0-2 | 3 | 0-4 | 0-4 | 4 | 5 | 19-22 | 2 | 17-39 |
| **2** | 3 | - | - | - | 5 | 6 | 23-24 | 3 | 40-43 |
| **3** | - | 4 | 5 | 5 | 6 | 7 | 25-29 | 4-5 | 44-49 |
| **4** | 4 | 5 | 6 | 6-7 | 7-8 | 8-9 | 30-34 | 6 | 50-58 |
| **5** | - | 6 | 7 | 8 | 9 | 10 | 35-39 | 7-8 | 59-66 |
| **6** | 5 | 7 | 8-9 | 9-10 | 10-11 | 11-12 | 40-45 | 9-10 | 67-73 |
| **7** | - | 8 | 10 | 11-12 | 12-13 | 13-14 | 46-51 | 11 | 74-78 |
| **8** | 6 | 9 | 11 | 13 | 14 | 15 | 52-57 | 12-13 | 79-83 |
| **9** | - | - | 12 | 14-15 | 15-16 | 16-17 | 58-61 | 14-15 | 84-88 |
| **10** | 7 | 10 | 13 | 16 | 17 | 18 | 62-65 | 16 | 89-93 |
| **11** | - | 11 | 14 | 17-18 | 18-19 | 19 | 66-69 | 17 | 94-95 |
| **12** | - | - | 15 | 19 | 20 | 20 | 70-73 | 18-19 | 96-100 |
| **13** | - | 12 | 16 | 20 | 21 | 21 | 74-76 | 20 | 100-104 |
| **14** | 8 | - | 17 | 21 | 22 | 22 | 77-79 | 21 | 105-106 |
| **15** | - | 13 | - | 22 | - | - | 80-81 | 22 | 107-112 |
| **16** | - | - | 18 | - | 23 | 23 | 82-83 | - | 113-116 |
| **17** | - | - | - | 23 | - | - | 84 | 23 | 117-123 |
| **18** | - | - | - | - | - | - | - | - | 124-127 |
| **19** | - | - | - | - | - | - | 85 | - | 128-133 |
| **20** | - | - | - | - | - | - | - | - | >133 |

<sup>a</sup> Scaled scores are provided as a step in determining the demographically-corrected T-scores using the equations in the Supplemental Online Materials. These scaled scores are not adjusted for any demographic variables and represent performance at the mean age, education and sex of the normative sample. See Table 1 for variable definitions.
*Note.* Table used with permission of Mayo Foundation of Medical Education and Research, all rights reserved.

**Table 4.** Table for converting raw scores to unadjusted scaled scores for Symbols Test (SYM) secondary variables. ^a^.

| **Unadj.**<br>**SS** | **SYM Total**<br>**Correct** | **SYM Trial**<br>**1** | **SYM Trial**<br>**2** | **SYM Trial**<br>**3** | **SYM Trial**<br>**4** | **SYM Middle 2**<br>**Avg Sec** | **SYM All 4**<br>**Avg Sec** | **SYM Avg RT**<br>**Corr Items** |
| --- | --- | --- | --- | --- | --- | --- | --- | --- |
| **0** | 0-31 | >228.6 | >126.4 | >106.8 | >95.2 | >115.4 | >121.2 | >9.9 |
| **1** | 32-34 | 178.4-228.6 | 124.7-126.4 | 104.7-106.8 | 93.1-95.2 | 104.0-115.4 | 113.4-121.2 | 9.0-9.9 |
| **2** | 35-36 | 123.3-178.3 | 99.5-124.6 | 89.3-104.6 | 88.9-93.0 | 91.1-103.9 | 98.3-113.3 | 8.0-8.9 |
| **3** | 37-41 | 102.6-123.2 | 83.4-99.4 | 80.0-89.2 | 76.6-88.8 | 87.8-91.0 | 83.7-98.2 | 6.8-7.9 |
| **4** | 42 | 89.6-102.5 | 71.0-83.3 | 67.5-79.9 | 66.5-76.5 | 69.9-81.7 | 70.8-83.7 | 5.7-6.7 |
| **5** | 43-44 | 79.6-89.5 | 63.0-70.9 | 59.6-67.4 | 57.7-66.4 | 60.4-69.8 | 62.2-70.7 | 5.0-5.6 |
| **6** | 45 | 68.7-79.5 | 55.7-62.9 | 53.3-59.5 | 51.2-57.6 | 55.4-60.3 | 56.8-62.1 | 4.5-4.9 |
| **7** | - | 62.0-68.6 | 50.6-55.6 | 47.2-53.2 | 46.3-51.1 | 49.2-55.3 | 51.0-56.7 | 4.0-4.4 |
| **8** | 46 | 55.0-61.9 | 45.4-50.5 | 42.7-47.1 | 41.6-46.2 | 44.9-49.1 | 46.1-50.9 | 3.6-3.9 |
| **9** | - | 49.6-54.9 | 41.4-45.3 | 39.3-42.6 | 38.2-41.5 | 41.2-44.8 | 42.5-46.0 | 3.3-3.5 |
| **10** | 47 | 44.5-49.5 | 37.8-41.3 | 35.6-39.2 | 34.9-38.1 | 37.6-41.1 | 38.9-42.4 | 3.0-3.2 |
| **11** | - | 40.8-44.4 | 34.7-37.7 | 33.0-35.5 | 32.0-34.8 | 34.4-37.5 | 35.5-38.8 | 2.7-2.9 |
| **12** | - | 36.9-40.7 | 31.9-34.6 | 30.5-32.9 | 29.8-31.9 | 32.0-34.3 | 32.9-35.4 | 2.5-2.6 |
| **13** | 48 | 34.2-36.8 | 29.5-31.8 | 28.6-30.4 | 27.8-29.7 | 30.0-31.9 | 30.6-32.8 | 2.3-2.4 |
| **14** | - | 31.6-34.1 | 27.6-29.5 | 26.6-28.5 | 26.1-27.7 | 28.2-29.9 | 29.2-30.5 | 2.2 |
| **15** | - | 29.6-31.5 | 25.6-27.5 | 24.7-26.5 | 24.8-26.0 | 26.6-28.1 | 27.2-29.1 | 2.1 |
| **16** | - | 27.9-29.5 | 23.9-25.5 | 22.9-24.6 | 22.6-24.7 | 24.6-26.5 | 25.3-27.1 | 1.9-2.0 |
| **17** | - | 24.2-27.8 | 22.2-23.8 | 20.9-22.8 | 20.1-22.5 | 21.9-24.5 | 22.3-25.2 | 1.7-1.8 |
| **18** | - | 22.0-24.1 | 20.2-22.1 | 19.3-20.8 | 19.0-20.0 | 21.1-21.8 | 21.2-22.2 | 1.5-1.6 |
| **19** | - | 20.0-21.9 | 20.1 | 17.0-19.2 | 18.0-18.9 | 19.9-21.0 | 20.2-21.1 | 1.4 |
| **20** | - | <20.0 | <20.1 | <17.0 | <18.0 | <19.9 | <20.2 | <1.4 |

<sup>a</sup> Scaled scores are provided as a step in determining the demographically-corrected T-scores using the equations in the Supplemental Online Materials. These scaled scores are not adjusted for any demographic variables and represent performance at the mean age, education and sex of the normative sample. See Table 1 for variable definitions.
*Note.* Table used with permission of Mayo Foundation of Medical Education and Research, all rights reserved.

### Neuroimaging Methods

A subset of participants had neuroimaging data available, which we include in this manuscript for sample characterization. Specifically, for those with available data, we report mean biomarker levels by group and describe the percentage of the normative and validation samples with positive biomarkers based on the amyloid tau neurodegeneration (AT2N) framework (Jack et al., 2024; Jack et al., 2018) using the imaging data closest to the baseline MTD session within three years. Amyloid and tau positivity was determined using Pittsburgh Compound B PET (PiB-PET) and tau PET (flortaucipir) (Jack et al., 2008; Jack et al., 2017; Vemuri et al., 2017) acquired using a GE Discovery RX or DXT PET/CT scanner. A global cortical PiB PET standard uptake value ratio (SUVR) was computed by calculating the median uptake over voxels in the prefrontal, orbitofrontal, parietal, temporal, anterior cingulate, and posterior cingulate/precuneus regions of interest (ROIs) for each participant and dividing this by the median uptake over voxels in the cerebellar crus gray matter (Jack et al., 2017).

For tau PET we utilized median uptake over the voxels in the meta regions consisting of entorhinal, amygdala, parahippocampal, fusiform, inferior temporal, and middle temporal ROIs normalized to the cerebellar crus gray matter (Jack et al., 2017). Cutoffs used to determine amyloid (A) and tau (T) positivity were SUVR ≥ 1.48 (centiloid 22) (Klunk et al., 2015) and ≥ 1.29 (Lowe et al., 2019), respectively. MRI scans were conducted on a Siemens 3T Prisma scanner using a 3D Magnetization Prepared Rapid Acquisition Gradient-Echo (MPRAGE) sequence. As previously described (Ashburner & Friston, 2005; N. H. Stricker et al., 2022), SPM12 Unified Segmentation was used for tissue-class segmentation with Mayo Clinic Adult Lifespan Template and Advanced Normalization Tools (ANTs) symmetric normalization was used to warp the MCALT-ADIR122 atlas for computing intracranial volume (ICV) and hippocampal volume (Avants et al., 2011; Schwarz et al., 2016). Hippocampal volume was adjusted for ICV as previously described (N. H. Stricker et al., 2022). Values were additionally natural log transformed and z-scored. The cutoff applied to determine neurodegeneration (N) positivity was hippocampal volume ICV adjusted z <= -0.76.

### Additional Statistical Methods

We winsorized age at the 1^st^ and 99^th^ percentile (39.2 and 92.2, respectively). Data were descriptively summarized using counts and percentages for categorical variables and means and standard deviations for continuous variables. Comparison of variable distributions across groups were performed using chi-square tests for categorical variables and ANOVA tests for continuous variables. Pearson correlation coefficients measured the linear relationship between performance on MTD measures and demographic variables. We applied the normative formulas to independent validation samples (CU concordant, CU discordant, MCI, dementia). We calculated the observed proportions of participants performing below a cut-off of -1 SD (T<40, unadj. SS<7). We also calculated 95% confidence intervals (CIs) around those observed proportions. Rates were considered significantly different than expected when 95% CIs did not include the expected 14.7% frequency (since the cumulative distribution function up to -1 SD from the mean is 14.7%) (Karstens et al., 2023; N. H.

Stricker et al., 2021). We also checked these rates in the normative sample itself. Age, sex, and education-adjusted T-scores were hypothesized to have CIs that included the expected 14.7% base rate for the concordant CU participants (normative sample and independent validation sample), and to have CIs that did not include the 14.7% base rate for MCI and dementia participants (independent validation samples). Discordant CU analyses were exploratory. We also completed secondary, exploratory analyses comparing the frequency of low test scores. Chi-square analyses were used to compare the proportion of low test performance across samples (e.g., CU concordant validation sample versus MCI, etc.). To compare use of different normative options that provide varying degrees of demographic adjustment on the same individuals, paired comparisons were completed using a 2x2 table approach to statistically compare agreement across two norms using McNamar *p*-values (performed using the /agree option in PROC FREQ in SAS); we report these analyses only within the MCI group to reduce the number of comparisons and focus on when small differences in sensitivity by normative selection may be most of interest.

Analyses were performed using SAS (SAS Institute Inc., Cary, NC) version 9.4. All tests- were 2-sided, and a p<0.05 was considered significant.

## Results

### Characteristics of the Normative Sample

The normative sample was comprised of 1240 concordant CU adults aged 32-100 years (mean=69.8, SD=11.8) with 51.5% female and 95.5% non-Hispanic White participants, and the mean education was 15.8 years (SD=2.3; **Table 5**). Nearly all participants (99.5%) completed MTD remotely (e.g., unassisted and unsupervised); six participants completed MTD in clinic based on our documentation of in-clinic MTD visits (n=6). Self-report of completing MTD “in a clinic” was slightly higher (n=9), potentially reflecting individuals who work at a clinic or research center or an error in location selection. Most participants reported completing the test session at home (91.9%), and some completed testing at work (6.6%) or in a public space (0.7%). Most participants reported completing MTD on a personal computer (64%), with fewer participants using mobile devices (21% smartphone, 14% tablet). Mouse was the most frequently reported response input source (53%), followed by touch or touchpad/trackpad (44%); a minority used a stylus (2%) or were not sure of the response input source (1%). The frequency of potential interference was 15.9% overall (**Table 5**, see **Table S2** for more granularity).

**Table 5.**
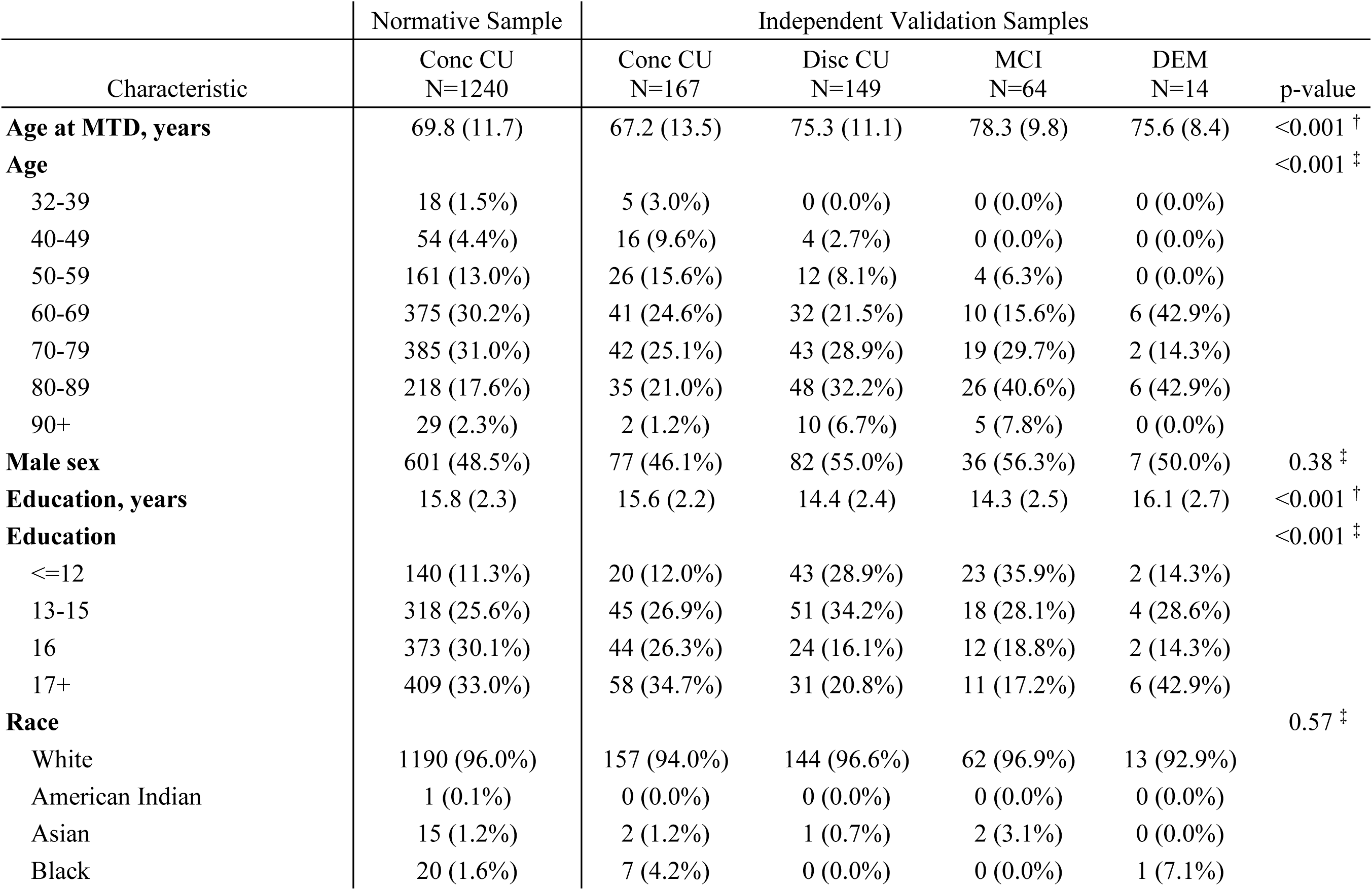
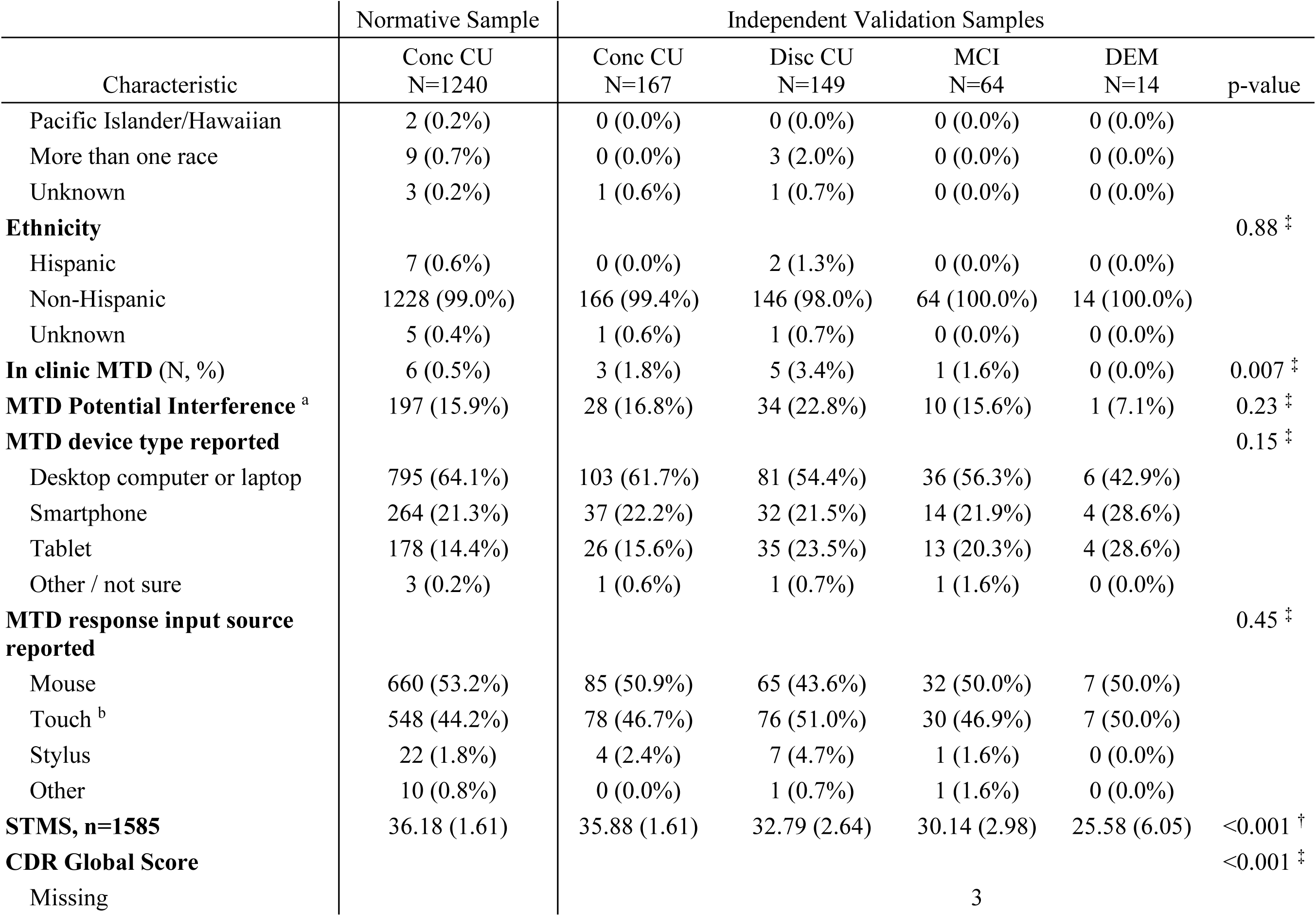
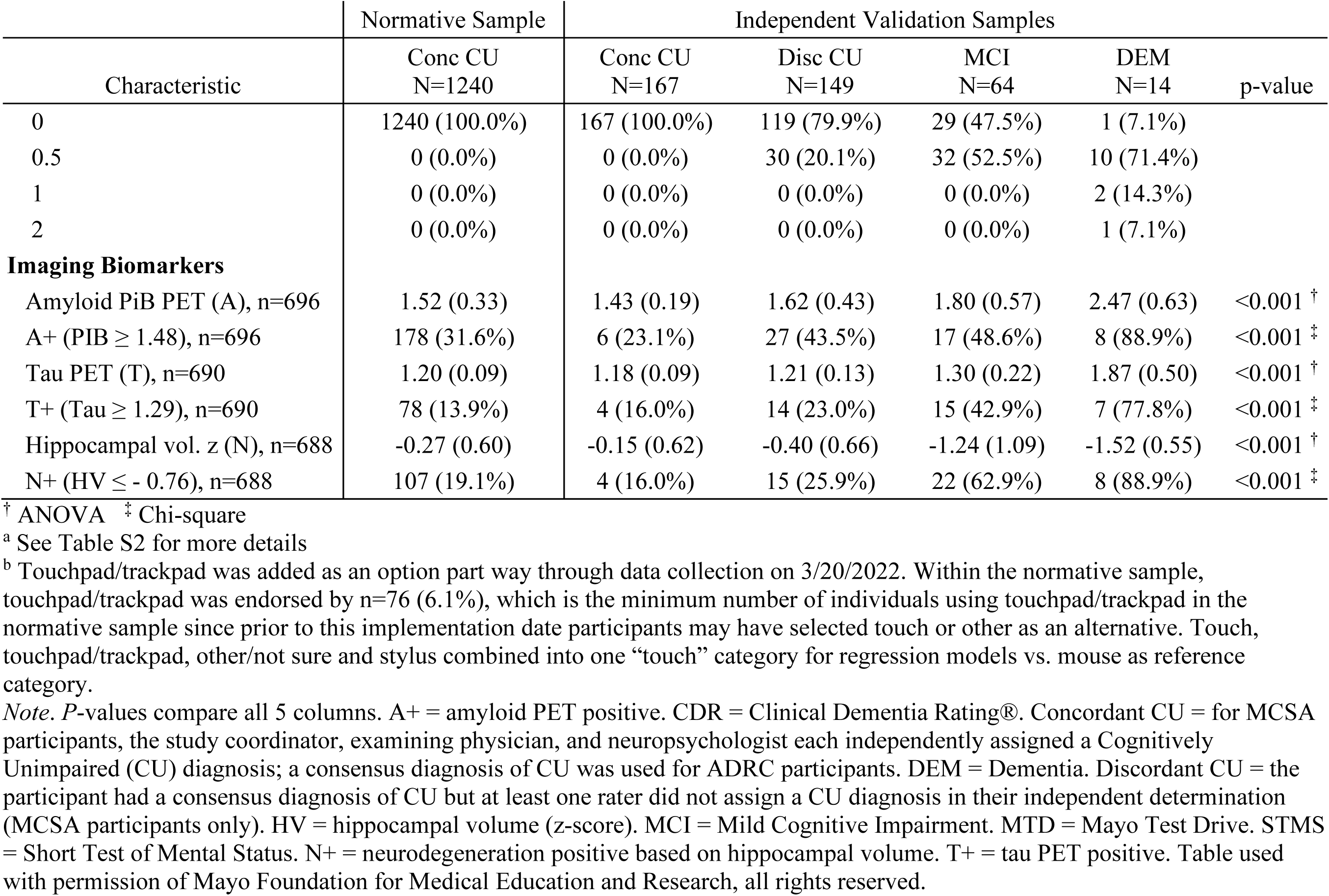
Demographic and other characteristics of the normative sample and the independent validation samples; mean (SD) or count (percent).

### Associations of demographics and other variables of interest with MTD performance

Pearson correlations between test performance (raw scores) and age, sex, and education were significant for all MTD primary variables (**Figure 2**) and nearly all secondary MTD variables (**Table S3**). Increments in adjusted R-squared for scaled score measures are presented in **Table 6**. Age associations were strongest for Symbols, sex effects were more prominent for the SLS relative to Symbols, and education effects were present for all variables but relatively small in magnitude relative to age and sex. Line plots showing model-predicted raw scores for age, age^2^, sex, and education (12, 16 and 20 years) are provided in **Figure 3** to illustrate effects of demographic predictors.

**Table 6.**
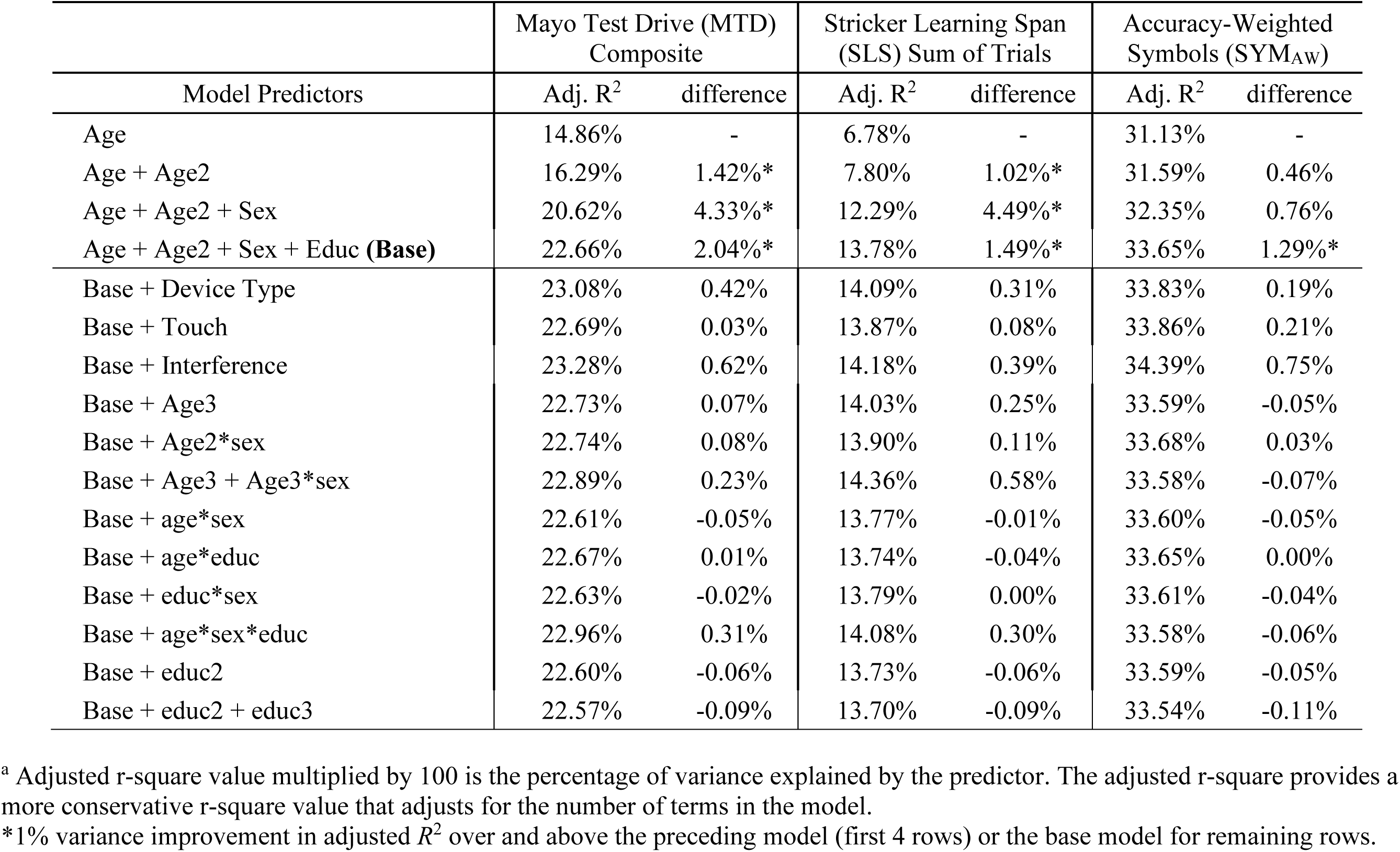
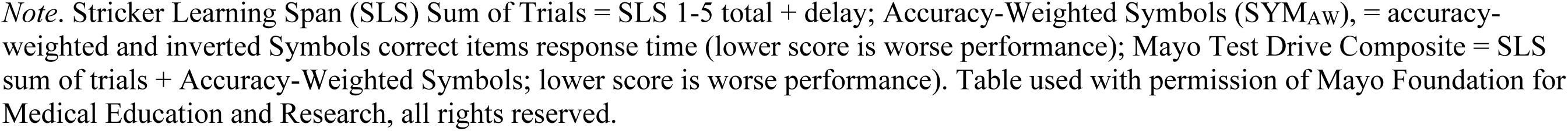
Adjusted r-squared ^a^ where the unadjusted scaled score is the outcome and the predictors included in each separate linear regression model is listed. The difference represents the incremental percentage variance explained (adjusted *R*^2^*100) relative to the preceding model (first four rows) or relative to the base model (Age + Age2 + Sex + Educ).

| Model Predictors | Mayo Test Drive (MTD) Composite |  | Stricker Learning Span (SLS) Sum of Trials |  | Accuracy-Weighted Symbols (SYM<sub>AW</sub>) |  |
| --- | --- | --- | --- | --- | --- | --- |
| | Adj. $R^2$ | difference | Adj. $R^2$ | difference | Adj. $R^2$ | difference |
| Age | 14.86% | - | 6.78% | - | 31.13% | - |
| Age + Age2 | 16.29% | 1.42%* | 7.80% | 1.02%* | 31.59% | 0.46% |
| Age + Age2 + Sex | 20.62% | 4.33%* | 12.29% | 4.49%* | 32.35% | 0.76% |
| Age + Age2 + Sex + Educ (**Base**) | 22.66% | 2.04%* | 13.78% | 1.49%* | 33.65% | 1.29%* |
| Base + Device Type | 23.08% | 0.42% | 14.09% | 0.31% | 33.83% | 0.19% |
| Base + Touch | 22.69% | 0.03% | 13.87% | 0.08% | 33.86% | 0.21% |
| Base + Interference | 23.28% | 0.62% | 14.18% | 0.39% | 34.39% | 0.75% |
| Base + Age3 | 22.73% | 0.07% | 14.03% | 0.25% | 33.59% | -0.05% |
| Base + Age2*sex | 22.74% | 0.08% | 13.90% | 0.11% | 33.68% | 0.03% |
| Base + Age3 + Age3*sex | 22.89% | 0.23% | 14.36% | 0.58% | 33.58% | -0.07% |
| Base + age*sex | 22.61% | -0.05% | 13.77% | -0.01% | 33.60% | -0.05% |
| Base + age*educ | 22.67% | 0.01% | 13.74% | -0.04% | 33.65% | 0.00% |
| Base + educ*sex | 22.63% | -0.02% | 13.79% | 0.00% | 33.61% | -0.04% |
| Base + age*sex*educ | 22.96% | 0.31% | 14.08% | 0.30% | 33.58% | -0.06% |
| Base + educ2 | 22.60% | -0.06% | 13.73% | -0.06% | 33.59% | -0.05% |
| Base + educ2 + educ3 | 22.57% | -0.09% | 13.70% | -0.09% | 33.54% | -0.11% |

<sup>a</sup> Adjusted r-square value multiplied by 100 is the percentage of variance explained by the predictor. The adjusted r-square provides a more conservative r-square value that adjusts for the number of terms in the model.
\*1% variance improvement in adjusted $R^2$ over and above the preceding model (first 4 rows) or the base model for remaining rows.
*Note.* Stricker Learning Span (SLS) Sum of Trials = SLS 1-5 total + delay; Accuracy-Weighted Symbols (SYM<sub>AW</sub>), = accuracy-weighted and inverted Symbols correct items response time (lower score is worse performance); Mayo Test Drive Composite = SLS sum of trials + Accuracy-Weighted Symbols; lower score is worse performance). Table used with permission of Mayo Foundation for Medical Education and Research, all rights reserved.

### Normative Model Selection Results

For primary MTD variables, no additional predictors met the criterion of at least 1% incremental variance explained in the model beyond *a priori* predictors (**Table 6**). Although device type, response input type, and potential interference did not meet the *a priori* criteria, some of these variables were significantly associated with the primary outcome variables, even after adjusting for age, sex and education. These variables are likely important to consider for individual-level interpretation and thus are briefly summarized for primary variables (**Table S4**). Test performance was lower when potential interference was reported during the session for all three primary variables. Device type was significant for MTD composite and SLS Sum of Trials, with those completing a session on a tablet performing worse than those completing a session on a personal computer.

Response input source type was not significantly associated with SLS performance or the MTD Composite but was significantly associated with Accuracy-Weighted SYM performance, with lower performance for touch relative to mouse.

### Normative Sample Model Checks

**Figure 4** illustrates that derived fully adjusted T-scores correct for age effects, with T-scores showing a mean of approximately 50 across all ages. Mean (SD) of age/sex/education adjusted t-scores were similarly within the desired range of mean=47-53 and SD=9.4-10.6 for age categories, sex, and education categories for all outcomes (**Figure 5, Table S5**). There were some combinations of these age/sex/educ categories that did not fall in the desired ranges (e.g. men aged 80+ with 10-12 years of education for MTD raw composite, **Figure S2**), but these combinations had very small sample sizes (**Table S6**). Within the normative sample, the confidence limit of proportion with age/sex/education adjusted T-score contained 14.7% for all variables (**Table S7**). Fully adjusted T-scores removed relationships to demographic variables (all Pearson correlation *p*’s > 0.79), as expected. See Supplemental Online materials for additional model checking methods.

**Figure 4.**
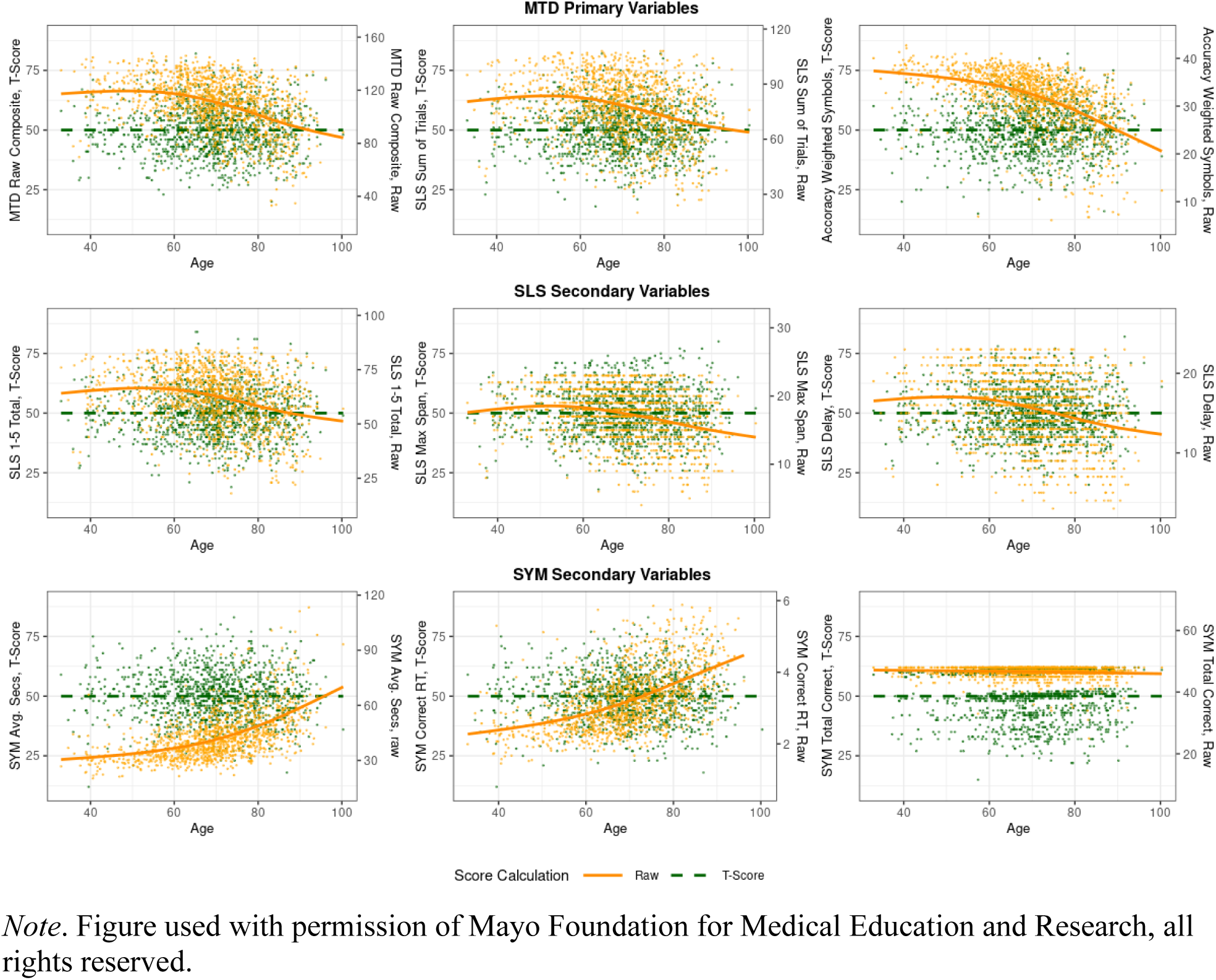
Raw MTD scores (orange, right side of Y-axis) and demographically corrected (for age/age^2^/sex/education) MTD T-scores (green, left side of Y-axis) demonstrating that fully-corrected MNS T-scores correct for age effects.

**Figure 5.**
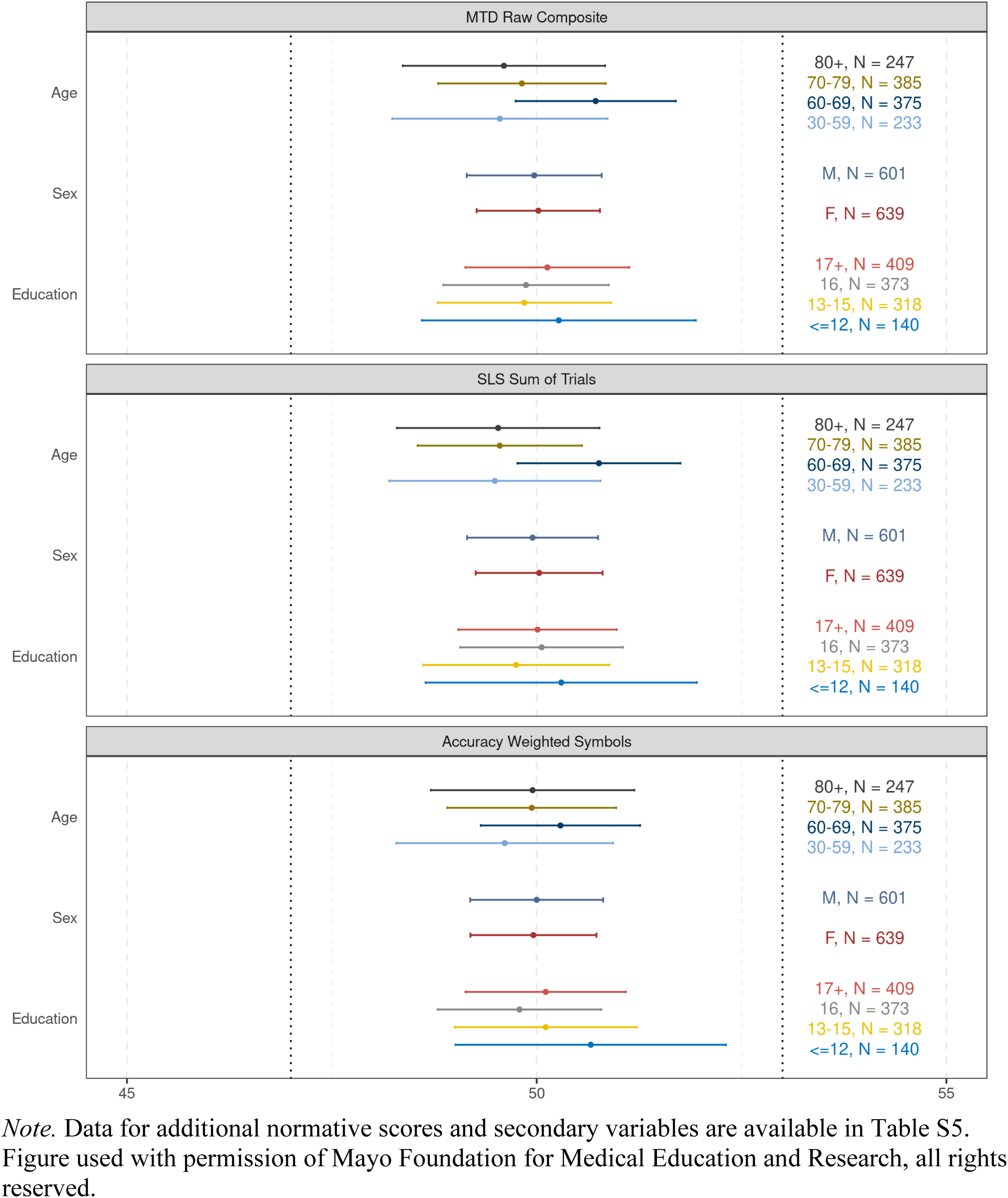
Fully-adjusted T-score statistics for normative sample by categorized age, sex, or education show that mean and SD values for the T-score statistics are within expected ranges (mean 50 +/-3; SD 10 +/- .6).

### Independent validation sample results

The independent validation samples were comprised of 167 concordant CU, 149 discordant CU, 64 MCI, and 14 dementia participants. When comparing these 4 validation samples and the normative sample, there were no significant differences in sex, race/ethnicity, rates of interference reported, device type, or response input type (**Table 5**). There were differences between groups in age and education. Those with some degree of cognitive impairment (discordant CU, MCI, dementia) were older than concordant CU participants, and discordant CU and MCI participants had lower education on average.

Both the fully-adjusted T-scores and the age- and sex-adjusted T-scores showed expected rates of low test performance in the concordant CU validation sample (95% CI includes 14.7%) for all primary variables (**Table S7**). Application of the age-adjusted T-scores showed slightly higher than expected base rates of low test performance for SLS sum of trials (21.0%), but not for the other two primary variables (**Figure 6** and **Table S7**).

**Figure 6.**
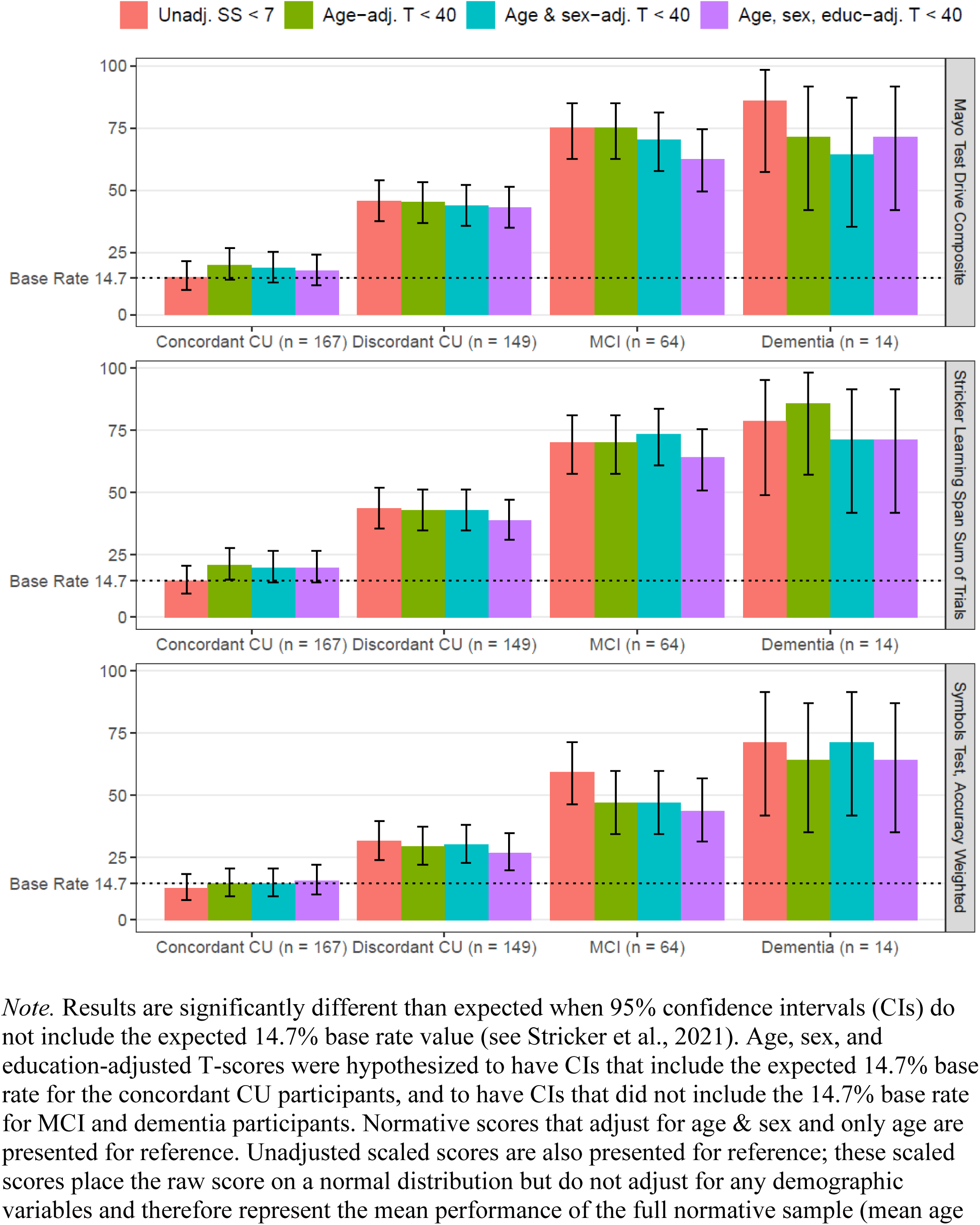
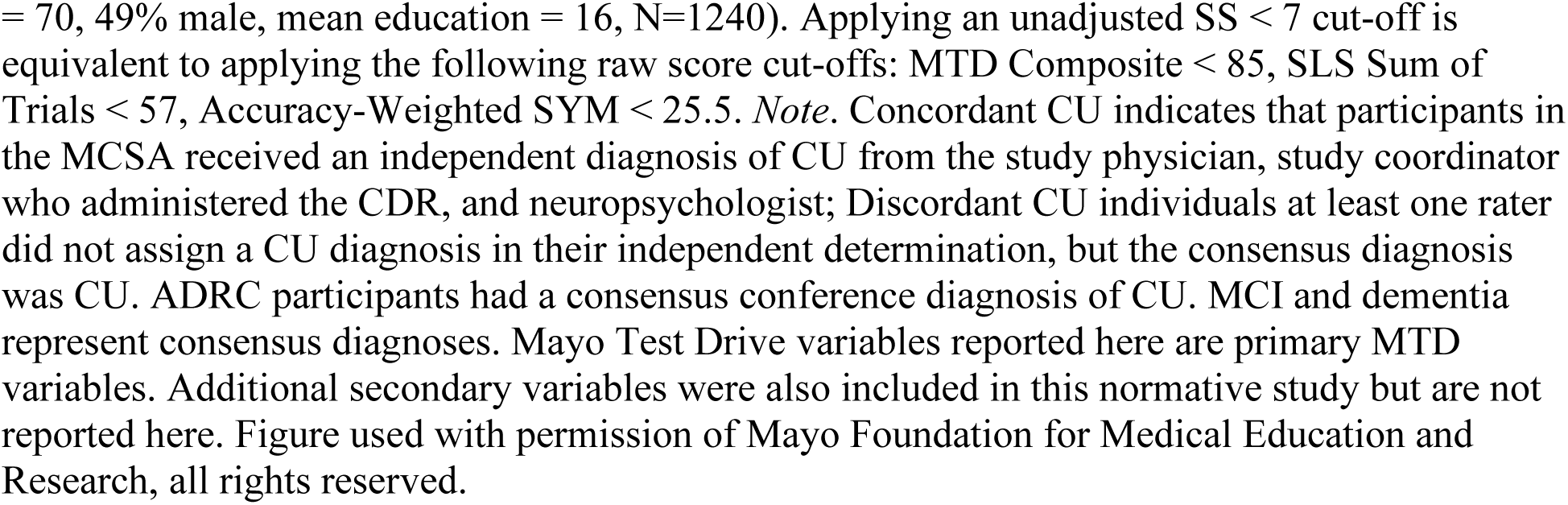
Proportion of individuals in the independent validation samples with normative scores < -1 standard deviation (SD; T < 40; unadj. SS < 7).

Application of norms showed sensitivity to cognitive impairment, with the discordant CU, MCI, and dementia validation samples all showing significantly greater levels of low test performance than typical base rates for all primary variables (**Figure 6** and **Table S7**). For example, low test performance frequencies for the MTD composite were 43.0% in discordant CU, 62.5% in MCI and 71.4% in dementia for the fully-adjusted T-scores (**Figure 6**). Most secondary variables also showed significantly greater than expected levels of low test performance for the discordant CU, MCI and dementia groups, with the exception of Symbols total correct (i.e., Symbols accuracy; **Table S7**).

Chi-square tests comparing the frequency of low test performance across all groups were significant (all *p*’s < .001 for primary variables; data not shown). Chi-square tests comparing paired groups (**Table S7**) showed a significantly higher rate of low test performance in discordant CU, MCI, and dementia validation samples compared to the concordant CU validation sample for the MTD Composite and SLS Sum of Trials (*p* < .001 for all), and Symbols test (*p* < .01). MCI versus dementia group comparisons generally showed no significant difference in the frequency of low test performance, though this comparison is limited by low power. There was a significantly higher rate of low test performance in the MCI group compared to the discordant CU group for the MTD Composite and SLS Sum of Trials (all *p*’s < .01), as well as the Symbols test (*p*’s < .05). A similar pattern was generally seen for secondary MTD variables.

Exploratory cross-norm comparisons are reported within the MCI group to examine the impact of varying levels of demographic adjustment on sensitivity to MCI (**Table S8**). Relative to fully-adjusted T-scores, age-adjusted T-scores and unadjusted scaled scores showed higher sensitivity than fully-adjusted T-scores (*p*=0.005 and *p*=0.03, respectively) for the MTD composite. For SLS sum of trials, sex-adjusted T-scores showed higher sensitivity than fully-adjusted T-scores (*p*=0.03). For SYM_AW_, unadjusted scaled scores showed higher sensitivity than all T-score options (*p*’s ranged from .01-.03). No other within T-score comparisons for the MCI group were significant.

Exploratory analyses looking at amyloid PET, tau PET and hippocampal volume among the subset of participants with imaging data available (42%) shows that the normative sample and concordant CU validation samples tend to have the lowest rates of neuropathological abnormality, but some disease pathology is present in these groups (**Table 5**). For example, abnormal amyloid PET is seen in 23-32% of concordant CU participants. Slightly higher rates (43.5% and 48.6%) are seen in discordant CU and MCI participants, respectively, and dementia participants show the highest rate of amyloid positivity (89%).

## Discussion

This study provides regression-based normative data for self-administered multi-device compatible neuropsychological measures completed in predominantly unsupervised remote environments (99.5% remote) in a well-characterized normative sample of community-dwelling individuals ranging in age from 32-100 years. We demonstrated best model fit when incorporating age (in linear and quadratic forms), sex, and education as demographics. Factors specific to remote digital assessment (e.g., test interference, device type, etc.) did not explain enough additional variance to be included in normative models for MTD measures. Sensitivity of MTD normative data to discordant diagnosis of CU, MCI, and dementia was also examined, with expected greater levels of low test performance seen in these samples. These normative data are provided at a time when production of publicly available and peer-reviewed norms for remote and unsupervised digital cognitive assessments is still in its infancy. However, use of digital cognitive assessment as part of clinical care is becoming increasingly commonplace. With this normative data provided for MTD neuropsychological measures, it is our aim to make remote cognitive assessment with MTD more accessible, transparent, and interpretable for clinical use.

As expected, age, sex, and education were all significantly associated with test performance. Age explained the greatest percentage variance for the Symbols test. Given that the Symbols test is a measure of processing speed/executive function, this is not surprising, as it is well established that these cognitive domains, especially processing speed, are most impacted in normal aging (Bosnes et al., 2022; Salthouse, 2010). Age also explained a significant percentage of the variance for the SLS; however, the amount of variance explained was relatively modest compared to the Symbols test. We previously showed that age correlations with SLS were qualitatively larger in magnitude than age correlations with the AVLT (N. H. Stricker et al., 2022). We have also demonstrated that differing inclusion criteria can impact the magnitude of age associations (N. H. Stricker, Christianson, et al., 2024). Here, our strict inclusion criteria (concordant diagnosis of CU, CDR=0) and self-selection for participating in remote digital assessment may have led to a normative sample with less potential pathological neurodegeneration, and in turn may have reduced age associations. It is worth noting that while it has been suggested that neurodegeneration (particularly AD-related pathological change) should explain more variance in cognitive performance compared to age (Jutten et al., 2023), age- related variance likely accounts for other factors (e.g., other non-AD pathologies/neurodegeneration, medical factors, and accumulative structural and social determinants of health) that meaningfully contribute to cognitive performance. Sex contributed significant additional variance particularly for the SLS, which was expected given known sex differences wherein women outperform men on verbal memory measures across the lifespan (N. H. Stricker et al., 2021). Additionally, once age and sex were accounted for, education only accounted for 1-2% of additional explained variance. This pattern is also true for in-person traditional neuropsychological test normative data from the Mayo Normative Studies. Education still explains a significant amount of variance, which is in line with findings that those with higher levels of education generally perform better on cognitive measures.

Given the digital, remote and self-administration emphasis of MTD, we examined whether other variables related to this form of assessment may be necessary to include in normative models, such as device type, response input source, and potential for interference/distraction. We chose to retain sessions that included potential for interference within the normative dataset because it is most representative of remote self-administered assessments and is part of the rationale for providing norms collected under the same context as planned use for the digital assessment. We characterized the frequency of endorsed subtest interference (range 2-9%, depending on subtest and subtest duration) as well as the frequency of noise reported in the environment (4%). While we did show a statistically significant detrimental effect of potential interference, at the group level the overall magnitude of this effect was relatively small (e.g., potential interference resulted in about 0.7 lower performance on MTD Composite after controlling for age, sex and education). Most importantly, none of the remote, digital testing variables met the 1% incremental variance threshold for inclusion in normative models (only 0.4% for MTD Composite for example). Visser and colleagues (Visser et al., 2021) also reported the frequency of interruptions and noise in their remote self-administered normative sample, although they did not evaluate whether these factors warranted inclusion in normative data. We are aware of one digital assessment, TestMyBrain, that includes device/operating system type along with age, sex, and education in its normative data, presumably because several TestMyBrain subtests show substantial device difference effects (Passell et al., 2021) although a specific normative data publication is not yet available. While consideration of device and response input source effects is important, there are potential drawbacks to including these types of variables in normative models. For example, it may not be feasible or practical for broad users of the normative data to easily include these variables when applying normative data to individual persons because self-report of device type as well as automated collection of device details (e.g., parsing of user agent data) are both subject to error and to changes over time. Device and response input source options also continually expand and evolve. Together, our findings demonstrate that inclusion of these variables for MTD as part of normative data was not warranted. However, we still strongly encourage consideration of these variables for aiding individual-level test interpretation given their potential impacts on test performance. Further, this finding may not generalize to other digital cognitive assessments as we made specific efforts to limit the potential impact of varying device use in the test development phase (J. L. Stricker et al., 2022).

Normative model performance was validated by applying normative data to four independent validation samples, and we saw expected greater levels of low test performance for all primary MTD variables in the discordant CU, mild cognitive impairment, and dementia groups relative to the concordant CU group. While we did not see significant differences in low test performance between the MCI versus dementia groups, potentially due to limited power given the small sample size and the predominantly very mild severity of our participants with dementia (e.g., 79% had a global CDR of 0.5 or less), we did see greater frequency of low test performance in MCI compared to discordant CU. These results mirror findings seen with AVLT normative data (N. H. Stricker, Christianson, et al., 2024) and are expected given findings that MTD performance differs between CU, MCI, and dementia (Boots et al., 2024) in a similar fashion as other memory measures. Further, these findings continue to highlight the importance of removing individuals with MCI from normative data, which now extends to remote digital normative data. Our exploratory findings with the discordant CU group highlight that even more stringent criteria may be beneficial when developing normative data, as individuals still considered CU, but with dissent among assessors, had lower test performance, and were more likely to have abnormal amyloid, tau, and neurodegeneration on neuroimaging metrics. The importance of a refined normative sample for digital cognitive assessment measures was similarly apparent in our past work applying a < -1 SD cut-off using available Cogstate normative data, wherein we showed 38% sensitivity of the Cogstate Brief Battery Learning/Working Memory composite to MCI (Alden et al., 2021).

Norms are provided that adjust for a varying number of demographic variables, from fully-adjusted regression-based norms (age, age squared, sex and education) to regression-based norms that adjust for age and sex, or only age, to none (unadjusted scaled scores that represent performance based on the entire normative sample without demographic adjustment). While we typically recommend use of the fully-adjusted norms to best understand how an individual is performing relative to peers, our prior work demonstrates that this recommendation may need some flexibility. For the AVLT, for example, we demonstrated that norms that only adjust for age were more sensitive to MCI/dementia than fully-adjusted norms for men (N. H. Stricker, Christianson, et al., 2024). Our current MCI/dementia validation samples for Mayo Test Drive are too small to examine potential sex-specific differences in sensitivity of norms and cut-offs applied; this is an important planned future direction.

Additionally, while normative data remains a gold standard for understanding cognitive performance in the context of multiple factors, raw score cut-offs have their own utility in clinical practice, particularly for screening purposes, and can be complementary to normative data. For example, a < 40 T-score cut-off (< -1 SD) with fully-adjusted norms showed 63% sensitivity to MCI and 71.4% sensitivity to dementia, whereas applying an unadjusted scaled score < 7 (< -1 SD, which is equivalent to using a MTD composite raw score cut-off of < 85) showed 75% sensitivity to MCI and 86% sensitivity to dementia while maintaining 85% specificity in the independent Concordant CU validation sample. Future work with MTD will derive raw score clinical cut-offs to complement normative data in order make MTD more accessible for other practitioners (Neurology, Primary Care) for screening purposes. Indeed, some have argued that because advancing age and low education are themselves causal in the dementia pathway that we should not adjust for age and education for screening measures (Piccininni, Rohmann, Wechsung, Logroscino, & Kurth, 2023).

This study has several strengths. It is among the first normative data studies of a remote, self-administered, digital assessment. As such, provision of normative data will aid in introduction of MTD to clinical settings and allow for greater ease and accessibility for cognitive assessment at a time where early screening for cognitive decline is important, particularly as related to assessing risk for Alzheimer’s disease. Our ability to include and assess the influence of digital and remote-specific variables, such as test interference and device and response input types, extends our understanding of how these factors impact test performance at group levels. Additionally, these normative data benefit from a large, population-based sample with a broad age range; this sample was also created with the ability to exclude individuals with any indication of cognitive impairment, increasing the robustness of this normative data. However, this study is also limited by its sample – given the normative sample predominantly resides in Olmsted County, MN, there is a dearth of racial/ethnic diversity in these normative data. As such, we are actively working to include more individuals from historically underrepresented and underserved communities in our work and future iterations of normative data.

Although education had a modest effect, the sample was well above the national average in educational achievement. For example, based on 2015 U.S. Census data, 49.7% of individuals 65 and older have some college or more, whereas 88.7% of the MTD normative sample (mean age 70) has 13 years of education or more and the average education in this sample was almost a 4 year college degree (Ryan, 2016). This may reflect the general demographic of Olmsted County given the population-based design of the MCSA, or this may be additionally related to the lower rates of internet use reported among older adults with high school education or less compared to college graduates (Pew Research Center, 2017). Additionally, although the normative age range is from 32-100, most (92%) of the individuals were ages 50-89 (see **Table 5**). MTD was developed with a goal of assessing subtle cognitive change as related to Alzheimer’s disease and thus will most likely be used predominantly in this age range; nevertheless, utility for younger adults will benefit from increased sample sizes in the future. Another limitation is that while multiple variables and normative score options may be helpful for clinical practice within neuropsychology (like the numerous process scores provided for many neuropsychological tests, for example), this can complicate straightforward use in primary care or for clinical trial study entry decisions. MTD variable selection and cut-off choice should ideally be validated for each specific context of use.

In addition to potential utility for cognitive screening, self-administered remote neuropsychological tests can be used to complement traditional tele-neuropsychology and in-person neuropsychology evaluations. The guidelines for the practice of telepsychology by APA include both synchronous and asynchronous methods in the definition of telepsychology (Joint Task Force for the Development of Telepsychology Guidelines for, 2013). Asynchronous methods can be a helpful complement to real-time tele-neuropsychology (Singh & Germine, 2021). Self-administered measures can also be facilitated and monitored via interactive telehealth methods. The Inter Organizational Practice Committee (IOPC) posted guidelines during the COVID pandemic on remote assessment that state, “Conceptually, it would seem reasonable to consider computerized or web-based assessments to assist with remote testing, when our patients are interacting via their own computer” (Bilder et al., 2020; "Inter Organizational Practice Committee. Recommendations/guidance for teleneuropsychology (TeleNP) in response to the COVID-19 pandemic," 2020). However, concerns were raised that the necessary normative and validation studies typically were not available or normative data was based on in-clinic settings. The current manuscript and our prior validation studies (Boots et al., 2024; N. H. Stricker, Stricker, et al., 2024; N. H. Stricker et al., 2022) offer support for using MTD (or other remote self-administered cognitive assessments with appropriate supporting data) to complement available tele-neuropsychology evaluations in the manner conceptualized by the IOPC. While asynchronous remote cognitive measures completed in isolation should largely be viewed as cognitive screening, when used as part of a more comprehensive neuropsychological or neurological work-up they can contribute meaningfully to an overall assessment. We advocate for a hybrid neuropsychology approach, as described by Singh & Germine (2021). Further, incorporating self-administered computerized neuropsychological tests alongside traditional neuropsychological tests can help neuropsychologists understand how new tests perform in the context of more familiar measures. The addition of self-administered digital neuropsychological measures may be particularly important for tele-neuropsychology evaluations to provide measures of processing speed/executive function that can be logistically challenging if the examinee is in a home setting. If such measures are completed remotely pre-visit, they can be added without having to give up administration of person-administered tests, thus maximizing appointment time for measures requiring interactive administration.

Additionally, memory measures with auditory-only administration, while often well-suited to telehealth evaluation, may be problematic for use with adults with hearing impairment. The SLS provides a helpful alternative for assessment of verbal memory in individuals with hearing impairment. Pre-visit results may also be helpful for assessment planning. Increased familiarity and expertise with new digital measures through hybrid use will help neuropsychologists be better prepared to help oversee and facilitate their use in other multidisciplinary clinics. See Feenstra et al. (2018) for an excellent review of the myriad factors that should be considered and addressed to facilitate reliable online self-administered neuropsychological test administration. With the involvement of neuropsychologists to help oversee appropriate use, the use of remote neuropsychological tests can help promote equitable access to sensitive cognitive screening measures and can assist with earlier diagnosis and intervention for individuals with cognitive impairment.

In summary, this study generated normative data for remote, multi-device compatible, self-administered digital cognitive assessments administered via the MTD platform. Results showed that variables such as test interference, device type, and other factors specific to digital assessment did not meet inclusion criteria for normative data modeling, but still have relevance for individual test performance interpretation. Further, validation findings demonstrate expected lower test performance in discordant CU, MCI, and dementia groups, providing further evidence of sensitivity to cognitive impairment. Together, this work shows the utility of developing normative data for remote, self-administered neuropsychological tests and their potential for clinical use.

## Data Availability

All data produced in the present study are available upon reasonable request to the authors and with the approval of the associated parent study primary investigators.

## Funding, Disclosure Statement and Acknowledgements

Research reported in this publication was supported by the National Institute on Aging of the National Institutes of Health under Award Numbers R01AG081955, R21 AG073967, P30 AG062677, U01 AG006786, R01 AG041851, R37 AG011378, RF1 AG069052, and R01 AG034676 (the Rochester Epidemiology Project). This work was also supported by the Kevin Merszei Career Development Award in Neurodegenerative Diseases Research IHO Janet Vittone, MD, the GHR Foundation, and the Mayo Foundation for Education and Research. The content is solely the responsibility of the authors and does not necessarily represent the official views of the National Institutes of Health or other sponsors. A Mayo Clinic invention disclosure has been submitted for the Stricker Learning Span and the Mayo Test Drive platform (NHS, JLS). We have no other conflicts of interest to disclose related to this work. The authors wish to thank the participants and staff at the Mayo Clinic Study of Aging and Mayo Alzheimer’s Disease Research Center.

## References

Alden, E. C., Pudumjee, S. B., Lundt, E. S., Albertson, S. M., Machulda, M. M., Kremers, W. K., … Stricker, N. H. (2021). Diagnostic accuracy of the Cogstate Brief Battery for prevalent MCI and prodromal AD (MCI A(+) T(+) ) in a population-based sample. Alzheimers Dement, 17(4), 584–594. doi:10.1002/alz.12219

American Psychiatric Association. (1994). Diagnostic and Statistical Manual of Mental Disorders (DSM-IV) (4th ed.). Washington, D.C.: American Psychiatric Association.

Ashburner, J., & Friston, K. J. (2005). Unified segmentation. Neuroimage, 26(3), 839–851. doi:10.1016/j.neuroimage.2005.02.018

Avants, B. B., Tustison, N. J., Song, G., Cook, P. A., Klein, A., & Gee, J. C. (2011). A reproducible evaluation of ANTs similarity metric performance in brain image registration. Neuroimage, 54(3), 2033–2044. doi:10.1016/j.neuroimage.2010.09.025

Backx, R., Skirrow, C., Dente, P., Barnett, J. H., & Cormack, F. K. (2020). Comparing Web-Based and Lab-Based Cognitive Assessment Using the Cambridge Neuropsychological Test Automated Battery: A Within-Subjects Counterbalanced Study. Journal of Medical Internet Research, 22(8), e16792. doi:10.2196/16792

Bilder, R. M., Postal, K. S., Barisa, M., Aase, D. M., Cullum, C. M., Gillaspy, S. R., … Woodhouse, J. (2020). InterOrganizational practice committee recommendations/guidance for teleneuropsychology (TeleNP) in response to the COVID-19 pandemic. Clinical Neuropsychologist, 34(7-8), 1314–1334. doi:10.1080/13854046.2020.1767214

Boots, E. A., Frank, R. D., Fan, W. Z., Christianson, T. J., Kremers, W. K., Stricker, J. L., … Stricker, N. H. (2024). Continuous Associations Between Remote Self-Administered Cognitive Measures and Imaging Biomarkers of Alzheimer’s Disease. J Prev Alzheimers Dis. doi:10.14283/jpad.2024.99

Bosnes, I., Bosnes, O., Stordal, E., Nordahl, H. M., Myklebust, T. A., & Almkvist, O. (2022). Processing speed and working memory are predicted by components of successful aging: a HUNT study. BMC Psychol, 10(1), 16. doi:10.1186/s40359-022-00718-7

Casaletto, K. B., Umlauf, A., Beaumont, J., Gershon, R., Slotkin, J., Akshoomoff, N., & Heaton, R. K. (2015). Demographically Corrected normative standards for the English Version of the NIH Toolbox Cognition Battery. Journal of the International Neuropsychological Society, 21(5), 378–391. doi:10.1017/S1355617715000351

Cromer, J. A., Harel, B. T., Yu, K., Valadka, J. S., Brunwin, J. W., Crawford, C. D., … Maruff, P. (2015). Comparison of Cognitive Performance on the Cogstate Brief Battery When Taken In-Clinic, In-Group, and Unsupervised. Clinical Neuropsychologist, 29(4), 542–558. doi:10.1080/13854046.2015.1054437

Feenstra, H. E., Vermeulen, I. E., Murre, J. M., & Schagen, S. B. (2018). Online Self-Administered Cognitive Testing Using the Amsterdam Cognition Scan: Establishing Psychometric Properties and Normative Data. Journal of Medical Internet Research, 20(5), e192. doi:10.2196/jmir.9298

Gualtieri, C. T., & Johnson, L. G. (2006). Reliability and validity of a computerized neurocognitive test battery, CNS Vital Signs. Archives of Clinical Neuropsychology, 21(7), 623–643. doi:10.1016/j.acn.2006.05.007

Heaton, R. K., Miller, S. W., Taylor, M. J., & Grant, I. (2004). Revised Comprehensive Norms for an Expanded Halstead–Reitan Battery: Demographically Adjusted Neuropsychological Norms for African American and Caucasian Adults. Odessa, FL: Psychological Assessment Resources.

Hughes, M. A., Frank, R. D., Fan, W. Z., Christianson, T. J., Kremers, W. K., Stricker, J. L., … Stricker, N. H. (2024). Reliability of the remote digital self-administered Stricker Learning Span, Symbols Test and Mayo Test DRIVE Screening Battery Composite (poster presentation). Paper presented at the Alzheimer’s Association International Conference, Philadelphia, PA.

Inter Organizational Practice Committee. Recommendations/guidance for teleneuropsychology (TeleNP) in response to the COVID-19 pandemic. (2020). Retrieved from https://iopc.online/remote-neuropsychological-assessment-models

Jack, C. R., Jr., Andrews, S. J., Beach, T. G., Buracchio, T., Dunn, B., Graf, A., … Carrillo, M. C. (2024). Revised criteria for the diagnosis and staging of Alzheimer’s disease. Nature Medicine, 30(8), 2121–2124. doi:10.1038/s41591-024-02988-7

Jack, C. R., Jr., Bennett, D. A., Blennow, K., Carrillo, M. C., Dunn, B., Haeberlein, S. B., … Contributors. (2018). NIA-AA Research Framework: Toward a biological definition of Alzheimer’s disease. Alzheimers Dement, 14(4), 535–562. doi:10.1016/j.jalz.2018.02.018

Jack, C. R., Jr., Lowe, V. J., Senjem, M. L., Weigand, S. D., Kemp, B. J., Shiung, M. M., … Petersen, R. C. (2008). 11C PiB and structural MRI provide complementary information in imaging of Alzheimer’s disease and amnestic mild cognitive impairment. Brain, 131(Pt 3), 665–680. Retrieved from http://www.ncbi.nlm.nih.gov/pubmed/18263627

Jack, C. R., Jr., Wiste, H. J., Weigand, S. D., Therneau, T. M., Lowe, V. J., Knopman, D. S., … Petersen, R. C. (2017). Defining imaging biomarker cut points for brain aging and Alzheimer’s disease. Alzheimers Dement, 13(3), 205–216. doi:10.1016/j.jalz.2016.08.005

Joint Task Force for the Development of Telepsychology Guidelines for, P. (2013). Guidelines for the practice of telepsychology. American Psychologist, *68*(9), 791-800. doi:10.1037/a0035001

Jutten, R. J., Papp, K. V., Hendrix, S., Ellison, N., Langbaum, J. B., Donohue, M. C., … Sikkes, S. A. M. (2023). Why a clinical trial is as good as its outcome measure: A framework for the selection and use of cognitive outcome measures for clinical trials of Alzheimer’s disease. Alzheimers Dement, 19(2), 708–720. doi:10.1002/alz.12773

Karstens, A. J., Christianson, T. J., Lundt, E. S., Machulda, M. M., Mielke, M. M., Fields, J. A., … Stricker, N. H. (2023). Mayo normative studies: regression-based normative data for ages 30- 91 years with a focus on the Boston Naming Test, Trail Making Test and Category Fluency. Journal of the International Neuropsychological Society, 1-13. doi:10.1017/S1355617723000760

Klunk, W. E., Koeppe, R. A., Price, J. C., Benzinger, T. L., Devous, M. D., Sr., Jagust, W. J., … Mintun, M. A. (2015). The Centiloid Project: standardizing quantitative amyloid plaque estimation by PET. Alzheimers Dement, 11(1), 1–15 e11-14. doi:10.1016/j.jalz.2014.07.003

Kokmen, E., Smith, G. E., Petersen, R. C., Tangalos, E., & Ivnik, R. C. (1991). The short test of mental status: Correlations with standardized psychometric testing. Archives of Neurology, 48(7), 725–728. doi:10.1001/archneur.1991.00530190071018

Lowe, V. J., Lundt, E. S., Albertson, S. M., Przybelski, S. A., Senjem, M. L., Parisi, J. E., … Murray, M. E. (2019). Neuroimaging correlates with neuropathologic schemes in neurodegenerative disease. Alzheimers Dement, 15(7), 927–939. doi:10.1016/j.jalz.2019.03.016

Madero, E. N., Anderson, J., Bott, N. T., Hall, A., Newton, D., Fuseya, N., … Glenn, J. M. (2021). Environmental Distractions during Unsupervised Remote Digital Cognitive Assessment. J Prev Alzheimers Dis, 8(3), 263–266. doi:10.14283/jpad.2021.9

Morris, J. C. (1993). The Clinical Dementia Rating (CDR): Current version and scoring rules. Neurology, 43(11), 2412–2414. doi:10.1212/WNL.43.11.2412-a

Nicosia, J., Aschenbrenner, A. J., Balota, D., Sliwinski, M., Tahan, M., Adams, S., … J, H. (2022). Unsupervised High-frequency Smartphone-based Cognitive Assessments Are Reliable, Valid, and Feasible in Older Adults at Risk for Alzheimer Disease. PsyArXiv. doi:10.31234/osf.io/wtsyn.

Passell, E., Strong, R. W., Rutter, L. A., Kim, H., Scheuer, L., Martini, P., … Germine, L. (2021). Cognitive test scores vary with choice of personal digital device. Behavior Research Methods, 53(6), 2544–2557. doi:10.3758/s13428-021-01597-3

Patel, J. S., Christianson, T. J., Monahan, L. T., Frank, F. D., Fan, W. Z., Stricker, J. L., … Stricker, N. H. (2024). *Usability of the Mayo Test Drive remote self-administered web-based cognitive screening battery in adults ages 35 to 100 with and without cognitive impairme*nt. medRxiv [Preprint] 2024.05.28.24307909.

Petersen, R. C. (2004). Mild cognitive impairment as a diagnostic entity. Journal of Internal Medicine, 256(3), 183–194. doi:10.1111/j.1365-2796.2004.01388.x.

Pew Research Center. (2017). Tech Adoption Climbs Among Older Adults. Retrieved from https://www.pewresearch.org/internet/2017/05/17/tech-adoption-climbs-among-older-adults/

Piccininni, M., Rohmann, J. L., Wechsung, M., Logroscino, G., & Kurth, T. (2023). Should Cognitive Screening Tests Be Corrected for Age and Education? Insights From a Causal Perspective. American Journal of Epidemiology, 192(1), 93–101. doi:10.1093/aje/kwac159

Roberts, R. O., Geda, Y. E., Knopman, D. S., Cha, R. H., Pankratz, V. S., Boeve, B. F., … Rocca, W. A. (2008). The Mayo Clinic Study of Aging: Design and sampling, participation, baseline measures and sample characteristics. Neuroepidemiology, 30(1), 58–69. doi:10.1159/000115751

Ryan, C. L., Bauman, K. (2016). Educational Attainment in the United States: 2015. Retrieved from https://www.census.gov/content/dam/Census/library/publications/2016/demo/p20-578.pdf

Salthouse, T. (2010). Selective review of cognitive aging. Journal of the International Neuropsychological Society, 16, 754–760. doi:10.1017/S1355617710000706

Schwarz, C. G., Gunter, J. L., Wiste, H. J., Przybelski, S. A., Weigand, S. D., Ward, C. P., … Jack, C. R. (2016). A large-scale comparison of cortical thickness and volume methods for measuring Alzheimer’s disease severity. NeuroImage: Clinical, 11, 802–812. doi:10.1016/j.nicl.2016.05.017

Singh, S., & Germine, L. (2021). Technology meets tradition: a hybrid model for implementing digital tools in neuropsychology. International Review of Psychiatry, 33(4), 382–393. doi:10.1080/09540261.2020.1835839

Singh, S., Strong, R. W., Jung, L., Li, F. H., Grinspoon, L., Scheuer, L. S., … Germine, L. (2021). The TestMyBrain Digital Neuropsychology Toolkit: Development and Psychometric Characteristics. Journal of Clinical and Experimental Neuropsychology, 43(8), 786–795. doi:10.1080/13803395.2021.2002269

St Sauver, J. L., Grossardt, B. R., Yawn, B. P., Melton, L. J., 3rd, Pankratz, J. J., Brue, S. M., & Rocca, W. A. (2012). Data resource profile: the Rochester Epidemiology Project (REP) medical records-linkage system. International Journal of Epidemiology, *41*(6), 1614-1624. doi:10.1093/ije/dys195

St. Sauver, J. L., Grossardt, B. R., Yawn, B. P., Melton, L. J. r., & Rocca, W. A. (2011). Use of a medical records linkage system to enumerate a dynamic population over time: The Rochester Epidemiology Project. American Journal of Epidemiology, 173(9), 1059–1068. doi:10.1093/aje/kwq482

Stiver, J., Zimmerman, M. E., Cham, H., Nosheny, R. L., Weiner, M. W., Maruff, P., … Mindt, M. R. (2024). Robust demographically-adjusted norms for remote cognitive assessment: Examining the unsupervised Cogstate Brief Battery in an ethnoculturally-diverse sample. Paper presented at the International Neuropsychological Society Conference, New York, NY.

Stricker, J. L., Corriveau-Lecavalier, N., Wiepert, D. A., Botha, H., Jones, D. T., & Stricker, N. H. (2022). Neural network process simulations support a distributed memory system and aid design of a novel computer adaptive digital memory test for preclinical and prodromal Alzheimer’s disease. Neuropsychology. doi:10.1037/neu0000847

Stricker, N. H., Christianson, T. J., Lundt, E. S., Alden, E. C., Machulda, M. M., Fields, J. A., … Petersen, R. C. (2021). Mayo Normative Studies: Regression-Based Normative Data for the Auditory Verbal Learning Test for Ages 30-91 Years and the Importance of Adjusting for Sex. Journal of the International Neuropsychological Society, 27(3), 211–226. doi:10.1017/S1355617720000752

Stricker, N. H., Christianson, T. J., Pudumjee, S. B., Polsinelli, A. J., Lundt, E. S., Frank, R. D., … Petersen, R. C. (2024). Mayo Normative Studies: Amyloid and Neurodegeneration Negative Normative Data for the Auditory Verbal Learning Test and Sex-Specific Sensitivity to Mild Cognitive Impairment/Dementia. Journal of Alzheimer’s Disease, 100(3), 879–897. doi:10.3233/JAD-240081

Stricker, N. H., Lundt, E. S., Alden, E. C., Albertson, S. M., Machulda, M. M., Kremers, W. K., … Mielke, M. M. (2020). Longitudinal Comparison of in Clinic and at Home Administration of the Cogstate Brief Battery and Demonstrated Practice Effects in the Mayo Clinic Study of Aging. The Journal of Prevention of Alzheimer’s Disease, 7(1), 21–28. doi:10.14283/jpad.2019.35

Stricker, N. H., Lundt, E. S., Edwards, K. K., Machulda, M. M., Kremers, W. K., Roberts, R. O., … Mielke, M. M. (2019). Comparison of PC and iPad administrations of the Cogstate Brief Battery in the Mayo Clinic Study of Aging: Assessing cross-modality equivalence of computerized neuropsychological tests. The Clinical Neuropsychologist, 33(6), 1102–1126. doi:10.1080/13854046.2018.1519085

Stricker, N. H., Stricker, J. L., Frank, R. D., Fan, W. Z., Christianson, T. J., Patel, J. S., … Petersen, R. C. (2024). Stricker Learning Span criterion validity: a remote self-administered multi-device compatible digital word list memory measure shows similar ability to differentiate amyloid and tau PET-defined biomarker groups as in-person Auditory Verbal Learning Test. Journal of the International Neuropsychological Society, 30(2), 138–151. doi:10.1017/S1355617723000322

Stricker, N. H., Stricker, J. L., Karstens, A. J., Geske, J. R., Fields, J. A., Hassenstab, J., … Mielke, M. M. (2022). A novel computer adaptive word list memory test optimized for remote assessment: Psychometric properties and associations with neurodegenerative biomarkers in older women without dementia. Alzheimers Dement (Amst), 14(1), e12299. doi:10.1002/dad2.12299

Tsoy, E., Possin, K. L., Thompson, N., Patel, K., Garrigues, S. K., Maravilla, I., … Ritchie, C. S. (2020). Self-Administered Cognitive Testing by Older Adults At-Risk for Cognitive Decline. The Journal of Prevention of Alzheimer’s Disease, 7(4), 283–287. doi:10.14283/jpad.2020.25

Vemuri, P., Lowe, V. J., Knopman, D. S., Senjem, M. L., Kemp, B. J., Schwarz, C. G., … Jack, C. R., Jr. (2017). Tau-PET uptake: Regional variation in average SUVR and impact of amyloid deposition. Alzheimers Dement (Amst), 6, 21–30. doi:10.1016/j.dadm.2016.12.010

Visser, L. N. C., Dubbelman, M. A., Verrijp, M., Wanders, L., Pelt, S., Zwan, M. D., … van der Flier, W. M. (2021). The Cognitive Online Self-Test Amsterdam (COST-A): Establishing norm scores in a community-dwelling population. Alzheimers Dement (Amst), 13(1), e12234. doi:10.1002/dad2.12234

