## Supplementary Material for "Mayo Normative Studies: regression-based normative data for remote self-administration of the Stricker Learning Span, Symbols Test and Mayo Test Drive Screening Battery Composite and validation in individuals with Mild Cognitive Impairment and dementia"

**This is supplementary material for a preprint. The associated manuscript has not yet peer reviewed by a journal.**

**Copyright 2024 Mayo Foundation for Medical Education and Research. All materials in the Supplementary Material used with permission of Mayo Foundation of Medical Education and Research, all rights reserved.**

**Table S5.** Normative score (unadjusted scaled score and T-score) statistics by categorized age, sex, or education to show that mean and SD values for the T-score statistics are within expected ranges (mean 50 +/-3; SD 10 +/- .6). See manuscript Table 1 for MTD variable (outcome) abbreviation key. T-ASE adj = age-, sex- and education-adjusted T-scores; T-AS adj = age- and sex-adjusted T-scores; T-A adj = age-adjusted T-scores. SS unadj. = unadjusted scaled scores from manuscript Tables 2-4. .......... 19

**Supplemental Methods.**

*Additional model checking methods*
As a potential alternative to uncorrected scaled scores, we reviewed potential use of scaling with more granularity (T-scores with a mean of 50 and standard deviation [SD] of 10) but there was high concordance across both methods, so we retained use of scaled scores for simplicity and consistency with past work.

As described in the manuscript, fully corrected T-scores had a mean of approximately 50 across all age values, education values, and sex when collapsing across levels. However, investigation of mean and SD values across subgroups of levels (e.g., males, ages 80+, 10-12 years of education; n=13) showed that some mean values fell outside the desired range of 47-53 (see Supplemental Figure 2). We thoroughly explored whether additional model terms could help address this (e.g., we attempted adding model terms that explained <1% variance). Ultimately, we identified one model that improved the number of values falling outside the desired ranges at these subgroup levels, with a model that included a three way interaction with age*sex*education (and all two-way interactions as required for modeling) as well as $age^3$ and $age^3$*sex. This model resulted in only one subgroup having a mean value outside the desired range; however, visual evaluation of this model was counterintuitive, and we concluded it likely would not generalize well to other independent samples. We therefore retained our base model as our fully-adjusted model.

For regression models testing whether additional model terms led to incremental percentage variance explained as displayed in manuscript Table 6, the following categorical variables were assigned. Device type = coded 0 (reference) if desktop computer or laptop, 1 if smartphone, 2 if tablet, 3 if other/not sure. Touch = coded 0 (reference) if mouse, 1 if touch or touchpad/trackpad, 2 if stylus/digital pen, 3 if other/not sure. Interference = coded 1 if any potential interference endorsed across any subtest (SLS 1-5, SLS delay, SYM) or noise endorsed in the testing environment (see Table 1).

*Detailed review of how to calculate the Mayo Test Drive Composite and Symbols, accuracy-weighted variables.*
Note that this information is also presented in the supplementary material of Boots et al. (2024, JPAD).

mtdrawcompositev1 = MTD raw composite (MTD-SBCr)
symsumcorr = Symbols total correct across all 4 trials (0-48)
symsumcorr1to5scale = Symbols accuracy weighting to apply to SYM (1-5, as defined below)
symall4corrart = Symbols average correct items response time, seconds, across all 4 trials (SYM), range 0+

symall4corrartaccweighted = Accuracy-weighted Symbols average correct items response time ($SYM_{aw}$)
slssumoftrials = Stricker Learning Span (SLS) sum of trials (range 0-108)

If one of the variables needed to do the calculation is missing, return as missing.

Compute symsumcorr1to5scale as follows:

If symsumcorr >= 47 then symsumcorr1to5scale = 5
If symsumcorr = 46 then symsumcorr1to5scale = 4.75
If symsumcorr = 45 then symsumcorr1to5scale = 4.5
If symsumcorr = 44 then symsumcorr1to5scale = 4.25
If symsumcorr = 43 then symsumcorr1to5scale = 4
If symsumcorr = 42 then symsumcorr1to5scale = 3
If symsumcorr < 42 AND symsumcorr > 34 then symsumcorr1to5scale = 2
If symsumcorr < 35 then symsumcorr1to5scale = 1

Compute tenminussymall4corrartsec: 10 - symall4corrartsec

Compute symall4corrartaccweighted:

- If tenminussymall4corrartsec < 0 then symall4corrartaccweighted = 0
- If tenminussymall4corrartsec > 0 then symall4corrartaccweighted = tenminussymall4corrartsec * symsumcorr1to5scale

Compute mtdrawcompositev1 = slssumoftrials + symall4corrartaccweighted

Range Notes: The range of the symall4corrartaccweighted is 0 – 50, in theory. A score of 50 is essentially impossible to attain, as this would require a 0 symbols average correct items response time (all timed responses must be > 0). The range of the MTD-SBCr is 0 – 158, in theory. Because of inclusion of symall4corrartaccweighted, this ceiling is theoretical and impossible to attain in practice.

### Supplemental Results

*Normative Model Selection Results, Smoothing*

Only one variable met our criteria of a SD within 0.6 *SD* of 10 without application of smoothing (SLS Max Span). Values closer to the ideal (mean 50, SD 10) when collapsing across levels were achieved for this variable when smoothing was added, thus we applied smoothing to all variables.

*Normative Model Selection Results, Secondary MTD variables*

For secondary MTD variables, one variable was just over the 1% cut-off for incremental variance explained beyond the base model. Specifically, SLS Trial 1 had an additional model term meeting these criteria; age*sex*education explained 1.01% variance over and above the base model and we considered adding this as a term to the normative model for this variable. However, addition of this term would have resulted in an overly complicated model, and this was a single, secondary variable (not recommended for use as a primary outcome measure but included for comprehensiveness for clinical use), thus we chose to not add this term to the normative model.

### Supplemental Tables

**Table S1.** T-score formulas and smoothing applied. For each variable, the smoothing applied and the formula for 1) age-, sex- and education-adjusted T-scores (ASE), 2) age- and sex-adjusted T-scores (AS), and 3) age-adjusted T-scores (A) are displayed. SS value in formulas are the unadjusted scaled scores from manuscript Tables 2-4. Male_sex indicates male is coded as 1, female is coded as 0. Educ = education. Age_sq = age-squared. Due to limited representation at the ends of the age distribution in the normative sample, when applying formulas, we windsorized (i.e., capped) age values at the high 99th and low 1st percentile, respectively (39.2 and 92.2). These caps are applied in automated calculations (MTD platform). Thus, although ages 32 to 100 are represented in the normative samples, normative formulas apply these caps to optimize model performances.

**Table S1. T-score formulas**

| Characteristic (see Table 1 for variable abbreviations) | formula |
|---|---|
| **mtdrawcompositev1** | |
| ASE smoother | age+age2+age3 |
| ASE formula | round( 50 + (((ss_mtdrawcompositev1 - ( 4.38697137264937 + age * 0.20665910572595 + age_sq * -0.00223576037823 + male * -1.41752176822815 + educ * 0.19463048824666)) / ( 6.27194045472001 - 0.15397806680306 * age + 0.00169902972549 * age_sq - 0.00000520640068 * age_3) - 0.00028862336951)/ 1.24681335136331)*10 ) |
| AS smoother | age+age2+sex |
| AS formula | round( 50 + (((ss_mtdrawcompositev1 - ( 8.25404213972117 + age * 0.18465737878266 + age_sq * -0.00210347183521 + male * -1.26425614065104 )) / ( 5.12329174730128 - 0.09545779678642 * age + 0.00072082574450 * age_sq + 0.14397851374657 * male) - 0.00033429091404)/ 1.24968407527250)*10 ) |
| A smoother | age+age2+age3 |
| A formula | round( 50 + (((ss_mtdrawcompositev1 - ( 7.82277691622409 + z_age * 0.18023189855613 + age_sq * -0.00207803702362 )) / ( 8.62161900948592 - 0.26747435468179 * z_age + 0.00352948859803 * age_sq - 0.00001472617169 * age_3) - 0.00092168009183)/ 1.24568709751986)*10 ) |
| **slssumoftrials** | |
| ASE smoother | age+age2+e+e2+e3+sex |
| ASE formula | round( 50 + (((ss_slssumoftrials - ( 4.01679185240990 + age * 0.19628383646939 + age_sq * -0.00192410810390 + male * -1.41819207177601 + educ * 0.16754437405037)) / (45.45839738079050 - 0.06017646852403 * age + 0.00048609613629 * age_sq - 8.24313396886358 * educ + 0.53735447357732 * educ_sq - 0.01151723976105 * educ_3 + 0.18471354912023 * male) + 0.00232487893626)/ 1.24759056193174)*10 ) |
| AS smoother | age+age2+sex |
| AS formula | round( 50 + (((ss_slssumoftrials - ( 7.34569451773346 + age * 0.17734402081178 + age_sq * -0.00181022974221 + male * -1.28625594056053 )) / ( 4.02077958663848 - 0.06293314166853 * age + 0.00050515102433 * age_sq + 0.17483638363932 * male) - 0.00029960459560)/ 1.25190974752968)*10 ) |
| A smoother | age+age2 |

**Table S1. T-score formulas**

| Characteristic (see Table 1 for variable abbreviations) | formula |
|---|---|
| A formula | round( 50 + (((ss_slssumoftrials - ( 6.90692468475119 + z_age * 0.17284153112778 + age_sq * -0.00178435232984 )) / ( 3.48979916654954 - 0.04448439127132 * z_age + 0.00038597457127 * age_sq) - 0.00019275416688)/ 1.24416739705583)*10 ) |
| **symall4weighted** | |
| ASE smoother | age+age2+e+e2+e3+sex |
| ASE formula | round( 50 + (((ss_symall4corrartaccweighted - (11.95033118197410 + age * 0.03482627024263 + age_sq * -0.00130091995869 + male * -0.66453623575159 + educ * 0.15546169516553)) / ( 4.81915852782770 - 0.05785484768859 * age + 0.00037803493823 * age_sq + 0.03087543612493 * educ - 0.01243135291866 * educ_sq + 0.00045687531787 * educ_3 + 0.05735518788655 * male) + 0.00012270754480)/ 1.26255476597547)*10 ) |
| AS smoother | age+age2+sex |
| AS formula | round( 50 + (((ss_symall4corrartaccweighted - (15.03916573297290 + age * 0.01725232384817 + age_sq * -0.00119525408212 + male * -0.54211484900796 )) / ( 3.98833344998132 - 0.05571512695760 * age + 0.00036397183685 * age_sq + 0.06183035814122 * male) - 0.00008603181409)/ 1.26577590256641)*10 ) |
| A smoother | age+age2+age3 |
| A formula | round( 50 + (((ss_symalltwo - (14.85423858569550 + z_age * 0.01535467157056 + age_sq * -0.00118434759837 )) / ( 6.69827217642655 - 0.19205879637426 * z_age + 0.00258866813429 * age_sq - 0.00001167992617 * age_3) - 0.00005945153346)/ 1.26876947647917)*10 ) |
| **symall4corrartsec** | |
| ASE smoother | educ+educ2 |
| ASE formula | round( 50 + (((ss_symall4corrartsec - (13.31426123442880 + age * 0.03738204821030 + age_sq * -0.00135123587411 + male * -0.49183596030696 + educ * 0.06829118397130)) / ( 6.40644689705995 - 0.55567438972786 * educ + 0.01690511994600 * educ_sq) + 0.00239491028920)/ 1.26682767812889)*10 ) |
| AS smoother | No smoothing |
| AS formula | round( 50 + (((ss_symall4corrartsec - (14.67112389023970 + age * 0.02966216837078 + age_sq * -0.00130481898788 + male * -0.43805871675362 )) / ( 1 ) - 0.00000000000054)/ 2.46168161626039)*10 ) |
| A smoother | age+age2+age3 |
| A formula | round( 50 + (((ss_symallone - (14.52169254930240 + z_age * 0.02812876060101 + age_sq * -0.00129600594710 )) / ( 9.81379411491229 - 0.35253323262041 * z_age + 0.00512396110834 * age_sq - 0.00002424788258 * age_3) - 0.00001600405387)/ 1.26764839624928)*10 ) |
| **slsr1corr** | |
| ASE smoother | age+age2+educ+educ2 |
| ASE formula | round( 50 + (((ss_slsr1corr - ( 9.30350729647067 + age * 0.06888384576011 + age_sq * -0.00092586890466 + male * -0.60942980347925 + educ * 0.04449765765995)) / ( 7.60309578662162 - 0.06534347752873 * age + 0.00050483559407 * age_sq - 0.38095385316347 * educ + 0.01189812770264 * educ_sq) + 0.00021082543737)/ 1.21218587590322)*10 ) |
| AS smoother | age+age2 |
| AS formula | round( 50 + (((ss_slsr1corr - (10.18762156186710 + age * 0.06385367128812 + age_sq * -0.00089562425824 + male * -0.57438924283328 )) / ( 4.75047245333013 - 0.07062281942349 * age + 0.00055110915008 * age_sq) - 0.00015183743629)/ 1.21369109135093)*10 ) |

**Table S1. T-score formulas**

| Characteristic (see Table 1 for variable abbreviations) | formula |
|---|---|
| A smoother | age3 |
| A formula | round( 50 + (((ss_slsr1corr - ( 9.99168491800219 + z_age * 0.06184304373033 + age_sq * -0.00088406846531 )) / ( 2.43354727998437 + 0.00000040845228 * age_3) - 0.00000391177945)/ 1.21694304885425)*10 ) |
| **slsr2corr** | |
| ASE smoother | age+age2 |
| ASE formula | round( 50 + (((ss_slsr2corr - ( 5.59050643109868 + age * 0.13239827094348 + age_sq * -0.00143859952027 + male * -0.97591423795075 + educ * 0.18219640261568)) / ( 3.91493739709764 - 0.05887510578430 * age + 0.00051707912492 * age_sq) - 0.00017896781897)/ 1.21448860994191)*10 ) |
| AS smoother | age+age2 |
| AS formula | round( 50 + (((ss_slsr2corr - ( 9.21052705976713 + age * 0.11180213753955 + age_sq * -0.00131476231027 + male * -0.83244007703823 )) / ( 4.79827631501535 - 0.08405910276031 * age + 0.00069623082886 * age_sq) - 0.00027892605344)/ 1.21430199338376)*10 ) |
| A smoother | age+age2 |
| A formula | round( 50 + (((ss_slsr2corr - ( 8.92656367265877 + z_age * 0.10888821291442 + age_sq * -0.00129801494726 )) / ( 4.67693590608616 - 0.08039320490417 * z_age + 0.00067318568085 * age_sq) - 0.00029226941923)/ 1.21705075829320)*10 ) |
| **slsr3corr** | |
| ASE smoother | age+age2 |
| ASE formula | round( 50 + (((ss_slsr3corr - ( 5.40831991087844 + age * 0.16107809858438 + age_sq * -0.00159854021794 + male * -1.22193572121273 + educ * 0.12771316355440)) / ( 3.69349738506666 - 0.05787597279940 * age + 0.00051996205224 * age_sq) - 0.00046574180116)/ 1.24057052930495)*10 ) |
| AS smoother | age+age2+sex |
| AS formula | round( 50 + (((ss_slsr3corr - ( 7.94582493283354 + age * 0.14664094551088 + age_sq * -0.00151173476169 + male * -1.12136546452604 )) / ( 3.85626945090666 - 0.06345625694387 * age + 0.00055538506255 * age_sq + 0.12098020704819 * male) - 0.00047962556678)/ 1.24060028347278)*10 ) |
| A smoother | age+age2+age3 |
| A formula | round( 50 + (((ss_slsr3corr - ( 7.56330282190143 + z_age * 0.14271564862648 + age_sq * -0.00148917468198 )) / ( 7.02499876489912 - 0.22018947313895 * z_age + 0.00308247022363 * age_sq - 0.00001295260526 * age_3) - 0.00175070781489)/ 1.23434159006326)*10 ) |
| **slsr4corr** | |
| ASE smoother | age+age2 |
| ASE formula | round( 50 + (((ss_slsr4corr - ( 4.41180298081234 + age * 0.18154421188143 + age_sq * -0.00173145639236 + male * -1.34514278711706 + educ * 0.14223245298385)) / ( 3.52025694484107 - 0.04807724466558 * age + 0.00041580464495 * age_sq) - 0.00011842741410)/ 1.26548384399012)*10 ) |
| AS smoother | age+age2 |
| AS formula | round( 50 + (((ss_slsr4corr - ( 7.23778860114178 + age * 0.16546574636857 + age_sq * -0.00163478230935 + male * -1.23313902879131 )) / ( 3.87076402839835 - 0.05678835979902 * age + 0.00046865201264 * age_sq) - 0.00011721919953)/ 1.26918126601678)*10 ) |
| A smoother | age+age2 |

**Table S1. T-score formulas**

| Characteristic (see Table 1 for variable abbreviations) | formula |
|---|---|
| A formula | round( 50 + (((ss_slsr4corr - ( 6.81713810022289 + z_age * 0.16114919040098 + age_sq * -0.00160997352429 )) / ( 3.24554044296643 - 0.03678954381982 * z_age + 0.00033062180569 * age_sq) - 0.00011749795259)/ 1.25483605419434)*10 ) |
| **slsr5corr** | |
| ASE smoother | age+sex+educ |
| ASE formula | round( 50 + (((ss_slsr5corr - ( 2.09044294738784 + age * 0.23349505619824 + age_sq * -0.00213277145435 + male * -1.31528769777689 + educ * 0.18275034904326)) / ( 2.12405264003883 + 0.00432980770128 * age - 0.02400717860730 * educ + 0.17160941632396 * male) + 0.00013117276797)/ 1.26539347202616)*10 ) |
| AS smoother | age+age2+sex |
| AS formula | round( 50 + (((ss_slsr5corr - ( 5.72146981693825 + age * 0.21283630270765 + age_sq * -0.00200855773208 + male * -1.17137732079344 )) / ( 3.55245188915537 - 0.05135162383286 * age + 0.00041585101773 * age_sq + 0.20758024047207 * male) - 0.00025956393104)/ 1.26862965597795)*10 ) |
| A smoother | age+age2 |
| A formula | round( 50 + (((ss_slsr5corr - ( 5.32188757580218 + z_age * 0.20873594123476 + age_sq * -0.00198499149380 )) / ( 2.92389089963768 - 0.02805712403136 * z_age + 0.00025320198312 * age_sq) - 0.00009965008902)/ 1.25036920752539)*10 ) |
| **slsmaxspan** | |
| ASE smoother | age+sex+educ |
| ASE formula | round( 50 + (((ss_slsmaxspan - ( 3.26310533798604 + age * 0.20554352226043 + age_sq * -0.00195660709619 + male * -1.38749498712351 + educ * 0.17653231187893)) / ( 2.02414845208242 + 0.00370281650983 * age - 0.00336988578369 * educ + 0.13750316446260 * male) + 0.00002091720474)/ 1.23903880484813)*10 ) |
| AS smoother | age+age2+sex |
| AS formula | round( 50 + (((ss_slsmaxspan - ( 6.77058738183435 + age * 0.18558767794427 + age_sq * -0.00183661971628 + male * -1.24848112649201 )) / ( 3.21304406084706 - 0.03205022844106 * age + 0.00025261579629 * age_sq + 0.16192776673944 * male) - 0.00009813582463)/ 1.23822249966500)*10 ) |
| A smoother | age+age2 |
| A formula | round( 50 + (((ss_slsmaxspan - ( 6.34470335822883 + z_age * 0.18121741755095 + age_sq * -0.00181150227275 )) / ( 2.79609425763337 - 0.01551120159691 * z_age + 0.00013463536395 * age_sq) - 0.00001517101555)/ 1.23253901052392)*10 ) |
| **slstotcorr** | |
| ASE smoother | age+age2 |
| ASE formula | round( 50 + (((ss_slstotcorr - ( 3.75312190595082 + age * 0.19942501134690 + age_sq * -0.00193350470365 + male * -1.36968608085352 + educ * 0.16873908166691)) / ( 3.73330048031698 - 0.05336072872143 * age + 0.00043835342989 * age_sq) - 0.00014040532804)/ 1.26123862595018)*10 ) |
| AS smoother | age+age2+sex |
| AS formula | round( 50 + (((ss_slstotcorr - ( 7.10576195661797 + age * 0.18035014166465 + age_sq * -0.00181881431025 + male * -1.23680915349521 )) / ( 4.09162115972632 - 0.06459106313587 * age + 0.00051686908837 * age_sq + 0.12180059385210 * male) - 0.00025316711652)/ 1.25692095014987)*10 ) |
| A smoother | age+age2 |

**Table S1. T-score formulas**

| Characteristic (see Table 1 for variable abbreviations) | formula |
|---|---|
| A formula | round( 50 + (((ss_slstotcorr - ( 6.68385949646408 + z_age * 0.17602073856602 + age_sq * -0.00179393168815 )) / ( 3.56149147878272 - 0.04678963907689 * z_age + 0.00040134265172 * age_sq) - 0.00017767055051)/ 1.24649288472797)*10 ) |
| **slsretention** | |
| ASE smoother | age+age2+e+e2+e3+sex |
| ASE formula | round( 50 + (((ss_slsretention - ( 9.89636907277642 + age * 0.01891901918909 + age_sq * -0.00033731685819 + male * -0.39924841660831 + educ * 0.03782813022431)) / (24.09964821860220 - 0.05611977841787 * age + 0.00054991258056 * age_sq - 3.97683820784206 * educ + 0.25368703297025 * educ_sq - 0.00540435139935 * educ_3 + 0.42896689927867 * male) + 0.00173633716394)/ 1.26200346943920)*10 ) |
| AS smoother | age+age2+sex |
| AS formula | round( 50 + (((ss_slsretention - (10.64796795063580 + age * 0.01464279196761 + age_sq * -0.00031160542789 + male * -0.36945990725848 )) / ( 2.83886610373365 - 0.04319966945149 * age + 0.00046366123352 * age_sq + 0.36208301219812 * male) - -0.00031140530769)/ 1.26255043768947)*10 ) |
| A smoother | age+age2 |
| A formula | round( 50 + (((ss_slsretention - (10.52193715327300 + z_age * 0.01334951170462 + age_sq * -0.00030417248545 )) / ( 3.03929748945085 - 0.04446951281173 * z_age + 0.00047728768362 * age_sq) - 0.00012315878064)/ 1.26521505916373)*10 ) |
| **slsdelaycorr** | |
| ASE smoother | age+age2+e+e2+e3+sex |
| ASE formula | round( 50 + (((ss_slsdelaycorr - ( 4.74512328032410 + age * 0.16270612655085 + age_sq * -0.00165779246138 + male * -1.33898367719940 + educ * 0.18037663173053)) / (47.29417627421930 - 0.05883055869854 * age + 0.00046137956202 * age_sq - 8.46277663999085 * educ + 0.54168165627198 * educ_sq - 0.01137520862720 * educ_3 + 0.19807748973812 * male) + 0.00407491900738)/ 1.24342584383899)*10 ) |
| AS smoother | age+age2+sex |
| AS formula | round( 50 + (((ss_slsdelaycorr - ( 8.32898727774671 + age * 0.14231570656046 + age_sq * -0.00153519213281 + male * -1.19694253086374 )) / ( 3.85009780081601 - 0.05268982995979 * age + 0.00040408589843 * age_sq + 0.17036817125886 * male) - 0.00016059082064)/ 1.25109948799306)*10 ) |
| A smoother | age+age2 |
| A formula | round( 50 + (((ss_slsdelaycorr - ( 7.92068418829501 + z_age * 0.13812585504832 + age_sq * -0.00151111156340 )) / ( 3.05674049922090 - 0.02433084721133 * z_age + 0.00019984287514 * age_sq) - 0.00003276764788)/ 1.23596712265174)*10 ) |
| **symsumcorr** | |
| ASE smoother | age+age2+e+e2+e3+sex |
| ASE formula | round( 50 + (((ss_symsumcorr - ( 7.32386880632089 + age * 0.04084434015405 + age_sq * -0.00045194277547 + male * -0.24592752787878 + educ * 0.13492980377165)) / ( 8.31994691273094 - 0.05122951965336 * age + 0.00043058605285 * age_sq - 0.59768226421423 * educ + 0.02321233065813 * educ_sq - 0.00025569002036 * educ_3 + 0.14700928478075 * male) + 0.00041586028309)/ 1.21852938655792)*10 ) |
| AS smoother | age+age2+sex |
| AS formula | round( 50 + (((ss_symsumcorr - (10.00475968047210 + age * 0.02559139224317 + age_sq * -0.00036023223560 + male * -0.13967438511468 )) / ( 3.94700352021324 - 0.06112555050934 * age + 0.00052470915595 * age_sq + 0.11876586268142 * male) - -0.00010655730735)/ 1.23199025675379)*10 ) |

**Table S1. T-score formulas**

| Characteristic (see Table 1 for variable abbreviations) | formula |
|---|---|
| A smoother | age+age2 |
| A formula | round( 50 + (((ss_symsumcorr - ( 9.95711371461406 + z_age * 0.02510246740460 + age_sq * -0.00035742221036 )) / ( 3.99460797416991 - 0.06117835881752 * z_age + 0.00052734620794 * age_sq) - -0.00009025144293)/ 1.23287247336915)*10 ) |
| **symr1sec** | |
| ASE smoother | educ+educ2 |
| ASE formula | round( 50 + (((ss_symr1sec - (12.32008489176200 + age * 0.05195943167253 + age_sq * -0.00134855106389 + male * -0.47007317516347 + educ * 0.06532245840073)) / ( 8.76747989942610 - 0.85478582938899 * educ + 0.02653930843860 * educ_sq) + 0.00069954072029)/ 1.26708427481297)*10 ) |
| AS smoother | age+age2+sex |
| AS formula | round( 50 + (((ss_symr1sec - (13.61796258600380 + age * 0.04457514719612 + age_sq * -0.00130415199298 + male * -0.41863371327927 )) / ( 3.10219412931164 - 0.03587789020642 * age + 0.00027311262170 * age_sq + 0.12568085910336 * male) - 0.00007931507720)/ 1.27083904203042)*10 ) |
| A smoother | age+age2 |
| A formula | round( 50 + (((ss_symr1sec - (13.47515753557480 + z_age * 0.04310973590745 + age_sq * -0.00129572975221 )) / ( 2.92931826163197 - 0.02772495192249 * z_age + 0.00020857197348 * age_sq) - 0.00000390722328)/ 1.26753216449541)*10 ) |
| **symr2sec** | |
| ASE smoother | educ+educ2 |
| ASE formula | round( 50 + (((ss_symr2sec - (13.51428465304440 + age * 0.03251148945539 + age_sq * -0.00128114856872 + male * -0.49789091495018 + educ * 0.05515136265569)) / ( 5.36953659367157 - 0.42565583124213 * educ + 0.01304156799955 * educ_sq) + 0.00268708092069)/ 1.26747491630524)*10 ) |
| AS smoother | No smoothing |
| AS formula | round( 50 + (((ss_symr2sec - (14.61007506189190 + age * 0.02627698204274 + age_sq * -0.00124366269754 + male * -0.45446088358508 )) / ( 1 ) - 0.00000000001813)/ 2.49895826164872)*10 ) |
| A smoother | age |
| A formula | round( 50 + (((ss_symr2sec - (14.45504858566770 + z_age * 0.02468615911582 + age_sq * -0.00123451967139 )) / ( 1.84110600136458 + 0.00201197041539 * z_age) - 0.00000001253688)/ 1.26622894104795)*10 ) |
| **symr3sec** | |
| ASE smoother | educ+educ2 |
| ASE formula | round( 50 + (((ss_symr3sec - (14.37760145417320 + age * -0.00393146337679 + age_sq * -0.00096514553135 + male * -0.49646571855951 + educ * 0.06128112690478)) / ( 5.04552221410160 - 0.35611237441880 * educ + 0.01022643509109 * educ_sq) + 0.00171641897908)/ 1.25542350143982)*10 ) |
| AS smoother | No smoothing |
| AS formula | round( 50 + (((ss_symr3sec - (15.59518281340180 + age * -0.01085890127762 + age_sq * -0.00092349331608 + male * -0.44820868309552 )) / ( 1 ) - -0.00000000002185)/ 2.55062894844681)*10 ) |
| A smoother | age+age2 |

**Table S1. T-score formulas**

| Characteristic (see Table 1 for variable abbreviations) | formula |
|---|---|
| A formula | round( 50 + (((ss_symr3sec - (15.44228909853400 + z_age * -0.01242783861604 + age_sq * -0.00091447607421 )) / ( 4.46523462101881 - 0.07374928707109 * z_age + 0.00054313688271 * age_sq) - -0.00000314058082)/ 1.25431035244633)*10 ) |
| **symr4sec** | |
| ASE smoother | educ+educ2 |
| ASE formula | round( 50 + (((ss_symr4sec - (13.72167520100860 + age * 0.02799609535848 + age_sq * -0.00123694970254 + male * -0.38518603849627 + educ * 0.04443503367821)) / ( 5.45154291767762 - 0.42633619998643 * educ + 0.01286743887506 * educ_sq) + 0.00218985772381)/ 1.25922195700139)*10 ) |
| AS smoother | age+age2+sex |
| AS formula | round( 50 + (((ss_symr4sec - (14.60454520415570 + age * 0.02297300012550 + age_sq * -0.00120674762106 + male * -0.35019479234336 )) / ( 4.16267765093164 - 0.06904809190141 * age + 0.00051375735006 * age_sq + 0.17029415482739 * male) - 0.00010165167793)/ 1.25743167055086)*10 ) |
| A smoother | age+age2 |
| A formula | round( 50 + (((ss_symr4sec - (14.48508615584150 + z_age * 0.02174715665983 + age_sq * -0.00119970226191 )) / ( 4.20885231665178 - 0.06830811372255 * z_age + 0.00051182884182 * age_sq) - 0.00002500852312)/ 1.25673386174152)*10 ) |
| **symmiddle2secavg** | |
| ASE smoother | educ+educ2 |
| ASE formula | round( 50 + (((ss_symmiddle2secavg - (14.14964318287220 + age * 0.02281870304257 + age_sq * -0.00125040463962 + male * -0.48803985520487 + educ * 0.04763225158553)) / ( 5.19544955902923 - 0.40348986978354 * educ + 0.01217707230784 * educ_sq) + 0.00212218569989)/ 1.27472728794160)*10 ) |
| AS smoother | No smoothing |
| AS formula | round( 50 + (((ss_symmiddle2secavg - (15.09603801204590 + age * 0.01743418285501 + age_sq * -0.00121802943870 + male * -0.45053089657289 )) / ( 1 ) - 0.00000000000987)/ 2.45829589624120)*10 ) |
| A smoother | age+age2 |
| A formula | round( 50 + (((ss_symmiddle2secavg - (14.94235213973310 + z_age * 0.01585711669720 + age_sq * -0.00120896547761 )) / ( 3.71669573884271 - 0.05350340796140 * z_age + 0.00039003049586 * age_sq) - -0.00001558085251)/ 1.27427946297540)*10 ) |
| **symall4secavg** | |
| ASE smoother | educ+educ2 |
| ASE formula | round( 50 + (((ss_symall4secavg - (13.27701203771190 + age * 0.03894208548574 + age_sq * -0.00136378906553 + male * -0.50459842156310 + educ * 0.06812649417877)) / ( 6.58676877508067 - 0.58215043386448 * educ + 0.01782983183694 * educ_sq) + 0.00274338448246)/ 1.26773723142367)*10 ) |
| AS smoother | No smoothing |
| AS formula | round( 50 + (((ss_symall4secavg - (14.63060250791570 + age * 0.03124082276977 + age_sq * -0.00131748411743 + male * -0.45095086624430 )) / ( 1 ) - -0.00000000002270)/ 2.45974266896199)*10 ) |
| A smoother | age3 |
| A formula | round( 50 + (((ss_symall4secavg - (14.47677337482810 + z_age * 0.02966228652420 + age_sq * -0.00130841170722 )) / ( 1.93076541447127 + 0.00000004101661 * age_3) - -0.00000039207609)/ 1.26940422930785)*10 ) |

**Table S2.** Self-reported location, environment, and subtest interference characteristics in the normative sample (database variable name).

| | Overall (N=1240) |
|---|---|
| **Location selection (location)** | |
| At home | 1140 (91.9%) |
| At work | 82 (6.6%) |
| In a clinic (medical or research center) | 9 (0.7%) |
| In a public space (park, library) | 9 (0.7%) |
| **Noise in testing environment selection endorsed as Yes (environment)** | 55 (4.4%) |
| Missing (by design, only asked if noise endorsed) | 1185 (95.6%) |
| People were talking in the background. | 7 (0.6%) |
| People were talking to me while I tried to take the test. | 1 (0.1%) |
| There was some noise in the background and it was distracting. | 21 (1.7%) |
| There was some noise in the background, but it did not distract me. | 26 (2.1%) |
| **SLS Trials 1-5 interference endorsed as Yes (slsinterf)** | 110 (8.9%) |
| Missing (by design, only asked if interference endorsed) | 1130 (91.1%) |
| I am not comfortable using technology. | 2 (0.2%) |
| I had technical problems. | 5 (0.4%) |
| I was confused about the instructions. | 1 (0.1%) |
| I was interrupted during this test. | 51 (4.1%) |
| Other (there will be a comments box at the end of the session). | 44 (3.5%) |
| Sometimes my selection did not register. | 6 (0.5%) |
| The words were hard for me to see. | 1 (0.1%) |
| **SLS Delay interference endorsed as Yes (slsdelayinterf)** | 20 (1.6%) |
| Missing (by design, only asked if interference endorsed) | 1220 (98.4%) |
| I am not comfortable using technology. | 1 (0.1%) |
| I had technical problems. | 2 (0.2%) |
| I was interrupted during this test. | 1 (0.1%) |
| Other (there will be a comments box at the end of the session). | 13 (1.0%) |
| Sometimes my selection did not register. | 2 (0.2%) |

| | Overall (N=1240) |
|---|---|
| The words were hard for me to see. | 1 (0.1%) |
| **Symbols Test interference endorsed as Yes (syminterf)** | 68 (5.5%) |
| Missing (by design, only asked if interference endorsed) | 1173 (94.6%) |
| I am not comfortable using technology. | 1 (0.1%) |
| I had technical problems. | 4 (0.3%) |
| I was confused about the instructions. | 3 (0.2%) |
| I was interrupted during this test. | 16 (1.3%) |
| Other (there will be a comments box at the end of the session). | 32 (2.6%) |
| Sometimes my selection did not register. | 10 (0.8%) |
| The symbols were hard for me to see. | 1 (0.1%) |

**Table S3**. Pearson correlations and adjusted R-squared values where the raw score is the outcome and each individual demographic variable is the predictor in a separate linear regression model, except for the last column that presents the adjusted r-squared when all demographic variances are in the model.

| | **Pearson Correlations** [a] | | | **Individual Adj. $R^2$** [b,c] | | | | **Combined Adj. $R^2$** |
|---|---|---|---|---|---|---|---|---|
| **MTD Measure (Raw)** | **Age** | **Male Sex** | **Educ** | **Age** | **Age+Age Squared** | **Male Sex** | **Education** | **All** |
| **Primary Variables** | | | | | | | | |
| mtdrawcompositev1 | -0.39 | -0.22 | 0.14 | 0.1526 | 0.1738 | 0.0455 | 0.0191 | 0.2378 |
| slssumoftrials | -0.27 | -0.22 | 0.11 | 0.0746 | 0.0867 | 0.0477 | 0.0124 | 0.1495 |
| symall4corrartaccweighted | -0.50 | -0.09 | 0.14 | 0.2513 | 0.2773 | 0.0072 | 0.0182 | 0.2949 |
| **Secondary Variables** | | | | | | | | |
| slsr1corr | -0.21 | -0.09 | 0.04 | 0.0450 | 0.0483 | 0.0068 | 0.0010 | 0.0553 |
| slsr2corr | -0.25 | -0.14 | 0.13 | 0.0641 | 0.0717 | 0.0175 | 0.0172 | 0.1064 |
| slsr3corr | -0.23 | -0.20 | 0.08 | 0.0540 | 0.0627 | 0.0369 | 0.0061 | 0.1073 |
| slsr4corr | -0.23 | -0.22 | 0.09 | 0.0507 | 0.0606 | 0.0471 | 0.0065 | 0.1169 |
| slsr5corr | -0.24 | -0.22 | 0.12 | 0.0595 | 0.0742 | 0.0457 | 0.0131 | 0.1369 |
| slsmaxspan | -0.27 | -0.22 | 0.11 | 0.0632 | 0.0749 | 0.0480 | 0.0111 | 0.1372 |
| slstotcorr | -0.25 | -0.22 | 0.11 | 0.0720 | 0.0849 | 0.0458 | 0.0115 | 0.1448 |
| slsdelaycorr | -0.26 | -0.21 | 0.11 | 0.0668 | 0.0742 | 0.0434 | 0.0121 | 0.1320 |
| slsretention | -0.12 | -0.07 | 0.04 | 0.0133 | 0.0130 | 0.0043 | 0.0004 | 0.0176 |
| symsumcorr | -0.11 | -0.03 | 0.12 | 0.0117 | 0.0130 | 0.0002 | 0.0132 | 0.0258 |
| symr1sec | 0.53 | 0.07 | -0.07 | 0.2074 | 0.2318 | 0.0033 | 0.0049 | 0.2360 |
| symr2sec | 0.46 | 0.06 | -0.08 | 0.2598 | 0.2856 | 0.0043 | 0.0024 | 0.2890 |
| symr3sec | 0.51 | 0.07 | -0.06 | 0.2323 | 0.2512 | 0.0034 | 0.0060 | 0.2557 |
| symr4sec | 0.48 | 0.06 | -0.08 | 0.2617 | 0.2852 | 0.0046 | 0.0053 | 0.2903 |
| symmiddle2secavg | 0.51 | 0.07 | -0.08 | 0.2803 | 0.3067 | 0.0049 | 0.0042 | 0.3114 |
| symall4secavg | 0.53 | 0.07 | -0.08 | 0.2803 | 0.3081 | 0.0047 | 0.0056 | 0.3133 |
| symall4corrartsec | 0.53 | 0.07 | -0.08 | 0.2794 | 0.3069 | 0.0047 | 0.0057 | 0.3123 |

[a] $p < .001$ for all Pearson correlations except sex and Symbols total correct ($p = 0.26$), education and SLS trial 1 correct ($p = 0.13$), and education and SLS retention ($p = .21$).

[b] Adjusted r-square value multiplied by 100 is the percentage of variance explained by the predictor. The adjusted r-square provides a more conservative r-square value that adjusts for the number of terms in the model.
[c] Most *p*-values were < .05 for the adjusted r-square with the following exceptions: sex and Symbols total correct ($p$=.27); education and SLS retention ($p$=.21).
*Note.* See manuscript Table 1 for MTD variable abbreviation key. Some of these results are subtly different than results in manuscript Table 6 because here raw scores are used as the outcome whereas Table 2 uses uncorrected scaled score as the outcome.

**Table S4.** Model estimates for unadjusted and fully-adjusted models to illustrate the magnitude of effects for interference, device and input source variables.

| | | Unadjusted | | | Age+Sex+Educ Adjusted | | |
|---|---|---|---|---|---|---|---|
| Outcome | N | Mean estimate (95% CI) | *p* | Adj. *p* | Mean estimate (95% CI) | *p* | Adj. *p* |
| **SS mtdrawcompositev1** | | | | | | | |
| Any Interference | 1240 | -0.16 (-0.62, 0.30) | 0.50 | 0.50 | -0.69 (-1.10, -0.28) | <.001 | <.001 |
| Touch | | | | | | | |
| *Mouse* | 660 | ref | - | 0.07 | ref | - | 0.12 |
| *Touch* | 548 | 0.33 (-0.01, 0.67) | 0.06 | | -0.15 (-0.46, 0.16) | 0.34 | |
| *Stylus* | 22 | -0.87 (-2.15, 0.40) | 0.18 | | -0.95 (-2.08, 0.18) | 0.10 | |
| *Other* | 10 | -0.77 (-2.65, 1.11) | 0.42 | | -0.46 (-2.12, 1.20) | 0.58 | |
| Device | | | | | | | |
| *Desktop computer or laptop* | 795 | ref | - | 0.002 | ref | - | 0.004 |
| *Smartphone* | 264 | 0.55 (0.13, 0.96) | 0.01 | | -0.21 (-0.59, 0.16) | 0.27 | |
| *Tablet* | 178 | -0.51 (-0.99, -0.02) | 0.04 | | -0.64 (-1.08, -0.21) | 0.004 | |
| *Other / not sure* | 3 | -2.30 (-5.70, 1.11) | 0.19 | | -0.75 (-3.76, 2.26) | 0.63 | |
| **SS slssumoftrials** | | | | | | | |
| Any Interference | 1240 | -0.13 (-0.59, 0.33) | 0.58 | 0.58 | -0.56 (-1.00, -0.13) | 0.01 | 0.01 |
| Touch | | | | | | | |
| *Mouse* | 660 | ref | - | 0.04 | ref | - | 0.28 |
| *Touch* | 548 | 0.36 (0.02, 0.70) | 0.04 | | -0.05 (-0.37, 0.28) | 0.78 | |
| *Stylus* | 22 | -1.09 (-2.36, 0.19) | 0.10 | | -1.22 (-2.41, -0.03) | 0.045 | |
| *Other* | 10 | -0.60 (-2.48, 1.27) | 0.53 | | -0.38 (-2.13, 1.37) | 0.67 | |
| Device | | | | | | | |
| *Desktop computer or laptop* | 795 | ref | - | 0.03 | ref | - | 0.02 |

| Outcome | N | Unadjusted<br>Mean estimate (95% CI) | *p* | Adj. *p* | Age+Sex+Educ Adjusted<br>Mean estimate (95% CI) | *p* | Adj. *p* |
|---|---|---|---|---|---|---|---|
| *Smartphone* | 264 | 0.43 (0.01, 0.85) | 0.045 | | -0.16 (-0.56, 0.24) | 0.44 | |
| *Tablet* | 178 | -0.41 (-0.90, 0.08) | 0.10 | | -0.61 (-1.07, -0.15) | 0.01 | |
| *Other / not sure* | 3 | -0.67 (-4.08, 2.73) | 0.70 | | 0.54 (-2.63, 3.71) | 0.74 | |
| **SS symall4corrartsecweighted** | | | | | | | |
| Any Interference | 1240 | -0.19 (-0.65, 0.27) | 0.41 | 0.41 | -0.75 (-1.12, -0.37) | <.001 | <.001 |
| Touch | | | | | | | |
| *Mouse* | 660 | ref | - | 0.82 | ref | - | 0.03 |
| *Touch* | 548 | 0.09 (-0.26, 0.43) | 0.62 | | -0.38 (-0.66, -0.10) | 0.009 | |
| *Stylus* | 22 | -0.20 (-1.48, 1.08) | 0.76 | | -0.05 (-1.10, 1.00) | 0.93 | |
| *Other* | 10 | -0.67 (-2.56, 1.21) | 0.49 | | -0.22 (-1.75, 1.32) | 0.78 | |
| Device | | | | | | | |
| *Desktop computer or laptop* | 795 | ref | - | 0.003 | ref | - | 0.055 |
| *Smartphone* | 264 | 0.50 (0.08, 0.91) | 0.02 | | -0.39 (-0.74, -0.04) | 0.03 | |
| *Tablet* | 178 | -0.36 (-0.85, 0.13) | 0.15 | | -0.22 (-0.63, 0.18) | 0.28 | |
| *Other / not sure* | 3 | -3.62 (-7.03, -0.22) | 0.04 | | -1.92 (-4.71, 0.86) | 0.18 | |

*Note*. Secondary variables and age-adjusted & age- and sex-adjusted models not shown. Adj. *p* = adjusted for age, sex and education. Other p-value (*p*) is unadjusted. SS mtdrawcompositev1 = unadjusted scaled score for MTD Composite. SS slssumoftrials = unadjusted scaled score for SLS Sum of Trials. SS symall4corrartsecweighted = unadjusted scaled score for Accuracy-Weighted Symbols.

**Table S5.** Normative score (unadjusted scaled score and T-score) statistics by categorized age, sex, or education to show that mean and SD values for the T-score statistics are within expected ranges (mean 50 +/-3; SD 10 +/- .6). See manuscript Table 1 for MTD variable (outcome) abbreviation key. T-ASE adj = age-, sex- and education-adjusted T-scores; T-AS adj = age- and sex-adjusted T-scores; T-A adj = age-adjusted T-scores. SS unadj. = unadjusted scaled scores from manuscript Tables 2-4.

**Table S5. Normative score statistics by categorized age, sex, and education**

| | Age | | | | | Sex | | | Years of Education | | | | |
|---|---|---|---|---|---|---|---|---|---|---|---|---|---|
| Outcome | 30-59 N=233 | 60-69 N=375 | 70-79 N=385 | 80+ N=247 | p † | Female N=639 | Male N=601 | p † | <=12 N=140 | 13-15 N=318 | 16 N=373 | 17+ N=409 | p † |
| **mtdrawcompositev1** | | | | | | | | | | | | | |
| SS unadj | 11.40 (2.84) | 10.85 (2.64) | 9.64 (2.77) | 7.94 (2.87) | <0.001 | 10.63 (2.90) | 9.33 (3.00) | <0.001 | 9.12 (3.10) | 9.69 (2.76) | 10.07 (3.03) | 10.48 (3.08) | <0.001 |
| T-ASE adj | 49.55 (10.23) | 50.72 (9.65) | 49.82 (10.23) | 49.60 (9.90) | 0.40 | 50.02 (9.71) | 49.97 (10.30) | 0.93 | 50.27 (10.09) | 49.85 (9.62) | 49.87 (9.94) | 50.13 (10.32) | 0.96 |
| T-AS adj | 49.53 (10.21) | 50.74 (9.71) | 49.86 (10.20) | 49.56 (9.90) | 0.37 | 50.04 (9.99) | 49.97 (10.02) | 0.91 | 47.67 (10.12) | 48.57 (9.64) | 49.98 (9.75) | 51.95 (10.13) | <0.001 |
| T-A adj | 49.72 (10.13) | 50.61 (9.89) | 49.81 (10.17) | 49.61 (9.86) | 0.55 | 52.24 (9.45) | 47.61 (10.05) | <0.001 | 48.26 (10.43) | 49.07 (9.54) | 49.61 (9.88) | 51.66 (10.15) | <0.001 |
| **slssumoftrials** | | | | | | | | | | | | | |
| SS unadj | 10.91 (2.77) | 10.74 (2.78) | 9.81 (2.95) | 8.51 (3.04) | <0.001 | 10.67 (2.85) | 9.36 (3.04) | <0.001 | 9.39 (3.27) | 9.78 (2.83) | 10.08 (3.03) | 10.42 (2.99) | 0.001 |
| T-ASE adj | 49.49 (10.04) | 50.76 (9.82) | 49.83 (10.19) | 49.53 (9.91) | 0.33 | 50.03 (10.00) | 49.95 (10.00) | 0.89 | 50.30 (9.99) | 49.75 (10.33) | 50.06 (9.78) | 50.01 (9.97) | 0.95 |
| T-AS adj | 49.55 (10.04) | 50.80 (9.69) | 49.82 (10.28) | 49.47 (9.99) | 0.30 | 50.02 (9.99) | 49.98 (10.02) | 0.94 | 48.15 (10.53) | 48.68 (9.75) | 50.09 (9.90) | 51.57 (9.89) | <0.001 |
| T-A adj | 49.69 (9.94) | 50.65 (9.91) | 49.86 (10.24) | 49.47 (9.83) | 0.46 | 52.16 (9.38) | 47.68 (10.14) | <0.001 | 48.64 (10.81) | 49.22 (9.59) | 49.73 (10.06) | 51.29 (9.87) | 0.009 |
| **Symall4weighted** | | | | | | | | | | | | | |
| SS unadj | 12.35 (2.64) | 10.84 (2.43) | 9.38 (2.58) | 7.48 (2.56) | <0.001 | 10.24 (3.02) | 9.74 (2.99) | 0.004 | 9.44 (3.08) | 9.81 (2.91) | 10.07 (3.09) | 10.28 (2.99) | 0.02 |
| T-ASE adj | 49.61 (10.30) | 50.29 (9.60) | 49.94 (10.31) | 49.95 (9.96) | 0.88 | 49.96 (9.91) | 50.01 (10.15) | 0.93 | 50.66 (9.96) | 50.01 (9.89) | 49.30 (10.26) | 50.36 (9.92) | 0.40 |
| T-AS adj | 49.66 (10.28) | 50.28 (9.64) | 49.93 (10.21) | 50.05 (10.06) | 0.90 | 50.01 (9.92) | 50.00 (10.12) | 0.98 | 49.74 (11.26) | 49.48 (9.99) | 49.40 (9.74) | 51.06 (9.78) | 0.08 |
| T-A adj | 49.65 (9.95) | 50.24 (9.89) | 49.93 (10.20) | 50.04 (9.91) | 0.91 | 50.86 (9.85) | 49.07 (10.07) | 0.002 | 49.89 (11.31) | 49.70 (10.06) | 49.28 (9.66) | 50.91 (9.72) | 0.13 |
| **Symall4corrartsec** | | | | | | | | | | | | | |
| SS unadj | 12.31 (2.80) | 10.88 (2.49) | 9.34 (2.50) | 7.51 (2.47) | <0.001 | 10.29 (2.99) | 9.69 (3.01) | <0.001 | 8.96 (3.01) | 9.68 (2.80) | 10.19 (3.05) | 10.43 (3.05) | <0.001 |
| T-ASE adj | 49.56 (10.00) | 50.54 (9.79) | 49.81 (10.34) | 49.92 (9.79) | 0.64 | 50.00 (10.01) | 50.02 (9.99) | 0.97 | 50.04 (10.01) | 50.11 (10.11) | 49.79 (9.82) | 50.11 (10.10) | 0.97 |
| T-AS adj | 49.57 (9.99) | 50.56 (9.91) | 49.80 (10.21) | 49.86 (9.79) | 0.61 | 50.01 (10.01) | 49.99 (9.98) | 0.97 | 47.70 (10.67) | 48.98 (10.15) | 49.89 (9.44) | 51.68 (9.89) | <0.001 |
| T-A adj | 49.62 (10.06) | 50.50 (9.92) | 49.80 (10.16) | 49.93 (9.89) | 0.70 | 51.08 (9.83) | 48.86 (10.07) | <0.001 | 48.02 (10.79) | 49.20 (10.13) | 49.74 (9.48) | 51.55 (9.92) | <0.001 |
| **slsr1corr** | | | | | | | | | | | | | |

**Table S5. Normative score statistics by categorized age, sex, and education**

| | Age | | | | | Sex | | | Years of Education | | | | |
|---|---|---|---|---|---|---|---|---|---|---|---|---|---|
| Outcome | 30-59 N=233 | 60-69 N=375 | 70-79 N=385 | 80+ N=247 | p † | Female N=639 | Male N=601 | p † | <=12 N=140 | 13-15 N=318 | 16 N=373 | 17+ N=409 | p † |
| SS unadj | 10.62 (3.05) | 10.43 (3.03) | 9.70 (3.12) | 8.65 (3.44) | <0.001 | 10.17 (3.23) | 9.58 (3.20) | 0.001 | 9.76 (3.56) | 9.69 (3.24) | 10.00 (3.16) | 9.97 (3.16) | 0.56 |
| T-ASE adj | 49.43 (9.69) | 50.64 (9.93) | 50.04 (10.03) | 49.49 (10.33) | 0.40 | 49.96 (10.09) | 50.03 (9.91) | 0.90 | 50.66 (10.10) | 49.60 (9.94) | 50.14 (10.02) | 49.94 (10.01) | 0.75 |
| T-AS adj | 49.42 (9.65) | 50.68 (10.02) | 50.08 (9.96) | 49.41 (10.36) | 0.33 | 49.99 (10.09) | 50.03 (9.92) | 0.95 | 50.14 (10.63) | 49.36 (9.98) | 50.16 (9.75) | 50.32 (10.04) | 0.60 |
| T-A adj | 49.48 (10.02) | 50.63 (9.80) | 50.09 (9.82) | 49.43 (10.59) | 0.40 | 50.90 (10.05) | 49.05 (9.88) | 0.001 | 50.36 (10.82) | 49.53 (9.93) | 50.05 (9.77) | 50.21 (10.03) | 0.78 |
| **slsr2corr** | | | | | | | | | | | | | |
| SS unadj | 10.96 (2.92) | 10.69 (2.73) | 9.76 (3.00) | 8.56 (3.25) | <0.001 | 10.44 (3.04) | 9.59 (3.05) | <0.001 | 9.24 (3.22) | 9.81 (3.02) | 9.95 (3.10) | 10.54 (2.98) | <0.001 |
| T-ASE adj | 49.64 (10.40) | 50.80 (9.70) | 49.85 (10.09) | 49.38 (9.98) | 0.29 | 50.00 (10.04) | 50.00 (9.99) | 1.00 | 50.16 (10.21) | 50.19 (10.08) | 49.51 (10.02) | 50.25 (9.90) | 0.73 |
| T-AS adj | 49.73 (10.27) | 50.82 (9.68) | 49.78 (10.14) | 49.32 (9.97) | 0.26 | 49.99 (10.00) | 50.00 (10.00) | 0.99 | 47.90 (10.14) | 49.06 (10.02) | 49.61 (9.90) | 51.79 (9.78) | <0.001 |
| T-A adj | 49.76 (10.20) | 50.77 (9.73) | 49.81 (10.16) | 49.34 (9.90) | 0.31 | 51.37 (9.89) | 48.53 (9.90) | <0.001 | 48.35 (10.25) | 49.38 (9.97) | 49.33 (9.96) | 51.64 (9.76) | <0.001 |
| **slsr3corr** | | | | | | | | | | | | | |
| SS unadj | 10.74 (2.70) | 10.78 (2.70) | 9.82 (2.93) | 8.75 (3.14) | <0.001 | 10.62 (2.83) | 9.48 (3.00) | <0.001 | 9.52 (3.31) | 9.90 (3.00) | 10.12 (2.80) | 10.34 (2.94) | 0.02 |
| T-ASE adj | 49.29 (10.33) | 51.09 (9.74) | 49.69 (10.15) | 49.55 (9.79) | 0.09 | 50.00 (9.72) | 50.02 (10.31) | 0.97 | 50.07 (10.56) | 49.91 (10.29) | 50.08 (9.57) | 50.01 (10.01) | 1.00 |
| T-AS adj | 49.30 (10.25) | 51.12 (9.72) | 49.66 (10.15) | 49.50 (9.91) | 0.07 | 50.01 (9.99) | 49.99 (10.04) | 0.98 | 48.46 (10.63) | 49.07 (10.31) | 50.14 (9.46) | 51.13 (9.93) | <0.01 |
| T-A adj | 49.39 (10.22) | 51.00 (9.90) | 49.67 (10.06) | 49.56 (9.79) | 0.14 | 51.89 (9.55) | 47.99 (10.08) | <0.001 | 48.93 (10.82) | 49.51 (10.19) | 49.82 (9.54) | 50.91 (9.93) | 0.12 |
| **slsr4corr** | | | | | | | | | | | | | |
| SS unadj | 10.69 (2.94) | 10.62 (2.83) | 9.82 (2.90) | 8.72 (3.10) | <0.001 | 10.61 (2.84) | 9.36 (3.06) | <0.001 | 9.36 (3.25) | 9.90 (2.91) | 9.98 (2.91) | 10.33 (3.07) | 0.009 |
| T-ASE adj | 49.58 (10.50) | 50.69 (9.82) | 49.82 (9.87) | 49.62 (9.98) | 0.44 | 50.02 (9.54) | 49.98 (10.47) | 0.95 | 49.81 (10.51) | 50.14 (9.83) | 49.84 (9.69) | 50.10 (10.25) | 0.97 |
| T-AS adj | 49.57 (10.41) | 50.71 (9.87) | 49.81 (9.88) | 49.59 (10.04) | 0.41 | 50.02 (9.55) | 49.97 (10.48) | 0.93 | 47.99 (10.53) | 49.22 (9.81) | 49.92 (9.62) | 51.36 (10.18) | 0.002 |
| T-A adj | 49.74 (10.43) | 50.62 (9.94) | 49.83 (9.91) | 49.63 (9.85) | 0.56 | 52.07 (9.31) | 47.83 (10.25) | <0.001 | 48.60 (10.77) | 49.73 (9.74) | 49.55 (9.71) | 51.14 (10.11) | 0.03 |
| **slsr5corr** | | | | | | | | | | | | | |
| SS unadj | 10.78 (2.67) | 10.52 (2.73) | 9.79 (2.88) | 8.56 (2.88) | <0.001 | 10.53 (2.75) | 9.34 (2.93) | <0.001 | 9.34 (3.19) | 9.65 (2.79) | 10.01 (2.88) | 10.36 (2.85) | <0.001 |
| T-ASE adj | 50.01 (10.11) | 50.35 (9.59) | 49.86 (10.26) | 49.66 (10.13) | 0.85 | 49.98 (10.00) | 50.02 (10.00) | 0.95 | 50.58 (10.39) | 49.70 (9.81) | 50.06 (9.88) | 49.97 (10.14) | 0.86 |
| T-AS adj | 50.12 (9.89) | 50.38 (9.82) | 49.81 (10.41) | 49.60 (9.79) | 0.77 | 50.01 (10.07) | 49.98 (9.94) | 0.96 | 48.17 (10.73) | 48.46 (9.97) | 50.18 (9.75) | 51.66 (9.75) | <0.001 |
| T-A adj | 50.16 (9.76) | 50.25 (10.00) | 49.85 (10.32) | 49.70 (9.73) | 0.89 | 52.03 (9.40) | 47.84 (10.16) | <0.001 | 48.66 (10.99) | 48.98 (9.80) | 49.80 (9.95) | 51.44 (9.68) | 0.002 |
| **slsmaxspan** | | | | | | | | | | | | | |
| SS unadj | 10.79 (2.80) | 10.52 (2.91) | 9.73 (2.99) | 8.50 (3.06) | <0.001 | 10.54 (2.89) | 9.27 (3.09) | <0.001 | 9.30 (3.24) | 9.67 (2.92) | 9.90 (3.03) | 10.36 (3.06) | <0.001 |
| T-ASE adj | 49.85 (9.97) | 50.43 (9.65) | 49.89 (10.13) | 49.69 (10.37) | 0.79 | 50.00 (9.90) | 50.02 (10.11) | 0.97 | 50.43 (10.41) | 49.85 (9.80) | 49.80 (9.92) | 50.17 (10.11) | 0.90 |
| T-AS adj | 49.88 (9.78) | 50.49 (9.77) | 49.87 (10.24) | 49.62 (10.22) | 0.72 | 50.02 (9.93) | 50.00 (10.10) | 0.98 | 48.21 (10.42) | 48.75 (9.82) | 49.90 (9.80) | 51.71 (9.96) | <0.001 |
| T-A adj | 50.01 (9.64) | 50.34 (9.97) | 49.87 (10.20) | 49.68 (10.16) | 0.86 | 52.06 (9.37) | 47.82 (10.21) | <0.001 | 48.73 (10.75) | 49.26 (9.71) | 49.52 (9.94) | 51.45 (9.91) | 0.004 |

**Table S5. Normative score statistics by categorized age, sex, and education**

| | Age | | | | | Sex | | | Years of Education | | | | |
|---|---|---|---|---|---|---|---|---|---|---|---|---|---|
| Outcome | 30-59 N=233 | 60-69 N=375 | 70-79 N=385 | 80+ N=247 | p † | Female N=639 | Male N=601 | p † | <=12 N=140 | 13-15 N=318 | 16 N=373 | 17+ N=409 | p † |
| **slstotcorr** | | | | | | | | | | | | | |
| SS unadj | 10.83 (2.77) | 10.69 (2.79) | 9.78 (2.91) | 8.47 (3.03) | <0.001 | 10.60 (2.86) | 9.34 (3.00) | <0.001 | 9.35 (3.26) | 9.74 (2.90) | 10.01 (2.99) | 10.38 (2.93) | 0.001 |
| T-ASE adj | 49.57 (9.99) | 50.77 (9.85) | 49.87 (10.20) | 49.44 (9.99) | 0.33 | 50.00 (9.78) | 50.00 (10.28) | 1.00 | 50.28 (10.53) | 49.84 (9.88) | 49.93 (10.10) | 50.09 (9.90) | 0.97 |
| T-AS adj | 49.59 (9.96) | 50.80 (9.83) | 49.83 (10.17) | 49.39 (10.01) | 0.28 | 50.01 (9.98) | 49.97 (10.02) | 0.96 | 48.16 (10.57) | 48.73 (9.90) | 49.98 (9.92) | 51.60 (9.73) | <0.001 |
| T-A adj | 49.70 (9.90) | 50.69 (10.02) | 49.87 (10.18) | 49.44 (9.85) | 0.42 | 52.09 (9.50) | 47.77 (10.07) | <0.001 | 48.71 (10.83) | 49.26 (9.81) | 49.66 (10.07) | 51.32 (9.73) | 0.009 |
| **slsretention** | | | | | | | | | | | | | |
| SS unadj | 10.23 (2.59) | 10.13 (2.66) | 9.84 (3.08) | 9.49 (3.51) | 0.02 | 10.12 (2.77) | 9.74 (3.17) | 0.02 | 9.74 (3.40) | 9.70 (2.90) | 10.13 (3.07) | 9.99 (2.78) | 0.23 |
| T-ASE adj | 49.30 (10.07) | 50.29 (9.58) | 50.10 (10.20) | 50.03 (10.18) | 0.68 | 50.00 (10.09) | 49.98 (9.87) | 0.97 | 49.84 (10.04) | 49.26 (9.80) | 50.73 (10.29) | 49.93 (9.81) | 0.29 |
| T-AS adj | 49.32 (10.06) | 50.27 (9.51) | 50.05 (10.29) | 50.11 (10.32) | 0.71 | 49.98 (10.09) | 50.01 (9.95) | 0.96 | 49.38 (11.27) | 48.98 (10.06) | 50.74 (10.34) | 50.30 (9.15) | 0.10 |
| T-A adj | 49.40 (9.97) | 50.19 (9.61) | 50.06 (10.27) | 50.05 (10.17) | 0.81 | 50.57 (9.28) | 49.34 (10.67) | 0.03 | 49.51 (11.06) | 49.19 (9.88) | 50.58 (10.42) | 50.19 (9.26) | 0.28 |
| **slsdelaycorr** | | | | | | | | | | | | | |
| SS unadj | 10.89 (2.86) | 10.57 (2.86) | 9.79 (2.95) | 8.60 (3.03) | <0.001 | 10.59 (2.87) | 9.37 (3.07) | <0.001 | 9.28 (3.16) | 9.64 (2.76) | 10.11 (3.03) | 10.42 (3.12) | <0.001 |
| T-ASE adj | 49.58 (9.98) | 50.50 (9.89) | 49.97 (10.14) | 49.71 (9.97) | 0.67 | 50.04 (10.04) | 49.97 (9.96) | 0.90 | 50.23 (9.91) | 49.52 (10.35) | 50.29 (9.80) | 50.05 (9.95) | 0.77 |
| T-AS adj | 49.60 (9.97) | 50.49 (9.77) | 49.96 (10.21) | 49.63 (10.10) | 0.66 | 49.99 (9.98) | 49.99 (10.04) | 1.00 | 47.91 (10.47) | 48.35 (9.47) | 50.34 (9.71) | 51.65 (10.22) | <0.001 |
| T-A adj | 49.78 (9.84) | 50.37 (9.88) | 49.96 (10.18) | 49.72 (10.09) | 0.84 | 51.99 (9.40) | 47.88 (10.19) | <0.001 | 48.42 (10.71) | 48.91 (9.31) | 49.98 (9.90) | 51.41 (10.20) | 0.001 |
| **symsumcorr** | | | | | | | | | | | | | |
| SS unadj | 10.30 (2.78) | 10.05 (2.73) | 9.86 (2.98) | 9.45 (3.23) | 0.01 | 9.99 (2.86) | 9.84 (3.00) | 0.37 | 9.11 (3.28) | 9.88 (3.04) | 9.97 (2.82) | 10.18 (2.78) | 0.003 |
| T-ASE adj | 50.06 (10.05) | 49.93 (9.71) | 50.01 (10.28) | 50.00 (10.00) | 1.00 | 49.98 (9.97) | 50.00 (10.04) | 0.97 | 49.29 (9.66) | 50.67 (10.19) | 49.98 (9.94) | 49.72 (10.04) | 0.49 |
| T-AS adj | 50.01 (10.01) | 50.00 (9.87) | 50.15 (10.24) | 49.93 (10.09) | 0.99 | 50.03 (10.07) | 50.04 (10.03) | 0.99 | 47.63 (10.93) | 49.89 (10.62) | 50.09 (9.63) | 50.92 (9.52) | 0.01 |
| T-A adj | 50.10 (9.91) | 50.00 (9.87) | 50.13 (10.32) | 49.96 (10.04) | 1.00 | 50.28 (9.82) | 49.81 (10.27) | 0.41 | 47.69 (10.88) | 49.94 (10.56) | 50.06 (9.70) | 50.93 (9.53) | 0.01 |
| **symr1sec** | | | | | | | | | | | | | |
| SS unadj | 12.09 (2.69) | 10.73 (2.54) | 9.54 (2.72) | 7.64 (2.60) | <0.001 | 10.23 (3.01) | 9.76 (3.01) | 0.006 | 9.43 (3.22) | 9.77 (2.87) | 10.13 (3.03) | 10.25 (3.01) | 0.02 |
| T-ASE adj | 49.68 (10.03) | 50.13 (9.66) | 50.29 (10.43) | 49.67 (9.81) | 0.83 | 49.97 (9.76) | 50.04 (10.25) | 0.91 | 50.43 (9.91) | 49.81 (9.78) | 49.71 (10.41) | 50.27 (9.83) | 0.80 |
| T-AS adj | 49.64 (9.94) | 50.05 (9.91) | 50.28 (10.47) | 49.73 (9.53) | 0.85 | 49.99 (10.04) | 49.98 (9.98) | 0.98 | 49.49 (11.62) | 49.35 (9.96) | 49.74 (9.65) | 50.87 (9.75) | 0.16 |
| T-A adj | 49.73 (9.93) | 50.01 (9.90) | 50.30 (10.47) | 49.79 (9.56) | 0.89 | 50.81 (9.73) | 49.15 (10.24) | 0.004 | 49.71 (11.59) | 49.53 (9.88) | 49.64 (9.66) | 50.81 (9.84) | 0.26 |
| **symr2sec** | | | | | | | | | | | | | |
| SS unadj | 12.28 (2.60) | 10.77 (2.49) | 9.42 (2.64) | 7.58 (2.62) | <0.001 | 10.25 (3.04) | 9.74 (2.97) | 0.003 | 9.62 (2.95) | 9.79 (2.89) | 10.03 (3.10) | 10.27 (3.04) | 0.07 |
| T-ASE adj | 49.70 (9.89) | 50.12 (9.73) | 50.07 (10.37) | 50.00 (10.04) | 0.96 | 49.97 (10.00) | 50.03 (10.04) | 0.91 | 51.12 (9.88) | 49.82 (9.76) | 49.23 (10.13) | 50.46 (10.12) | 0.18 |
| T-AS adj | 49.69 (9.93) | 50.11 (9.72) | 50.04 (10.32) | 50.06 (10.07) | 0.96 | 49.99 (9.99) | 50.01 (10.04) | 0.97 | 50.39 (10.79) | 49.40 (9.84) | 49.27 (9.73) | 51.00 (10.06) | 0.06 |

**Table S5. Normative score statistics by categorized age, sex, and education**

| | Age | | | | | Sex | | | Years of Education | | | | |
|---|---|---|---|---|---|---|---|---|---|---|---|---|---|
| Outcome | 30-59 N=233 | 60-69 N=375 | 70-79 N=385 | 80+ N=247 | p † | Female N=639 | Male N=601 | p † | <=12 N=140 | 13-15 N=318 | 16 N=373 | 17+ N=409 | p † |
| T-A adj | 49.71 (10.11) | 50.06 (9.77) | 50.04 (10.27) | 50.09 (9.89) | 0.97 | 50.87 (9.92) | 49.07 (10.01) | 0.002 | 50.59 (10.85) | 49.60 (9.83) | 49.11 (9.74) | 50.90 (10.02) | 0.06 |
| **symr3sec** | | | | | | | | | | | | | |
| SS unadj | 12.19 (2.74) | 10.80 (2.49) | 9.36 (2.64) | 7.72 (2.67) | <0.001 | 10.24 (3.03) | 9.74 (2.98) | 0.003 | 9.56 (3.09) | 9.83 (2.99) | 9.98 (3.01) | 10.30 (2.99) | 0.04 |
| T-ASE adj | 49.61 (10.38) | 50.49 (9.50) | 49.78 (10.19) | 49.98 (10.05) | 0.70 | 49.98 (10.00) | 50.02 (9.98) | 0.95 | 50.82 (9.90) | 50.02 (9.90) | 49.10 (9.91) | 50.53 (10.14) | 0.16 |
| T-AS adj | 49.56 (10.34) | 50.49 (9.50) | 49.76 (10.18) | 50.02 (10.19) | 0.67 | 49.98 (10.08) | 50.01 (9.94) | 0.95 | 49.97 (11.00) | 49.57 (10.18) | 49.15 (9.63) | 51.10 (9.79) | 0.04 |
| T-A adj | 49.62 (9.79) | 50.43 (9.85) | 49.77 (10.43) | 50.00 (9.70) | 0.75 | 50.84 (10.00) | 49.08 (9.90) | 0.002 | 50.15 (10.94) | 49.78 (10.29) | 49.04 (9.52) | 50.95 (9.77) | 0.06 |
| **symr4sec** | | | | | | | | | | | | | |
| SS unadj | 12.23 (2.66) | 10.88 (2.44) | 9.31 (2.58) | 7.64 (2.75) | <0.001 | 10.20 (2.94) | 9.79 (3.09) | 0.02 | 9.54 (3.16) | 9.86 (2.92) | 10.05 (3.05) | 10.22 (3.00) | 0.10 |
| T-ASE adj | 49.62 (10.23) | 50.57 (9.54) | 49.60 (10.01) | 50.13 (10.50) | 0.53 | 49.97 (9.76) | 50.03 (10.29) | 0.92 | 50.70 (10.37) | 50.10 (9.93) | 49.27 (10.01) | 50.36 (9.95) | 0.36 |
| T-AS adj | 49.55 (9.89) | 50.55 (9.81) | 49.62 (10.20) | 50.20 (10.14) | 0.52 | 50.00 (10.16) | 50.01 (9.86) | 1.00 | 50.13 (11.38) | 49.78 (10.16) | 49.36 (9.66) | 50.72 (9.69) | 0.28 |
| T-A adj | 49.60 (9.85) | 50.52 (9.85) | 49.59 (10.24) | 50.17 (10.09) | 0.55 | 50.67 (9.73) | 49.27 (10.27) | 0.01 | 50.24 (11.31) | 49.93 (10.13) | 49.20 (9.62) | 50.68 (9.80) | 0.23 |
| **symmiddle2secavg** | | | | | | | | | | | | | |
| SS unadj | 12.37 (2.63) | 10.84 (2.46) | 9.36 (2.56) | 7.50 (2.57) | <0.001 | 10.25 (3.01) | 9.74 (3.00) | 0.003 | 9.56 (3.01) | 9.82 (2.92) | 10.05 (3.12) | 10.24 (2.98) | 0.08 |
| T-ASE adj | 49.68 (10.20) | 50.30 (9.69) | 49.88 (10.21) | 50.03 (9.98) | 0.89 | 49.98 (9.91) | 50.02 (10.10) | 0.95 | 50.88 (9.95) | 49.90 (9.99) | 49.28 (10.28) | 50.43 (9.73) | 0.28 |
| T-AS adj | 49.67 (10.21) | 50.27 (9.72) | 49.89 (10.14) | 50.09 (10.09) | 0.90 | 50.00 (9.95) | 50.01 (10.08) | 0.99 | 50.22 (10.96) | 49.53 (10.08) | 49.33 (9.91) | 50.91 (9.66) | 0.12 |
| T-A adj | 49.68 (9.80) | 50.24 (9.96) | 49.88 (10.37) | 50.10 (9.70) | 0.91 | 50.91 (9.87) | 49.03 (10.05) | <0.001 | 50.45 (10.95) | 49.76 (10.18) | 49.20 (9.87) | 50.75 (9.60) | 0.16 |
| **symall4secavg** | | | | | | | | | | | | | |
| SS unadj | 12.36 (2.64) | 10.83 (2.42) | 9.38 (2.58) | 7.47 (2.57) | <0.001 | 10.25 (3.02) | 9.74 (2.99) | 0.003 | 9.45 (3.07) | 9.81 (2.90) | 10.06 (3.09) | 10.28 (2.99) | 0.02 |
| T-ASE adj | 49.68 (10.31) | 50.25 (9.55) | 49.96 (10.28) | 49.93 (9.99) | 0.92 | 49.97 (9.89) | 50.01 (10.13) | 0.94 | 50.72 (9.91) | 49.99 (9.88) | 49.29 (10.27) | 50.37 (9.88) | 0.36 |
| T-AS adj | 49.71 (10.33) | 50.24 (9.59) | 49.96 (10.18) | 50.02 (10.10) | 0.94 | 50.01 (9.92) | 50.01 (10.11) | 0.99 | 49.78 (11.22) | 49.48 (9.96) | 49.39 (9.74) | 51.07 (9.79) | 0.07 |
| T-A adj | 49.73 (10.34) | 50.17 (9.61) | 49.98 (10.23) | 50.02 (10.00) | 0.96 | 50.89 (9.87) | 49.05 (10.08) | 0.001 | 49.99 (11.34) | 49.68 (9.97) | 49.22 (9.72) | 50.95 (9.77) | 0.10 |

† p-value was an ANOVA for age and education, and a 2-sample t-test for sex

**Table S6.** Subsample n's for each level of age/education/sex shown in Supplemental Figures 1 and 2.

| Age Group | Education | F | M |
|---|---|---|---|
| 30-59 | 10-12 | 10* | 6** |
| | 13-15 | 33 | 14* |
| | 16 | 42 | 39 |
| | 17-20 | 44 | 45 |
| 60-69 | 10-12 | 17* | 16* |
| | 13-15 | 64 | 36 |
| | 16 | 46 | 77 |
| | 17-20 | 59 | 60 |
| 70-79 | 10-12 | 31 | 15* |
| | 13-15 | 67 | 42 |
| | 16 | 48 | 63 |
| | 17-20 | 51 | 68 |
| 80+ | 10-12 | 32 | 13* |
| | 13-15 | 34 | 28 |
| | 16 | 26 | 32 |
| | 17-20 | 35 | 47 |

*Note*. The confidence intervals displayed in Supplemental Figure 2 assumes normality, so subgroups with n < 20* should be viewed with caution, and results from the one subgroup with n < 10** should be viewed with significant caution.

**Table S7.** Proportion of individuals in the normative sample (far left column) and in the 4 independent validation samples with normative scores < -1 standard deviation (SD; T < 40; SS < 7).

| S7. Characteristic | Norm sample N=1240 | Conc CU N=167 | Norm v CCU p | Dis CU N=149 | MCI N=64 | DEM N=14 | CCU v MCI p | CCU v DEM p | MCI vs DEM p | CCU vs DCU p | DCU vs MCI p |
|---|---|---|---|---|---|---|---|---|---|---|---|
| **mtdrawcompositev1** | | | | | | | | | | | |
| TS-ASE <40 | 173 (14.0%) | 29 (17.4%) | 0.24 | 64 (43.0%) | 40 (62.5%) | 10 (71.4%) | <0.001 | <0.001 | 0.76 | <0.001 | 0.009 |
| *95% CI* | 12.1% - 16.0% | 11.9% - 24.0% | | 34.9% - 51.3% | 49.5% - 74.3% | 41.9% - 91.6% | | | | | |
| TS-AS <40 | 178 (14.4%) | 31 (18.6%) | 0.15 | 65 (43.6%) | 45 (70.3%) | 9 (64.3%) | <0.001 | <0.001 | 0.66 | <0.001 | <0.001 |
| *95% CI* | 12.4% - 16.4% | 13.0% - 25.3% | | 35.5% - 52.0% | 57.6% - 81.1% | 35.1% - 87.2% | | | | | |
| TS-A <40 | 175 (14.1%) | 33 (19.8%) | 0.054 | 67 (45.0%) | 48 (75.0%) | 10 (71.4%) | <0.001 | <0.001 | 0.75 | <0.001 | <0.001 |
| *95% CI* | 12.2% - 16.2% | 14.0% - 26.6% | | 36.8% - 53.3% | 62.6% - 85.0% | 41.9% - 91.6% | | | | | |
| SS <7 | 152 (12.3%) | 25 (15.0%) | 0.32 | 68 (45.6%) | 48 (75.0%) | 12 (85.7%) | <0.001 | <0.001 | 0.50 | <0.001 | <0.001 |
| *95% CI* | 10.5% - 14.2% | 9.9% - 21.3% | | 37.5% - 54.0% | 62.6% - 85.0% | 57.2% - 98.2% | | | | | |
| **slssumoftrials** | | | | | | | | | | | |
| TS-ASE <40 | 173 (14.0%) | 33 (19.8%) | 0.046 | 58 (38.9%) | 41 (64.1%) | 10 (71.4%) | <0.001 | <0.001 | 0.76 | <0.001 | <0.001 |
| *95% CI* | 12.1% - 16.0% | 14.0% - 26.6% | | 31.1% - 47.2% | 51.1% - 75.7% | 41.9% - 91.6% | | | | | |
| TS-AS <40 | 165 (13.3%) | 33 (19.8%) | 0.02 | 64 (43.0%) | 47 (73.4%) | 10 (71.4%) | <0.001 | <0.001 | 1.00 | <0.001 | <0.001 |
| *95% CI* | 11.5% - 15.3% | 14.0% - 26.6% | | 34.9% - 51.3% | 60.9% - 83.7% | 41.9% - 91.6% | | | | | |
| TS-A <40 | 191 (15.4%) | 35 (21.0%) | 0.07 | 64 (43.0%) | 45 (70.3%) | 12 (85.7%) | <0.001 | <0.001 | 0.33 | <0.001 | <0.001 |
| *95% CI* | 13.4% - 17.5% | 15.1% - 27.9% | | 34.9% - 51.3% | 57.6% - 81.1% | 57.2% - 98.2% | | | | | |
| SS <7 | 145 (11.7%) | 24 (14.4%) | 0.32 | 65 (43.6%) | 45 (70.3%) | 11 (78.6%) | <0.001 | <0.001 | 0.75 | <0.001 | <0.001 |
| *95% CI* | 10.0% - 13.6% | 9.4% - 20.6% | | 35.5% - 52.0% | 57.6% - 81.1% | 49.2% - 95.3% | | | | | |
| **symall4weighted** | | | | | | | | | | | |
| TS-ASE <40 | 169 (13.6%) | 26 (15.6%) | 0.50 | 40 (26.8%) | 28 (43.8%) | 9 (64.3%) | <0.001 | <0.001 | 0.16 | 0.01 | 0.02 |
| *95% CI* | 11.8% - 15.7% | 10.4% - 22.0% | | 19.9% - 34.7% | 31.4% - 56.7% | 35.1% - 87.2% | | | | | |
| TS-AS <40 | 174 (14.0%) | 24 (14.4%) | 0.91 | 45 (30.2%) | 30 (46.9%) | 10 (71.4%) | <0.001 | <0.001 | 0.14 | <0.001 | 0.02 |
| *95% CI* | 12.1% - 16.1% | 9.4% - 20.6% | | 23.0% - 38.3% | 34.3% - 59.8% | 41.9% - 91.6% | | | | | |
| TS-A <40 | 180 (14.5%) | 24 (14.4%) | 0.96 | 44 (29.5%) | 30 (46.9%) | 9 (64.3%) | <0.001 | <0.001 | 0.24 | 0.001 | 0.01 |
| *95% CI* | 12.6% - 16.6% | 9.4% - 20.6% | | 22.3% - 37.5% | 34.3% - 59.8% | 35.1% - 87.2% | | | | | |
| SS <7 | 152 (12.3%) | 21 (12.6%) | 0.91 | 47 (31.5%) | 38 (59.4%) | 10 (71.4%) | <0.001 | <0.001 | 0.55 | <0.001 | <0.001 |

| S7. Characteristic | Norm sample N=1240 | Conc CU N=167 | Norm v CCU p | Dis CU N=149 | MCI N=64 | DEM N=14 | CCU v MCI p | CCU v DEM p | MCI vs DEM p | CCU vs DCU p | DCU vs MCI p |
|---|---|---|---|---|---|---|---|---|---|---|---|
| *95% CI* | 10.5% - 14.2% | 8.0% - 18.6% | | 24.2% - 39.7% | 46.4% - 71.5% | 41.9% - 91.6% | | | | | |
| **slsr1corr** | | | | | | | | | | | |
| TS-ASE <40 | 182 (14.7%) | 24 (14.4%) | 0.92 | 43 (28.9%) | 29 (45.3%) | 8 (57.1%) | <0.001 | <0.001 | 0.42 | 0.002 | 0.02 |
| *95% CI* | 12.8% - 16.8% | 9.4% - 20.6% | | 21.7% - 36.8% | 32.8% - 58.3% | 28.9% - 82.3% | | | | | |
| TS-AS <40 | 183 (14.8%) | 24 (14.4%) | 0.89 | 46 (30.9%) | 30 (46.9%) | 8 (57.1%) | <0.001 | <0.001 | 0.49 | <0.001 | 0.03 |
| *95% CI* | 12.8% - 16.9% | 9.4% - 20.6% | | 23.6% - 39.0% | 34.3% - 59.8% | 28.9% - 82.3% | | | | | |
| TS-A <40 | 184 (14.8%) | 25 (15.0%) | 0.96 | 46 (30.9%) | 30 (46.9%) | 8 (57.1%) | <0.001 | <0.001 | 0.49 | <0.001 | 0.03 |
| *95% CI* | 12.9% - 16.9% | 9.9% - 21.3% | | 23.6% - 39.0% | 34.3% - 59.8% | 28.9% - 82.3% | | | | | |
| SS <7 | 230 (18.5%) | 32 (19.2%) | 0.85 | 57 (38.3%) | 33 (51.6%) | 9 (64.3%) | <0.001 | <0.001 | 0.39 | <0.001 | 0.07 |
| *95% CI* | 16.4% - 20.8% | 13.5% - 26.0% | | 30.4% - 46.6% | 38.7% - 64.2% | 35.1% - 87.2% | | | | | |
| **slsr2corr** | | | | | | | | | | | |
| TS-ASE <40 | 208 (16.8%) | 31 (18.6%) | 0.56 | 48 (32.2%) | 31 (48.4%) | 10 (71.4%) | <0.001 | <0.001 | 0.15 | 0.005 | 0.02 |
| *95% CI* | 14.7% - 19.0% | 13.0% - 25.3% | | 24.8% - 40.4% | 35.8% - 61.3% | 41.9% - 91.6% | | | | | |
| TS-AS <40 | 210 (16.9%) | 30 (18.0%) | 0.74 | 55 (36.9%) | 32 (50.0%) | 10 (71.4%) | <0.001 | <0.001 | 0.24 | <0.001 | 0.07 |
| *95% CI* | 14.9% - 19.1% | 12.5% - 24.6% | | 29.2% - 45.2% | 37.2% - 62.8% | 41.9% - 91.6% | | | | | |
| TS-A <40 | 210 (16.9%) | 33 (19.8%) | 0.36 | 62 (41.6%) | 34 (53.1%) | 10 (71.4%) | <0.001 | <0.001 | 0.25 | <0.001 | 0.12 |
| *95% CI* | 14.9% - 19.1% | 14.0% - 26.6% | | 33.6% - 50.0% | 40.2% - 65.7% | 41.9% - 91.6% | | | | | |
| SS <7 | 158 (12.7%) | 26 (15.6%) | 0.31 | 59 (39.6%) | 38 (59.4%) | 11 (78.6%) | <0.001 | <0.001 | 0.23 | <0.001 | 0.008 |
| *95% CI* | 10.9% - 14.7% | 10.4% - 22.0% | | 31.7% - 47.9% | 46.4% - 71.5% | 49.2% - 95.3% | | | | | |
| **slsr3corr** | | | | | | | | | | | |
| TS-ASE <40 | 202 (16.3%) | 30 (18.0%) | 0.58 | 56 (37.6%) | 36 (56.3%) | 10 (71.4%) | <0.001 | <0.001 | 0.38 | <0.001 | 0.01 |
| *95% CI* | 14.3% - 18.5% | 12.5% - 24.6% | | 29.8% - 45.9% | 43.3% - 68.6% | 41.9% - 91.6% | | | | | |
| TS-AS <40 | 191 (15.4%) | 32 (19.2%) | 0.21 | 59 (39.6%) | 42 (65.6%) | 10 (71.4%) | <0.001 | <0.001 | 0.76 | <0.001 | <0.001 |
| *95% CI* | 13.4% - 17.5% | 13.5% - 26.0% | | 31.7% - 47.9% | 52.7% - 77.1% | 41.9% - 91.6% | | | | | |
| TS-A <40 | 202 (16.3%) | 30 (18.0%) | 0.58 | 61 (40.9%) | 38 (59.4%) | 10 (71.4%) | <0.001 | <0.001 | 0.55 | <0.001 | 0.01 |
| *95% CI* | 14.3% - 18.5% | 12.5% - 24.6% | | 33.0% - 49.3% | 46.4% - 71.5% | 41.9% - 91.6% | | | | | |
| SS <7 | 174 (14.0%) | 26 (15.6%) | 0.59 | 64 (43.0%) | 43 (67.2%) | 11 (78.6%) | <0.001 | <0.001 | 0.53 | <0.001 | 0.001 |
| *95% CI* | 12.1% - 16.1% | 10.4% - 22.0% | | 34.9% - 51.3% | 54.3% - 78.4% | 49.2% - 95.3% | | | | | |

| S7. Characteristic | Norm sample N=1240 | Conc CU N=167 | Norm v CCU p | Dis CU N=149 | MCI N=64 | DEM N=14 | CCU v MCI p | CCU v DEM p | MCI vs DEM p | CCU vs DCU p | DCU vs MCI p |
|---|---|---|---|---|---|---|---|---|---|---|---|
| **slsr4corr** | | | | | | | | | | | |
| TS-ASE <40 | 176 (14.2%) | 28 (16.8%) | 0.38 | 61 (40.9%) | 36 (56.3%) | 9 (64.3%) | <0.001 | <0.001 | 0.58 | <0.001 | 0.04 |
| *95% CI* | 12.3% - 16.3% | 11.4% - 23.3% | | 33.0% - 49.3% | 43.3% - 68.6% | 35.1% - 87.2% | | | | | |
| TS-AS <40 | 181 (14.6%) | 31 (18.6%) | 0.18 | 62 (41.6%) | 39 (60.9%) | 9 (64.3%) | <0.001 | <0.001 | 0.82 | <0.001 | <0.01 |
| *95% CI* | 12.7% - 16.7% | 13.0% - 25.3% | | 33.6% - 50.0% | 47.9% - 72.9% | 35.1% - 87.2% | | | | | |
| TS-A <40 | 189 (15.2%) | 32 (19.2%) | 0.19 | 63 (42.3%) | 41 (64.1%) | 11 (78.6%) | <0.001 | <0.001 | 0.36 | <0.001 | 0.004 |
| *95% CI* | 13.3% - 17.4% | 13.5% - 26.0% | | 34.2% - 50.6% | 51.1% - 75.7% | 49.2% - 95.3% | | | | | |
| SS <7 | 161 (13.0%) | 21 (12.6%) | 0.88 | 61 (40.9%) | 42 (65.6%) | 11 (78.6%) | <0.001 | <0.001 | 0.53 | <0.001 | <0.001 |
| *95% CI* | 11.2% - 15.0% | 8.0% - 18.6% | | 33.0% - 49.3% | 52.7% - 77.1% | 49.2% - 95.3% | | | | | |
| **slsr5corr** | | | | | | | | | | | |
| TS-ASE <40 | 179 (14.4%) | 31 (18.6%) | 0.16 | 47 (31.5%) | 35 (54.7%) | 12 (85.7%) | <0.001 | <0.001 | 0.04 | 0.008 | 0.002 |
| *95% CI* | 12.5% - 16.5% | 13.0% - 25.3% | | 24.2% - 39.7% | 41.7% - 67.2% | 57.2% - 98.2% | | | | | |
| TS-AS <40 | 168 (13.5%) | 31 (18.6%) | 0.08 | 58 (38.9%) | 42 (65.6%) | 10 (71.4%) | <0.001 | <0.001 | 0.76 | <0.001 | <0.001 |
| *95% CI* | 11.7% - 15.6% | 13.0% - 25.3% | | 31.1% - 47.2% | 52.7% - 77.1% | 41.9% - 91.6% | | | | | |
| TS-A <40 | 190 (15.3%) | 36 (21.6%) | 0.04 | 57 (38.3%) | 43 (67.2%) | 11 (78.6%) | <0.001 | <0.001 | 0.53 | 0.001 | <0.001 |
| *95% CI* | 13.4% - 17.4% | 15.6% - 28.6% | | 30.4% - 46.6% | 54.3% - 78.4% | 49.2% - 95.3% | | | | | |
| SS <7 | 148 (11.9%) | 25 (15.0%) | 0.26 | 59 (39.6%) | 43 (67.2%) | 12 (85.7%) | <0.001 | <0.001 | 0.21 | <0.001 | <0.001 |
| *95% CI* | 10.2% - 13.9% | 9.9% - 21.3% | | 31.7% - 47.9% | 54.3% - 78.4% | 57.2% - 98.2% | | | | | |
| **slsmaxspan** | | | | | | | | | | | |
| TS-ASE <40 | 168 (13.5%) | 30 (18.0%) | 0.12 | 55 (36.9%) | 41 (64.1%) | 11 (78.6%) | <0.001 | <0.001 | 0.36 | <0.001 | <0.001 |
| *95% CI* | 11.7% - 15.6% | 12.5% - 24.6% | | 29.2% - 45.2% | 51.1% - 75.7% | 49.2% - 95.3% | | | | | |
| TS-AS <40 | 164 (13.2%) | 31 (18.6%) | 0.06 | 59 (39.6%) | 42 (65.6%) | 11 (78.6%) | <0.001 | <0.001 | 0.53 | <0.001 | <0.001 |
| *95% CI* | 11.4% - 15.2% | 13.0% - 25.3% | | 31.7% - 47.9% | 52.7% - 77.1% | 49.2% - 95.3% | | | | | |
| TS-A <40 | 180 (14.5%) | 30 (18.0%) | 0.24 | 64 (43.0%) | 44 (68.8%) | 12 (85.7%) | <0.001 | <0.001 | 0.33 | <0.001 | <0.001 |
| *95% CI* | 12.6% - 16.6% | 12.5% - 24.6% | | 34.9% - 51.3% | 55.9% - 79.8% | 57.2% - 98.2% | | | | | |
| SS <7 | 154 (12.4%) | 24 (14.4%) | 0.48 | 64 (43.0%) | 46 (71.9%) | 11 (78.6%) | <0.001 | <0.001 | 0.75 | <0.001 | <0.001 |
| *95% CI* | 10.6% - 14.4% | 9.4% - 20.6% | | 34.9% - 51.3% | 59.2% - 82.4% | 49.2% - 95.3% | | | | | |
| **slstotcorr** | | | | | | | | | | | |

| S7. Characteristic | Norm sample N=1240 | Conc CU N=167 | Norm v CCU p | Dis CU N=149 | MCI N=64 | DEM N=14 | CCU v MCI p | CCU v DEM p | MCI vs DEM p | CCU vs DCU p | DCU vs MCI p |
|---|---|---|---|---|---|---|---|---|---|---|---|
| TS-ASE <40 | 183 (14.8%) | 31 (18.6%) | 0.20 | 58 (38.9%) | 40 (62.5%) | 11 (78.6%) | <0.001 | <0.001 | 0.36 | <0.001 | 0.002 |
| *95% CI* | 12.8% - 16.9% | 13.0% - 25.3% | | 31.1% - 47.2% | 49.5% - 74.3% | 49.2% - 95.3% | | | | | |
| TS-AS <40 | 179 (14.4%) | 33 (19.8%) | 0.07 | 62 (41.6%) | 44 (68.8%) | 11 (78.6%) | <0.001 | <0.001 | 0.54 | <0.001 | <0.001 |
| *95% CI* | 12.5% - 16.5% | 14.0% - 26.6% | | 33.6% - 50.0% | 55.9% - 79.8% | 49.2% - 95.3% | | | | | |
| TS-A <40 | 184 (14.8%) | 35 (21.0%) | 0.04 | 65 (43.6%) | 43 (67.2%) | 11 (78.6%) | <0.001 | <0.001 | 0.53 | <0.001 | 0.002 |
| *95% CI* | 12.9% - 16.9% | 15.1% - 27.9% | | 35.5% - 52.0% | 54.3% - 78.4% | 49.2% - 95.3% | | | | | |
| SS <7 | 147 (11.9%) | 21 (12.6%) | 0.79 | 60 (40.3%) | 44 (68.8%) | 11 (78.6%) | <0.001 | <0.001 | 0.54 | <0.001 | <0.001 |
| *95% CI* | 10.1% - 13.8% | 8.0% - 18.6% | | 32.3% - 48.6% | 55.9% - 79.8% | 49.2% - 95.3% | | | | | |
| **slsretention** | | | | | | | | | | | |
| TS-ASE <40 | 183 (14.8%) | 27 (16.2%) | 0.63 | 41 (27.5%) | 23 (35.9%) | 6 (42.9%) | 0.001 | 0.01 | 0.63 | 0.01 | 0.22 |
| *95% CI* | 12.8% - 16.9% | 10.9% - 22.6% | | 20.5% - 35.4% | 24.3% - 48.9% | 17.7% - 71.1% | | | | | |
| TS-AS <40 | 186 (15.0%) | 26 (15.6%) | 0.85 | 45 (30.2%) | 24 (37.5%) | 6 (42.9%) | <0.001 | 0.01 | 0.71 | 0.002 | 0.30 |
| *95% CI* | 13.1% - 17.1% | 10.4% - 22.0% | | 23.0% - 38.3% | 25.7% - 50.5% | 17.7% - 71.1% | | | | | |
| TS-A <40 | 179 (14.4%) | 27 (16.2%) | 0.55 | 42 (28.2%) | 26 (40.6%) | 6 (42.9%) | <0.001 | 0.01 | 0.88 | <0.01 | 0.07 |
| *95% CI* | 12.5% - 16.5% | 10.9% - 22.6% | | 21.1% - 36.1% | 28.5% - 53.6% | 17.7% - 71.1% | | | | | |
| SS <7 | 154 (12.4%) | 21 (12.6%) | 0.95 | 39 (26.2%) | 26 (40.6%) | 6 (42.9%) | <0.001 | 0.002 | 0.88 | 0.002 | 0.04 |
| *95% CI* | 10.6% - 14.4% | 8.0% - 18.6% | | 19.3% - 34.0% | 28.5% - 53.6% | 17.7% - 71.1% | | | | | |
| **slsdelaycorr** | | | | | | | | | | | |
| TS-ASE <40 | 177 (14.3%) | 35 (21.0%) | 0.02 | 51 (34.2%) | 40 (62.5%) | 10 (71.4%) | <0.001 | <0.001 | 0.76 | 0.008 | <0.001 |
| *95% CI* | 12.4% - 16.3% | 15.1% - 27.9% | | 26.7% - 42.4% | 49.5% - 74.3% | 41.9% - 91.6% | | | | | |
| TS-AS <40 | 178 (14.4%) | 35 (21.0%) | 0.03 | 63 (42.3%) | 42 (65.6%) | 10 (71.4%) | <0.001 | <0.001 | 0.76 | <0.001 | 0.002 |
| *95% CI* | 12.4% - 16.4% | 15.1% - 27.9% | | 34.2% - 50.6% | 52.7% - 77.1% | 41.9% - 91.6% | | | | | |
| TS-A <40 | 167 (13.5%) | 29 (17.4%) | 0.17 | 60 (40.3%) | 46 (71.9%) | 12 (85.7%) | <0.001 | <0.001 | 0.50 | <0.001 | <0.001 |
| *95% CI* | 11.6% - 15.5% | 11.9% - 24.0% | | 32.3% - 48.6% | 59.2% - 82.4% | 57.2% - 98.2% | | | | | |
| SS <7 | 160 (12.9%) | 29 (17.4%) | 0.11 | 71 (47.7%) | 45 (70.3%) | 11 (78.6%) | <0.001 | <0.001 | 0.75 | <0.001 | 0.002 |
| *95% CI* | 11.1% - 14.9% | 11.9% - 24.0% | | 39.4% - 56.0% | 57.6% - 81.1% | 49.2% - 95.3% | | | | | |
| **symsumcorr** | | | | | | | | | | | |
| TS-ASE <40 | 212 (17.1%) | 37 (22.2%) | 0.11 | 36 (24.2%) | 15 (23.4%) | 5 (35.7%) | 0.83 | 0.25 | 0.34 | 0.67 | 0.91 |

| S7. Characteristic | Norm sample N=1240 | Conc CU N=167 | Norm v CCU p | Dis CU N=149 | MCI N=64 | DEM N=14 | CCU v MCI p | CCU v DEM p | MCI vs DEM p | CCU vs DCU p | DCU vs MCI p |
|---|---|---|---|---|---|---|---|---|---|---|---|
| *95% CI* | 15.0% - 19.3% | 16.1% - 29.2% | | 17.5% - 31.8% | 13.8% - 35.7% | 12.8% - 64.9% | | | | | |
| TS-AS <40 | 214 (17.3%) | 37 (22.2%) | 0.12 | 42 (28.2%) | 18 (28.1%) | 5 (35.7%) | 0.34 | 0.25 | 0.57 | 0.22 | 0.99 |
| *95% CI* | 15.2% - 19.5% | 16.1% - 29.2% | | 21.1% - 36.1% | 17.6% - 40.8% | 12.8% - 64.9% | | | | | |
| TS-A <40 | 215 (17.3%) | 37 (22.2%) | 0.13 | 43 (28.9%) | 18 (28.1%) | 5 (35.7%) | 0.34 | 0.25 | 0.57 | 0.17 | 0.91 |
| *95% CI* | 15.3% - 19.6% | 16.1% - 29.2% | | 21.7% - 36.8% | 17.6% - 40.8% | 12.8% - 64.9% | | | | | |
| SS <7 | 222 (17.9%) | 37 (22.2%) | 0.18 | 43 (28.9%) | 20 (31.3%) | 5 (35.7%) | 0.15 | 0.25 | 0.75 | 0.17 | 0.73 |
| *95% CI* | 15.8% - 20.2% | 16.1% - 29.2% | | 21.7% - 36.8% | 20.2% - 44.1% | 12.8% - 64.9% | | | | | |
| **symr1sec** | | | | | | | | | | | |
| TS-ASE <40 | 170 (13.7%) | 23 (13.8%) | 0.98 | 33 (22.1%) | 23 (35.9%) | 10 (71.4%) | <0.001 | <0.001 | 0.02 | 0.052 | 0.04 |
| *95% CI* | 11.8% - 15.8% | 8.9% - 19.9% | | 15.8% - 29.7% | 24.3% - 48.9% | 41.9% - 91.6% | | | | | |
| TS-AS <40 | 173 (14.0%) | 23 (13.8%) | 0.95 | 35 (23.5%) | 25 (39.1%) | 10 (71.4%) | <0.001 | <0.001 | 0.04 | 0.03 | 0.02 |
| *95% CI* | 12.1% - 16.0% | 8.9% - 19.9% | | 16.9% - 31.1% | 27.1% - 52.1% | 41.9% - 91.6% | | | | | |
| TS-A <40 | 172 (13.9%) | 22 (13.2%) | 0.81 | 35 (23.5%) | 26 (40.6%) | 10 (71.4%) | <0.001 | <0.001 | 0.04 | 0.02 | 0.01 |
| *95% CI* | 12.0% - 15.9% | 8.4% - 19.3% | | 16.9% - 31.1% | 28.5% - 53.6% | 41.9% - 91.6% | | | | | |
| SS <7 | 152 (12.3%) | 22 (13.2%) | 0.74 | 37 (24.8%) | 38 (59.4%) | 10 (71.4%) | <0.001 | <0.001 | 0.55 | 0.008 | <0.001 |
| *95% CI* | 10.5% - 14.2% | 8.4% - 19.3% | | 18.1% - 32.6% | 46.4% - 71.5% | 41.9% - 91.6% | | | | | |
| **symr2sec** | | | | | | | | | | | |
| TS-ASE <40 | 186 (15.0%) | 24 (14.4%) | 0.83 | 37 (24.8%) | 35 (54.7%) | 9 (64.3%) | <0.001 | <0.001 | 0.51 | 0.02 | <0.001 |
| *95% CI* | 13.1% - 17.1% | 9.4% - 20.6% | | 18.1% - 32.6% | 41.7% - 67.2% | 35.1% - 87.2% | | | | | |
| TS-AS <40 | 185 (14.9%) | 21 (12.6%) | 0.42 | 39 (26.2%) | 34 (53.1%) | 9 (64.3%) | <0.001 | <0.001 | 0.45 | 0.002 | <0.001 |
| *95% CI* | 13.0% - 17.0% | 8.0% - 18.6% | | 19.3% - 34.0% | 40.2% - 65.7% | 35.1% - 87.2% | | | | | |
| TS-A <40 | 184 (14.8%) | 22 (13.2%) | 0.57 | 37 (24.8%) | 35 (54.7%) | 8 (57.1%) | <0.001 | <0.001 | 0.87 | 0.008 | <0.001 |
| *95% CI* | 12.9% - 16.9% | 8.4% - 19.3% | | 18.1% - 32.6% | 41.7% - 67.2% | 28.9% - 82.3% | | | | | |
| SS <7 | 152 (12.3%) | 30 (18.0%) | 0.04 | 43 (28.9%) | 36 (56.3%) | 9 (64.3%) | <0.001 | <0.001 | 0.58 | 0.02 | <0.001 |
| *95% CI* | 10.5% - 14.2% | 12.5% - 24.6% | | 21.7% - 36.8% | 43.3% - 68.6% | 35.1% - 87.2% | | | | | |
| **symr3sec** | | | | | | | | | | | |
| TS-ASE <40 | 177 (14.3%) | 23 (13.8%) | 0.86 | 38 (25.5%) | 31 (48.4%) | 8 (57.1%) | <0.001 | <0.001 | 0.56 | 0.008 | 0.001 |
| *95% CI* | 12.4% - 16.3% | 8.9% - 19.9% | | 18.7% - 33.3% | 35.8% - 61.3% | 28.9% - 82.3% | | | | | |

| S7. Characteristic | Norm sample N=1240 | Conc CU N=167 | Norm v CCU p | Dis CU N=149 | MCI N=64 | DEM N=14 | CCU v MCI p | CCU v DEM p | MCI vs DEM p | CCU vs DCU p | DCU vs MCI p |
|---|---|---|---|---|---|---|---|---|---|---|---|
| TS-AS <40 | 175 (14.1%) | 23 (13.8%) | 0.91 | 40 (26.8%) | 30 (46.9%) | 8 (57.1%) | <0.001 | <0.001 | 0.49 | 0.004 | 0.004 |
| *95% CI* | 12.2% - 16.2% | 8.9% - 19.9% | | 19.9% - 34.7% | 34.3% - 59.8% | 28.9% - 82.3% | | | | | |
| TS-A <40 | 182 (14.7%) | 24 (14.4%) | 0.92 | 36 (24.2%) | 32 (50.0%) | 7 (50.0%) | <0.001 | <0.001 | 1.00 | 0.03 | <0.001 |
| *95% CI* | 12.8% - 16.8% | 9.4% - 20.6% | | 17.5% - 31.8% | 37.2% - 62.8% | 23.0% - 77.0% | | | | | |
| SS <7 | 152 (12.3%) | 15 (9.0%) | 0.22 | 47 (31.5%) | 39 (60.9%) | 10 (71.4%) | <0.001 | <0.001 | 0.55 | <0.001 | <0.001 |
| *95% CI* | 10.5% - 14.2% | 5.1% - 14.4% | | 24.2% - 39.7% | 47.9% - 72.9% | 41.9% - 91.6% | | | | | |
| **symr4sec** | | | | | | | | | | | |
| TS-ASE <40 | 172 (13.9%) | 26 (15.6%) | 0.55 | 36 (24.2%) | 29 (45.3%) | 9 (64.3%) | <0.001 | <0.001 | 0.20 | 0.055 | 0.002 |
| *95% CI* | 12.0% - 15.9% | 10.4% - 22.0% | | 17.5% - 31.8% | 32.8% - 58.3% | 35.1% - 87.2% | | | | | |
| TS-AS <40 | 176 (14.2%) | 26 (15.6%) | 0.63 | 37 (24.8%) | 30 (46.9%) | 9 (64.3%) | <0.001 | <0.001 | 0.24 | 0.04 | 0.002 |
| *95% CI* | 12.3% - 16.3% | 10.4% - 22.0% | | 18.1% - 32.6% | 34.3% - 59.8% | 35.1% - 87.2% | | | | | |
| TS-A <40 | 179 (14.4%) | 29 (17.4%) | 0.32 | 36 (24.2%) | 33 (51.6%) | 9 (64.3%) | <0.001 | <0.001 | 0.39 | 0.14 | <0.001 |
| *95% CI* | 12.5% - 16.5% | 11.9% - 24.0% | | 17.5% - 31.8% | 38.7% - 64.2% | 35.1% - 87.2% | | | | | |
| SS <7 | 152 (12.3%) | 19 (11.4%) | 0.74 | 45 (30.2%) | 39 (60.9%) | 10 (71.4%) | <0.001 | <0.001 | 0.55 | <0.001 | <0.001 |
| *95% CI* | 10.5% - 14.2% | 7.0% - 17.2% | | 23.0% - 38.3% | 47.9% - 72.9% | 41.9% - 91.6% | | | | | |
| **symmiddle2secavg** | | | | | | | | | | | |
| TS-ASE <40 | 184 (14.8%) | 22 (13.2%) | 0.57 | 35 (23.5%) | 32 (50.0%) | 9 (64.3%) | <0.001 | <0.001 | 0.33 | 0.02 | <0.001 |
| *95% CI* | 12.9% - 16.9% | 8.4% - 19.3% | | 16.9% - 31.1% | 37.2% - 62.8% | 35.1% - 87.2% | | | | | |
| TS-AS <40 | 183 (14.8%) | 21 (12.6%) | 0.45 | 39 (26.2%) | 32 (50.0%) | 9 (64.3%) | <0.001 | <0.001 | 0.33 | 0.002 | <0.001 |
| *95% CI* | 12.8% - 16.9% | 8.0% - 18.6% | | 19.3% - 34.0% | 37.2% - 62.8% | 35.1% - 87.2% | | | | | |
| TS-A <40 | 180 (14.5%) | 19 (11.4%) | 0.27 | 37 (24.8%) | 34 (53.1%) | 9 (64.3%) | <0.001 | <0.001 | 0.45 | 0.002 | <0.001 |
| *95% CI* | 12.6% - 16.6% | 7.0% - 17.2% | | 18.1% - 32.6% | 40.2% - 65.7% | 35.1% - 87.2% | | | | | |
| SS <7 | 152 (12.3%) | 20 (12.0%) | 0.92 | 44 (29.5%) | 38 (59.4%) | 9 (64.3%) | <0.001 | <0.001 | 0.73 | <0.001 | <0.001 |
| *95% CI* | 10.5% - 14.2% | 7.5% - 17.9% | | 22.3% - 37.5% | 46.4% - 71.5% | 35.1% - 87.2% | | | | | |
| **symall4secavg** | | | | | | | | | | | |
| TS-ASE <40 | 176 (14.2%) | 22 (13.2%) | 0.72 | 36 (24.2%) | 27 (42.2%) | 9 (64.3%) | <0.001 | <0.001 | 0.13 | 0.01 | 0.008 |
| *95% CI* | 12.3% - 16.3% | 8.4% - 19.3% | | 17.5% - 31.8% | 29.9% - 55.2% | 35.1% - 87.2% | | | | | |
| TS-AS <40 | 183 (14.8%) | 21 (12.6%) | 0.45 | 42 (28.2%) | 32 (50.0%) | 10 (71.4%) | <0.001 | <0.001 | 0.24 | <0.001 | 0.002 |

| S7. Characteristic | Norm sample N=1240 | Conc CU N=167 | Norm v CCU p | Dis CU N=149 | MCI N=64 | DEM N=14 | CCU v MCI p | CCU v DEM p | MCI vs DEM p | CCU vs DCU p | DCU vs MCI p |
|---|---|---|---|---|---|---|---|---|---|---|---|
| *95% CI* | 12.8% - 16.9% | 8.0% - 18.6% | | 21.1% - 36.1% | 37.2% - 62.8% | 41.9% - 91.6% | | | | | |
| TS-A <40 | 183 (14.8%) | 19 (11.4%) | 0.24 | 40 (26.8%) | 34 (53.1%) | 10 (71.4%) | <0.001 | <0.001 | 0.25 | <0.001 | <0.001 |
| *95% CI* | 12.8% - 16.9% | 7.0% - 17.2% | | 19.9% - 34.7% | 40.2% - 65.7% | 41.9% - 91.6% | | | | | |
| SS <7 | 152 (12.3%) | 19 (11.4%) | 0.74 | 45 (30.2%) | 40 (62.5%) | 9 (64.3%) | <0.001 | <0.001 | 0.90 | <0.001 | <0.001 |
| *95% CI* | 10.5% - 14.2% | 7.0% - 17.2% | | 23.0% - 38.3% | 49.5% - 74.3% | 35.1% - 87.2% | | | | | |
| **symall4corrartsec** | | | | | | | | | | | |
| TS-ASE <40 | 176 (14.2%) | 22 (13.2%) | 0.72 | 35 (23.5%) | 27 (42.2%) | 9 (64.3%) | <0.001 | <0.001 | 0.13 | 0.02 | 0.006 |
| *95% CI* | 12.3% - 16.3% | 8.4% - 19.3% | | 16.9% - 31.1% | 29.9% - 55.2% | 35.1% - 87.2% | | | | | |
| TS-AS <40 | 177 (14.3%) | 21 (12.6%) | 0.55 | 41 (27.5%) | 33 (51.6%) | 10 (71.4%) | <0.001 | <0.001 | 0.24 | <0.001 | <0.001 |
| *95% CI* | 12.4% - 16.3% | 8.0% - 18.6% | | 20.5% - 35.4% | 38.7% - 64.2% | 41.9% - 91.6% | | | | | |
| TS-A <40 | 178 (14.4%) | 20 (12.0%) | 0.41 | 39 (26.2%) | 35 (54.7%) | 10 (71.4%) | <0.001 | <0.001 | 0.37 | 0.001 | <0.001 |
| *95% CI* | 12.4% - 16.4% | 7.5% - 17.9% | | 19.3% - 34.0% | 41.7% - 67.2% | 41.9% - 91.6% | | | | | |
| SS <7 | 152 (12.3%) | 20 (12.0%) | 0.92 | 45 (30.2%) | 39 (60.9%) | 9 (64.3%) | <0.001 | <0.001 | 0.82 | <0.001 | <0.001 |
| *95% CI* | 10.5% - 14.2% | 7.5% - 17.9% | | 23.0% - 38.3% | 47.9% - 72.9% | 35.1% - 87.2% | | | | | |

*Note*. Results are significantly different than expected when 95% confidence intervals (CIs) do not include the expected 14.7% base rate value (see Stricker et al., 2021). Age, sex, and education-adjusted T-scores (TS-ASE) were hypothesized to have CIs that include the expected 14.7% base rate for the concordant CU participants, and to have CIs that did not include the 14.7% base rate for MCI and dementia participants. Discordant CU analyses were exploratory. TS-AS = age and sex-adjusted T-scores. TS-A = age-adjusted T-scores. Unadjusted scaled scores (SS) are presented for reference; these scaled scores place the raw score on a normal distribution but do not adjust for any demographic variables and therefore represent the mean performance of the full normative sample (mean age = 70, 49% male, mean education = 16, N=1240). For example, applying an unadjusted SS < 7 cut-off is equivalent to applying the following raw score cut-offs for the primary variables: MTD Composite < 85, SLS Sum of Trials < 57, $SYM_{AW}$ < 25.5. Concordant CU indicates that participants in the MCSA received an independent diagnosis of CU from the study physician, study coordinator who administered the CDR, and neuropsychologist; discordant CU received a consensus diagnosis of CU but one of the three diagnostic raters did not assign a CU diagnosis. ADRC participants had a consensus conference diagnosis of CU. MCI and dementia represent consensus diagnoses.

**Table S8.** Proportion of individuals in the MCI independent validation sample (N=64) with low test performance (< -1 SD), 95% CI; comparison of varying level of demographic adjustment.

| Characteristic | Age/Sex/Ed T <40 | Age/Sex T <40 | Age T <40 | Unadj. SS <7 |
|---|---|---|---|---|
| **MTD Composite, N (%)** | 40 (62.5%) [a b] | 45 (70.3%) | 48 (75.0%) [a] | 48 (75.0%) [b] |
| *95% CI* | 49.5% - 74.3%* | 57.6% - 81.1%* | 62.6% - 85.0%* | 62.6% - 85.0%* |
| **SLS Sum of Trials, N (%)** | 41 (64.1%) [c] | 47 (73.4%) [c] | 45 (70.3%) | 45 (70.3%) |
| *95% CI* | 51.1% - 75.7%* | 60.9% - 83.7%* | 57.6% - 81.1%* | 57.6% - 81.1%* |
| **SYM acc-weighted, N (%)** | 28 (43.8%) [d] | 30 (46.9%) [e] | 30 (46.9%) [f] | 38 (59.4%) [d e f] |
| *95% CI* | 31.4% - 56.7%* | 34.3% - 59.8%* | 34.3% - 59.8%* | 46.4% - 71.5%* |

*Note*. Age/Sex/Ed T = age, sex and education adjusted T-score (i.e., fully adjusted T); Age/Sex T = age and sex adjusted T-score; age T = age-adjusted T-score; Unadj. SS = unadjusted scaled score. We interpreted McNemar p-values < .05 as evidence of significantly different frequencies of low test scores for cross-norms comparisons (i.e., different sensitivity in the same individuals when different norms are applied); when the same letter appears for horizontal comparisons within the table for a given variable, there is a significant difference. Table used with permission of Mayo Foundation for Medical Education and Research, all rights reserved.

[a] p=0.005
[b] p=0.03
[c] p=0.03
[d] p=0.01
[e] p=0.03
[f] p=0.03

*Results are significantly different than expected base rates when 95% confidence intervals (CIs) do not include the expected 14.7% base rate value (see Stricker et al., 2021). Unadjusted scaled scores are presented for reference; these scaled scores place the raw score on a normal distribution but do not adjust for any demographic variables and therefore represent the mean performance of the full normative sample (mean age = 70, 49% male, mean education = 16, N=1240). Applying an unadjusted SS < 7 cut-off is equivalent to applying the following raw score cut-offs: MTD Composite < 85, SLS Sum of Trials < 57, $SYM_{AW}$ < 25.5.

**Supplemental Figures**

**Figure S1.** Mean (95% CI) MTD unadjusted scaled scores for each primary measure (columns) depicted by sex (men, blue; women, red), education group (10-12, 13-15, 16, and 17-20 years, at right), and age group (30-59, 60-69, 70-79, and 80+ years, at left).

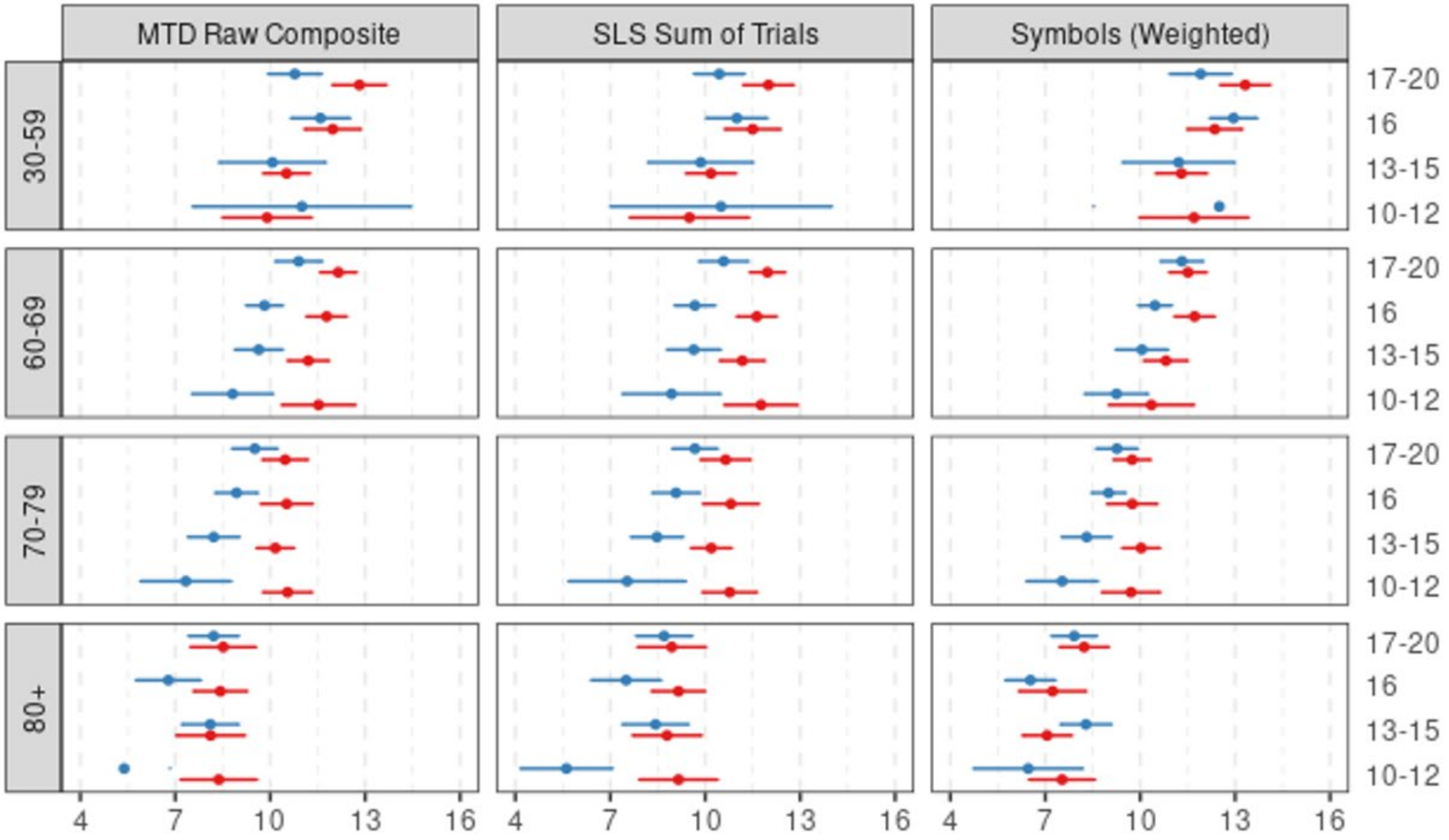

*Note*. Dot only shown for two groups (M 30-59, 10-12 Symbols Accuracy-Weighted and M 80+, 10-12 MTD raw composite) because the scaled scores plot parameters do not cover the full confidence interval for those groups (the other dot falls outside of the plotted range of scaled scores of 4-16). Because scaled scores are unadjusted, they show expected relationships with age, sex and education.

**Figure S2.** Mean (95% CI) MTD T-scores for each primary measure (columns) depicted by sex (men, blue; women, red), education group (10-12, 13-15, 16, and 17-20 years, at right), and age group (30-59, 60-69, 70-79, and 80+ years, at left).

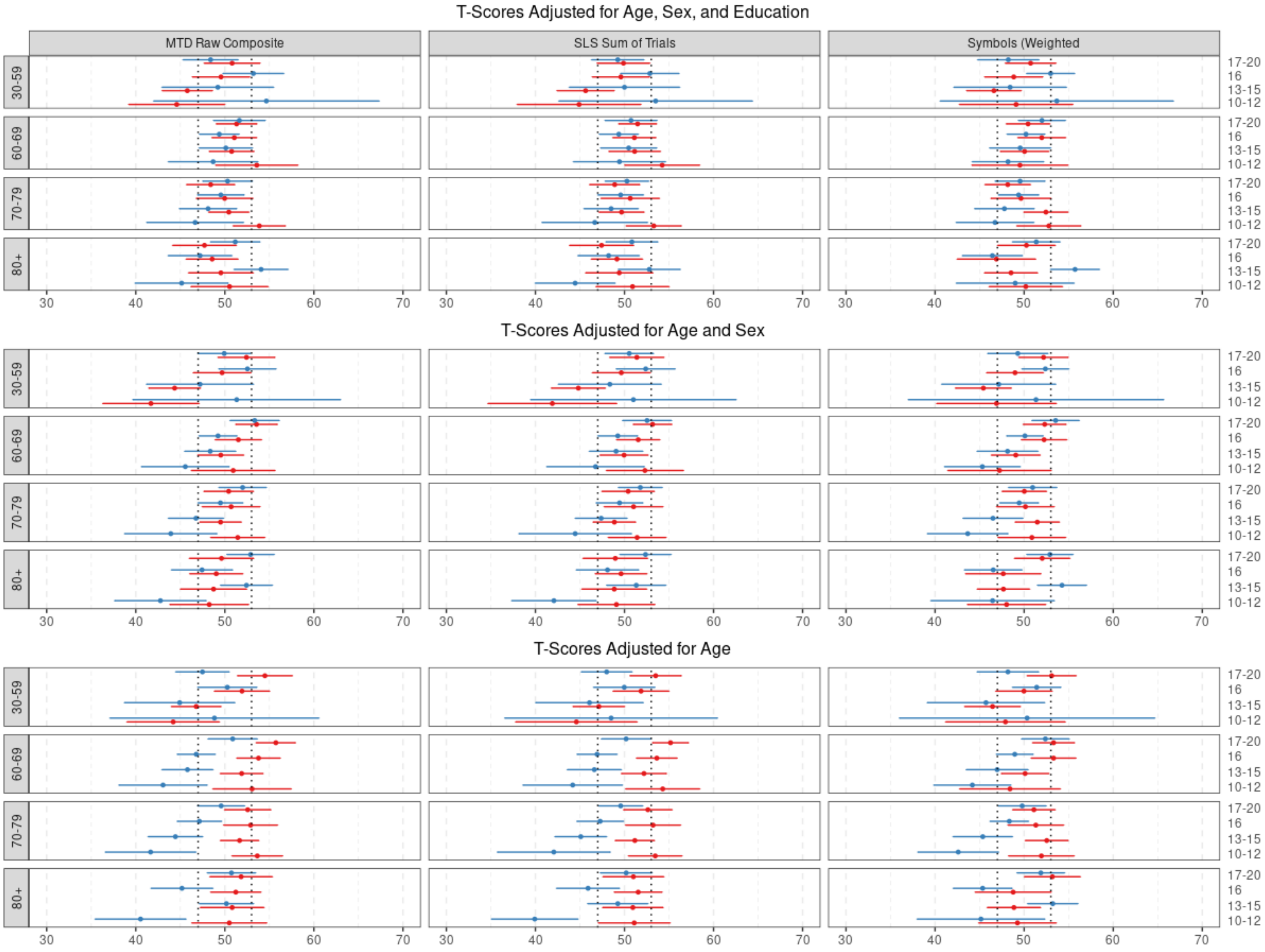

*Note*. This figure illustrates how T-scores perform across the different levels of demographic variables. The goal is for the mean to fall within 47-53 T; boundaries indicated with dotted lines. Full adjustment is shown in the top row, adjustment for age and sex in the next row, and adjustment for age only in the bottom row. For fully-adjusted T-scores, those falling outside the desired boundaries tended to have n<20 in that subcategory (specific age group, sex and education group); see Table S6.
